# Contribution of autosomal rare and *de novo* variants to sex differences in autism

**DOI:** 10.1101/2024.04.13.24305713

**Authors:** Mahmoud Koko, F. Kyle Satterstrom, Autism Sequencing Consortium, APEX consortium, Varun Warrier, Hilary Martin

## Abstract

Autism is four times more prevalent in males than females. To study whether this reflects a difference in genetic predisposition attributed to autosomal rare variants, we evaluated the sex differences in effect size of damaging protein-truncating and missense variants on autism predisposition in 47,061 autistic individuals, then compared effect sizes between individuals with and without cognitive impairment or motor delay. Although these variants mediated differential likelihood of autism with versus without motor or cognitive impairment, their effect sizes on the liability scale did not differ significantly by sex exome-wide or in genes sex-differentially expressed in the cortex. Although de novo mutations were enriched in genes with male-biased expression in the fetal cortex, the liability they conferred did not differ significantly from other genes with similar loss-of-function intolerance and sex-averaged cortical expression. In summary, autosomal rare coding variants confer similar liability for autism in females and males.

## Background

Large-scale rare variant association studies in autism have shown that rare protein-coding variants contribute significantly to autism liability. Genes associated with autism - particularly those with strong statistical or molecular evidence - are enriched for loss-of-function (LoF)-intolerant genes expressed in the brain.^1–3^ It remains unclear whether the rare variant contribution relates to the fact that autism in childhood is approximately four times more prevalent in males. This sex bias is less pronounced when autism is accompanied by cognitive impairment or motor developmental delay.^4,5^

The genetics of autism includes both rare and common variants.^6–8^ The collective effect of the different genetic factors predisposing to autism on the trait prevalence is typically studied using the Liability Threshold Model.^4^ This model is often used to explain how additive genetic factors relate to a dichotomous diagnosis - by postulating that the combined effect of these predisposition factors in the population is normally distributed, and individuals diagnosed with autism will have exceeded a certain diagnostic threshold.^9^ Under this model, a sex difference in the population prevalence may arise from a higher threshold in females relative to males so that females require higher genetic predisposition to be diagnosed,^10^ or a single threshold with sex-biased effect sizes (gene-by-sex interaction) causing the same set of variants to push males but not females past the threshold.^11^ Unlike comparisons on the observed scale (e.g., fold-enrichment in carrier rates in males versus females), examining effect sizes on the liability scale allows for a direct comparison between groups with different proportions of individuals with the trait.

Previous work on 12,270 autistic individuals showed that females have a higher burden of rare damaging variants.^12^ However, it was not clarified if those observed differences translate into differences in liability. A separate analysis of overlapping cohorts (11,986 autistic individuals) by the Autism Sequencing Consortium (ASC) suggested that the observed significant sex differences in *de novo* mutation (DNM) enrichment did not translate °into differences in the average effect size attributed to these damaging variants on the liability scale.^2^ Importantly, autistic individuals show different rates of DNMs and over-transmission of damaging rare alleles depending on the presence of co-occurring cognitive impairment or dysmorphism.^13,14^ More recent ASC work on 21k autistic individuals did not show evidence for gene-by-sex interaction in carriers of rare variants in autosomal genes significantly associated with autism, and suggested that sex and phenotypic severity additively determine rare variant burden in these autism predisposition genes.^1^ Given a larger proportion of autistic females than males have a co-occurring cognitive impairment, potentially because of an underdiagnosis of autistic females with otherwise typical cognitive and motor development,^15,16^ it is unclear if the observed sex difference in rare variant rate is simply a reflection of differences in proportion of individuals with cognitive impairment between sexes.

Larger, more recently released cohorts like the Simons Foundation Powering Autism for Research Knowledge (SPARK), which includes more than 40,000 autistic individuals, offer a chance to examine rare variant liability and its relation to autism and co-occurring conditions in depth. Here we assembled one of the largest datasets of exome-sequenced samples from autism cohorts to date, to explore whether there is a sex difference in rare variant liability, both exome-wide and in focused gene-sets of high-confidence autism-associated genes and genes with sex-biased expression in the fetal and adult cortex. We performed sex-stratified analyses of rare missense and protein-truncating variants in exonic coding regions in 47,061 autistic individuals and 25,593 siblings or controls not diagnosed with autism from cohorts curated by SPARK and the ASC (Supplementary Table S4 in section 4 of the Supplementary Methods; see the Supplementary Note). We then explored the relationship between the sex differences in liability and motor and cognitive difficulties co-occurring with autism in a subset of 27,227 autistic individuals from SPARK.

## Methods

### A note on terminology

We use neutral terminology, including ‘autistic individuals’ throughout the manuscript, in line with preferences from a large number of autistic people. However, we use standard statistical terminology (e.g., liability, Liability Threshold Model, risk ratio, gene burden) to be consistent with other literature.

### Ethics and approvals

We confirm that the datasets used for this study were obtained from research projects complying with relevant ethical regulations. The ASC studies were approved by Mass General Brigham Human Research Committee Institutional Review Board protocols no. 2012P001018 and 2013P000323. Access to SPARK phenotypic and genetic data was approved by the Simons Foundation Autism Research Initiative (SFARI). SPARK participants were recruited under Western Institutional Review Board (IRB) protocol no. 20151664.

### SPARK cohort

The latest release of integrated whole exome sequencing data (iWES2) spanned five sequencing waves (WES1-5) encompassing 106,744 individuals (44,304 of them were diagnosed with autism, the rest non-autistic parents, siblings, and a few extended family members). These included 25,386 trios (18,172 autistic individuals and 7,214 not diagnosed with autism), 23,346 samples with one sequenced parent (17,644 autistic individuals and 5,702 not diagnosed with autism), and 58,012 samples without parental WES (8,488 autistic individuals and 49,524 not diagnosed with autism - with the latter group formed mostly of parents of other individuals in the trio-/pair-sequenced groups i.e. few multi-generational families). The sex-stratified counts of autistic children, siblings not diagnosed with autism, and parents per population are given in Supplementary Table S1 in section 1 of the Supplementary Methods (see the Supplementary Note).

First, we performed exome quality control (QC) on all samples in iWES2, as detailed in section 1 of the Supplementary Methods (see the Supplementary Note). Briefly, we annotated the variant calls with coding consequences on Matched Annotation from NCBI and Ensembl transcripts (MANE; Ensembl release 108; Genome build GRCh38) and filtered for variants having synonymous or more damaging consequences, prioritizing the most severe consequence when two genes were affected. We removed variants failing a Random Forest quality filter, genotypes with low depth (DP < 10), low quality (GQ <10), or low variant allele fraction (VAF < 0.25), and outlier samples on these metrics: total and singleton variant count, transition-transversion ratio, insertion-deletion ratio, and heterozygous-homozygous ratio.

We then excluded all samples that were potentially part of the ASC cohort (by removing all individuals in SPARK who indicated their previous participation in ASC studies), parents and siblings reported to have a developmental disorder/motor delay or cognitive impairment, and autistic parents. We then defined a set of maximally unrelated probands and maximally unrelated siblings by incrementally removing individuals with the highest number of related people (within each of these two subsets), while preferentially retaining females. Following QC, we evaluated the genotypes of 20,236 trio-sequenced individuals (13,473 with autism and 6,763 not diagnosed with autism) to identify rare DNMs and inherited variants (SPARK & gnomAD minor allele frequency < 0.1%).

We also did supplementary analyses in which we examined ultra-rare DNMs (allele frequency < 0.005%) in these 20,236 trio-sequenced individuals, ultra-rare inherited variants (in one SPARK family and not in gnomAD) in an additional 18,816 child-parent pairs with one sequenced parent (13,435 with autism and 5,381 not diagnosed with autism), and ultra-rare variants (allele frequency < 0.005%) of undetermined origin in 8,905 individuals without sequenced parents (6,533 with autism and 2,372 not diagnosed with autism). Further details on rare and ultra-rare variant filtering are available in sections 2 and 3 of the Supplementary Methods (Supplementary Note).

### ASC cohort

The QC of this dataset is described elsewhere in the context of a large rare variant association analysis.^1^ This previous analysis primarily examined both sexes jointly, using data from the SPARK Pilot and first exome sequencing wave (WES1), the Simons Simplex Collection (SSC) and smaller ASC family-based cohorts, and Swedish and Danish case-control cohorts. From the ASC, we received sex-stratified gene-level rare variant counts (gnomAD minor allele frequency < 0.1%) grouped by their mode of inheritance into DNMs (in probands or siblings) or inherited variants (transmitted or untransmitted in the probands). *De novo* mutation (DNM) counts came from 10,488 individuals in the ASC/SSC family-based cohort only (8,028 autistic individuals and 2,460 not diagnosed with autism) and did not include those ascertained in SPARK Pilot and WES1 families, so was independent of the SPARK iWES2 dataset presented in the previous section. Some of the DNMs in the ASC cohort were collated from older studies and did not have accompanying information on inherited alleles. Therefore, the inherited variants were evaluated in 9,929 children (7,570 autistic children and 2,359 siblings). We also obtained ultra-rare variant counts (allele frequency < 0.005%) from 14,188 individuals from the ASC case-control cohorts (5,591 autistic individuals and 8,597 not diagnosed with autism). See sections 2.1 and 3.2 of the Supplementary Methods (Supplementary Note) for more details.

### *De novo* and inherited variants

We analyzed DNMs, rare transmitted variants, and rare untransmitted variants annotated as damaging protein-truncating variants (PTVs), damaging missense variants, or synonymous variants. The analysis was limited to 17,296 protein-coding genes annotated in both the ASC and SPARK after quality control. PTVs were considered damaging if they occurred in 1,739 highly LoF-intolerant genes in the most-constrained decile for Loss-Of-Function Observed over Expected Upper bound Fraction (LOEUF) score, and missense variants were considered damaging if they had a Missense Badness, Polyphen and Constraint (MPC) score ≥ 2 (in all genes). The primary analysis of rare variants (allele frequency < 0.1%) was performed in the trio-sequenced individuals in both SPARK and ASC (21,501 autistic individuals and 9,223 siblings not diagnosed with autism), and the remaining data (individuals with sequence data from one or neither parent) were used for analyses of ultra-rare variants (Supplementary Table S4 in section 4 of the Supplementary Methods; see the Supplementary Note). We carried out sex-stratified comparisons (autistic individuals against sex-matched siblings) as well as direct comparisons between sexes (autistic females versus autistic males), as described in the next section.

### Exome-wide enrichment

The following statistical analyses are described in detail in section 4 of the Supplementary Methods, and summarized briefly here. We used the ratio between the rate of DNMs in the probands (DNMs per sample) and rate of DNMs in the siblings as a measure of enrichment. For inherited variants, we calculated the ratio between parental alleles transmitted to the probands and the remaining untransmitted alleles. A ratio of 1 in the context of DNM analysis means that the probands and the siblings have equal rates of rare DNMs; in the context of transmission analysis, it means that there is transmission equilibrium (half of the rare parental alleles are transmitted to the probands). For simplicity, we may refer to both ratios as the “rate ratio”.

To test for the significance of observed deviations from a DNM rate ratio 1, we used a two-sided binomial exact test to compare the DNM counts in the probands and the siblings. This tested whether the proportion of DNM seen in the probands (from all DNMs in the probands and siblings) is significantly different from the proportion expected given their sample size (expected rate = N_probands_/(N_probands_+N_siblings_)). For inherited variants, we used a two-sided binomial exact test to compare the counts of transmitted and untransmitted alleles in the probands, examining whether the fraction of parental alleles transmitted to the probands was significantly different from 0.5. We obtained the confidence intervals for the rate ratio from these binomial tests. Comparisons of variant rates between cases and controls were evaluated in the same way as DNMs.

In direct tests of autistic females and males, we used the same method described above (a binomial test) to compare the fraction of total DNM counts (i.e. total in autistic males and females) that were observed in autistic females with the fraction expected given the fraction of all autistic individuals that were female. For transmission analysis, we calculated the ratio between the total number of parental alleles in autistic females and the total number in both autistic males and females and used this as the expected ratio for a binomial test comparing the transmitted variant counts in autistic females and the total transmitted variants (in both autistic males and females).

We performed these tests separately for each sex in SPARK and ASC and meta-analyzed the rate ratios for the sex-stratified comparisons using the inverse variance-weighted average of the rate ratios. In these exome-wide comparisons, the *p*-values from the binomial test were conservatively corrected for 54 tests using Bonferroni correction from these groups: three sex-stratified comparisons (males, females, sex difference), three cohorts (ASC, SPARK, Meta-analysis of both), three variant classes (synonymous, missense, protein-truncating), and two inheritance models (*de novo*, transmitted). We also used Benjamini-Hochberg False Discovery Rate (FDR) adjustment as Bonferroni correction is conservative given the non-independence of the meta-analyzed estimates. We used stars in the figures to indicate whether the *p*-values were < 0.05 after Bonferroni correction (***), after FDR-adjustment (**), or only before correction (*).

### Variant liability

Assuming that autism liability is additive and normally distributed in the general population, the difference in the average liability in carriers of a certain group of variants and the average in the general population is a measure of the average effect size of these variants, i.e. an estimate of how far this group of variant pushes their carriers (on average) on the liability scale. This variant liability can be estimated from the carrier rates in the study cohorts as detailed previously.^2^ The procedure we used to calculate the estimates is depicted in Figure S4 and detailed in section 5 of the Supplementary Methods (see the Supplementary Note). We used an autism population prevalence estimate of 2.5% in males (1 in 40), with male-to-female prevalence ratio^17^ of 4:1. We took the *p*-values obtained from a binomial test comparing the variant counts in the probands and siblings (outlined above), and estimated the standard errors of the average liability estimates, and subsequently the 95% confidence intervals, from these *p*-values. These calculations were performed separately for each sex and each cohort, and meta-analyzed between cohorts. To directly compare autistic females and males, we calculated the difference between the variant liability estimates obtained separately in female and male probands (Z-score difference). We corrected the *p*-values for multiple testing in a similar manner to the exome-wide enrichment (54 tests). Moreover, we explored whether removing 354 high-confidence and syndromic autosomal genes curated by the Simons Foundation Autism Research Initiative^18^ - hereafter, SFARI genes - would uncover any sex-biased exome-wide variant liability. PTVs in SFARI gene-set were considered damaging if they occurred in 218 SFARI genes that are highly LoF-intolerant (in the most-intolerant LOEUF decile), whereas missense variants were considered damaging if they had an MPC score ≥ 2 (in all 354 genes).

### Autism with co-occurring motor and cognitive difficulties

To explore how genetic architecture differed by phenotype, we split the autistic individuals in the SPARK cohort into those with autism with co-occurring cognitive impairment or motor delay, and those with autism who otherwise had typical cognitive and motor development (see section 1.9 of the Supplementary Methods; the Supplementary Note). Variant counts stratified by these co-occurring conditions were not available from the ASC/SSC cohort for this analysis. SPARK individuals reported to have an Intelligence Quotient (IQ) below 80, cognitive impairment (reported professional diagnosis of an intellectual disability, cognitive impairment, global developmental delay, or borderline intellectual functioning), or motor delay (reported professional diagnosis of delay in walking or developmental coordination disorder) constituted the ‘autism with motor or cognitive impairment group’ (4,209 trio-sequence, 4,714 with one sequenced parent, 1,778 without sequenced parents). We chose the IQ cutoff of 80 since it was previously suggested that defining cognitive impairment in SPARK based on this cutoff minimizes the grouping of average and borderline IQ individuals together, and that a diagnosis of an intellectual disability does not necessarily require an IQ below 70.^19^ Individuals reported not to have any of these co-occurring conditions formed the ‘autism without motor or cognitive impairment’ group (7,420 trio-sequenced, 6,938 with one sequenced parent, 2,141 without sequenced parents), whereas those with missing data on these phenotypes (1,844 trio-sequenced, 1,756 with one sequenced parent, 2,615 without sequenced parents) were considered unclassified (Supplementary Table S4 in section 4 of the Supplementary Methods).

Previous estimates suggested that about one third of autistic individuals in the population have cognitive impairment.^4,5^ Among 11,630 autistic individuals in SPARK who could be classified, ∼ 36% fell in the autism with motor or cognitive impairment group (35% in males & 40% in females). For estimating liability, we scaled the sex-specific population prevalence using these percentages, i.e., using a prevalence estimate of 0.88% in males (35% x 2.5%) and 0.25% in females (40% x 0.625%) for liability calculations in the autism with motor or cognitive impairment group; for the autism without motor or cognitive impairment group, we used a prevalence of 1.62% in males (2.5% - 0.88%) and 0.38% in females (0.625% - 0.25%). Male-specific Z-scores were subtracted from female-specific estimates to assess the sex difference. Last, we estimated the sex-stratified liability of having motor or cognitive impairment among autistic individuals by directly comparing those with motor or cognitive impairment to those without these co-occurring conditions (prevalence of 0.4 in females and 0.35 in males).

### Gene set burden

We evaluated the rare variant burden in high-confidence and syndromic autism-predisposition genes and genes with male-biased or female-biased expression in the fetal cortex^20^ as well as those with sex-biased expression in the adult human cortex.^21^ We examined these gene sets directly and also gauged the extent of the observed enrichment against the burden expected for a similarly-sized gene set selected from the remaining protein-coding genes. For each tested gene set, we selected a random gene set matched for LoF-constraint, brain expression, and coding sequence length distribution (described further in section 6 of the Supplementary Methods, see the Supplementary Note), and counted DNMs, transmitted variants, and untransmitted variants; we repeated this procedure 10,000 times with replacement and took the average ratio (rate ratio between DNM counts in probands and siblings or transmitted to untransmitted ratio in the probands); we then used this ratio as the expected ratio in a binomial test as described above. Specifically, we tested the difference between the rate of DNMs between probands and siblings against the permutation-averaged expected ratio for this gene set (instead of the sample size ratio used in the exome-wide analyses), and we similarly tested rare variant over-transmission against the permutation-averaged expected transmitted-to-untransmitted ratio for the given gene set (instead of 0.5 as used in the exome-wide analysis). We also used the average variant rates across these 10,000 permutations instead of the rate in siblings to estimate the variant liability attributed to a gene set in excess of what is expected for matched genes.

## Results

We examined the burden of rare autosomal *de novo* mutations and inherited variants (minor allele frequency < 0.1%) exome-wide and in specific gene sets in a cohort of 21,501 autistic individuals (13,473 from SPARK; 8,028 from ASC) and 9,223 siblings (6,763 from SPARK; 2,460 from ASC). In these trio-sequenced individuals, the sex-stratified synonymous variant rates (variants per sample) were comparable between the autistic probands and siblings not diagnosed with autism (i.e. rate ratio not significantly different from 1) (Figure S5 in section 1 of the Supplementary Results; see the Supplementary Note), whereas the rates of *de novo* high-confidence PTVs in highly LoF-intolerant genes (hereafter, damaging PTVs) and missense variants with an MPC score ≥ 2 (damaging missense) were higher in autistic probands (Figure 1). Over-transmission was most noticeable in damaging PTVs. Autistic females showed higher carrier rates of damaging variants than autistic males, particularly those occurring *de novo*, albeit to different degrees in SPARK and ASC. Additional analyses of ultra-rare variants in 13,435 autistic individuals with sequence data from one parent and 12,125 autistic individuals without parental sequence data also showed an enrichment in damaging PTVs (Figure S6 in section 1 of the Supplementary Results). These comparisons of DNMs and inherited variants between sexes, which we describe in more detail in section 2 of the Supplementary Results, recapitulated the known sex-differential patterns of enrichment of damaging PTVs and missense variants in these cohorts.^1,12,14^ These comparisons (rate ratios) do not take into account the differences in trait prevalence. Therefore, we next examined whether the sex differences on the observed scale translate into sex differences in liability, which allows comparing effect sizes between groups with different trait prevalences.

**Figure 1:**
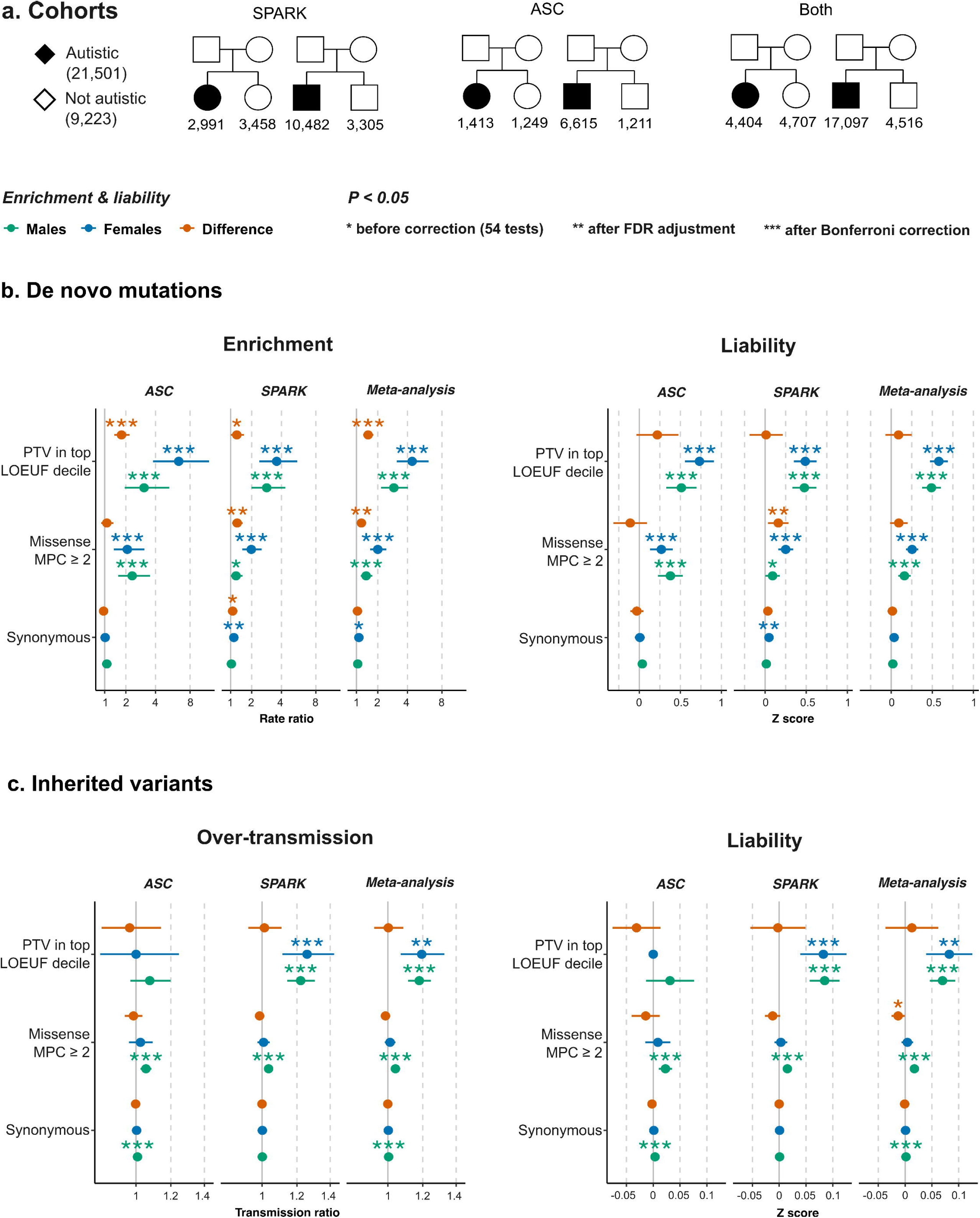
Exome-wide rare variant burden and liability in SPARK and ASC trio-sequenced cohorts. **a**, The sample size of the trio-sequenced individuals in the Simons Foundation Powering Autism Research for Knowledge study (SPARK) and the Autism Sequencing Consortium (ASC) cohorts. **b,** Sex-stratified *de novo* mutation rate ratios (left) and liability (right) (see Methods). For sex differences, a rate ratio > 1 indicates that females show a higher enrichment; a Z score > 0 indicates that females show a higher effect size on the liability scale. **c**, Over-transmission and liability of inherited variants were assessed using similar comparisons between parental alleles transmitted to autistic individuals and untransmitted alleles (see Methods). Error bars show 95% confidence intervals. See Supplementary Table S5 for the results shown here (Extended Tables) and sections 2.1.3 and 2.2.3 of the Supplementary Results (Supplementary Note) for further details on synonymous variant imbalances.

### Sex differences in genetic liability conferred by rare variants exome-wide

Here, we focus on the meta-analyzed (ASC & SPARK) cohort (Figure 1a) with a total sample size of 4,404 autistic females (*versus* 4,707 female siblings) and 17,097 autistic males (*versus* 4,516 male siblings). Despite the relatively higher enrichment (rate ratio) of damaging protein-truncating and missense *de novo* mutations in autistic females compared to males (Figure 1b), we did not find statistically significant differences between the male-derived and female-derived liability estimates.

Specifically, the effect sizes of damaging protein-truncating DNMs on the liability scale in autistic females and in autistic males were not significantly different (Z_Sex-Difference_ = 0.90; 95% CI −0.0684 - 0.25; p = 0.27). The difference between the effect sizes of damaging missense DNMs in females and in males was also not statistically significant (Z_Sex-Difference_ = 0.093; 95% CI = −0.014 - 0.20; p = 0.087). Rare inherited damaging PTVs conveyed significant liabilities in females and in males but these effect sizes were not significantly different between the two sexes (Z_Sex-Difference_ = 0.013; 95% CI = −0.037 - 0.062; p = 0.62). On the other hand, inherited damaging missense variants had higher liability in males compared to females. However, the difference was small and only significant before correcting for 54 multiple tests (Z_Sex-Difference_ = −0.013, 95% CI: −0.00052 - −0.026; p = 0.041; FDR-adjusted p = 0.096; Bonferroni-corrected p = 1). There was a significant imbalance in transmitted and untransmitted synonymous alleles in males (Rate ratio_Males/ASC_ = 1.0081 ; 95% CI = 1.0042 - 1.012; p = 4.1 ×10^-5^; Bonferroni-corrected p = 2.2×10^-3^), although at such a small effect size (Z_Males/ASC_ = 0.0034; 95% CI = 0.0018 - 0.0050) it is unlikely that it affected the main conclusions (see section 2.2.3 of the Supplementary Results).

The sex-stratified cohort-level effect sizes, which we present in more detail in section 2 of the Supplementary Results (see the Supplementary Note), were generally consistent with the meta-analyzed estimates except for inherited damaging PTVs; these conveyed significant liability only in SPARK (Figure 1c). Although the ASC cohort did not show a significant enrichment in inherited damaging PTVs exome-wide, it was significantly enriched for inherited damaging PTVs in known autism predisposition genes (i.e. high-confidence PTVs in 218 highly LoF-intolerant genes amongst 354 SFARI genes; Figure S9 in section 2.2 of the Supplementary Results). Conversely, removing SFARI genes did not change the overall conclusions regarding the sex differences in the average liability of DNM and rare inherited variants.

In terms of cohort-level sex differences in liability, damaging missense DNMs had a higher effect size in females in SPARK (Z_Sex-Difference/SPARK_ = 0.16, 95% CI = 0.034 - 0.28; p = 0.013; FDR-adjusted p = 0.031; Bonferroni-corrected p = 0.70) but this difference was neither significant in the meta-analysis (see above) nor when a subset of ultra-rare DNMs was evaluated (Figure S8 in section 2.1 of the Supplementary Results).

We note that synonymous DNMs in autistic females in SPARK also had a rate ratio that was significantly higher than 1 (Rate ratio_Females/SPARK_ = 1.14; 95% CI = 1.03-1.26; p = 0.012; FDR-adjusted p = 0.029; Bonferroni-corrected p = 0.60), corresponding to an effect size of 0.046 on the liability scale (95% CI = 0.010 - 0.082). This association of synonymous DNMs with autism did not persist when the two cohorts were meta-analyzed (Z_Females/Meta-analysis_ = 0.036; 95% CI = 0.0055 - 0.067; p = 0.052), nor when we examined a group of ultra-rare DNMs. As detailed in section 2.1.4 of the Supplementary Results (see the Supplementary Note), restricting to ultra-rare alleles controlled the spurious signal in synonymous DNMs but did not affect the outcomes of the protein-truncating DNMs (Figure S8).

An exome-wide analysis in the remaining cohorts reiterated the findings from trio analysis (sections 2.2 and 2.3 of the Supplementary Results). In brief, the average effect size of ultra-rare inherited variants in autistic individuals with sequence data from one parent in SPARK did not differ significantly between sexes after correction for multiple testing (Figure S10 in section 2.2 of the Supplementary Results). Similarly, there was no significant sex difference in the average liability conferred by ultra-rare variants in the ASC case-control cohorts and in the autistic individuals without parental sequence data in SPARK (Figure S11 in section 2.3 of the Supplementary Results).

To recapitulate, the average liability conferred by damaging *de novo* protein-truncating and missense mutations as well as inherited PTVs did not show a significant sex difference. Inherited damaging missense variants conferred higher liability in males but this difference was very small in magnitude and only significant before accounting for multiple testing. It was also not seen when examining the transmission of ultra-rare variants in a similarly sized, separate set of autistic individuals from SPARK with sequence data from one parent. Next, we examined whether rare variant liability differs when accounting for co-occurring motor and cognitive difficulties.

### Exome-wide burden in autistic individuals with *versus* without cognitive and motor difficulties

Both SPARK and ASC cohorts included a mixture of autistic individuals with a range of less pronounced and profound motor and cognitive difficulties.^1,14^ To control for any sex differences that may be driven by the differences in the relative frequency of males and females diagnosed with co-occurring cognitive impairment or motor delay, we repeated the exome-wide comparisons in SPARK sub-cohorts stratified by co-occurring cognitive impairment and motor delay (Figure 2a). Contrary to the observed sex differences seen before stratifying by phenotype (Rate Ratio_Sex-Difference_ = 1.52; 95% CI = 1.29 - 1.78; p = 3.3×10^-7^; Figure 1b), rates of damaging *de novo* protein-truncating mutations were not significantly different between autistic females and males with motor and cognitive impairment (Rate Ratio_Sex-Difference_ = 1.12; 95% CI = 0.79 - 1.57; p = 0.49) or autistic females and males without these difficulties (Rate Ratio_Sex-Difference_ = 1.25; 95% CI = 0.84 - 1.83; p = 0.23).

**Figure 2:**
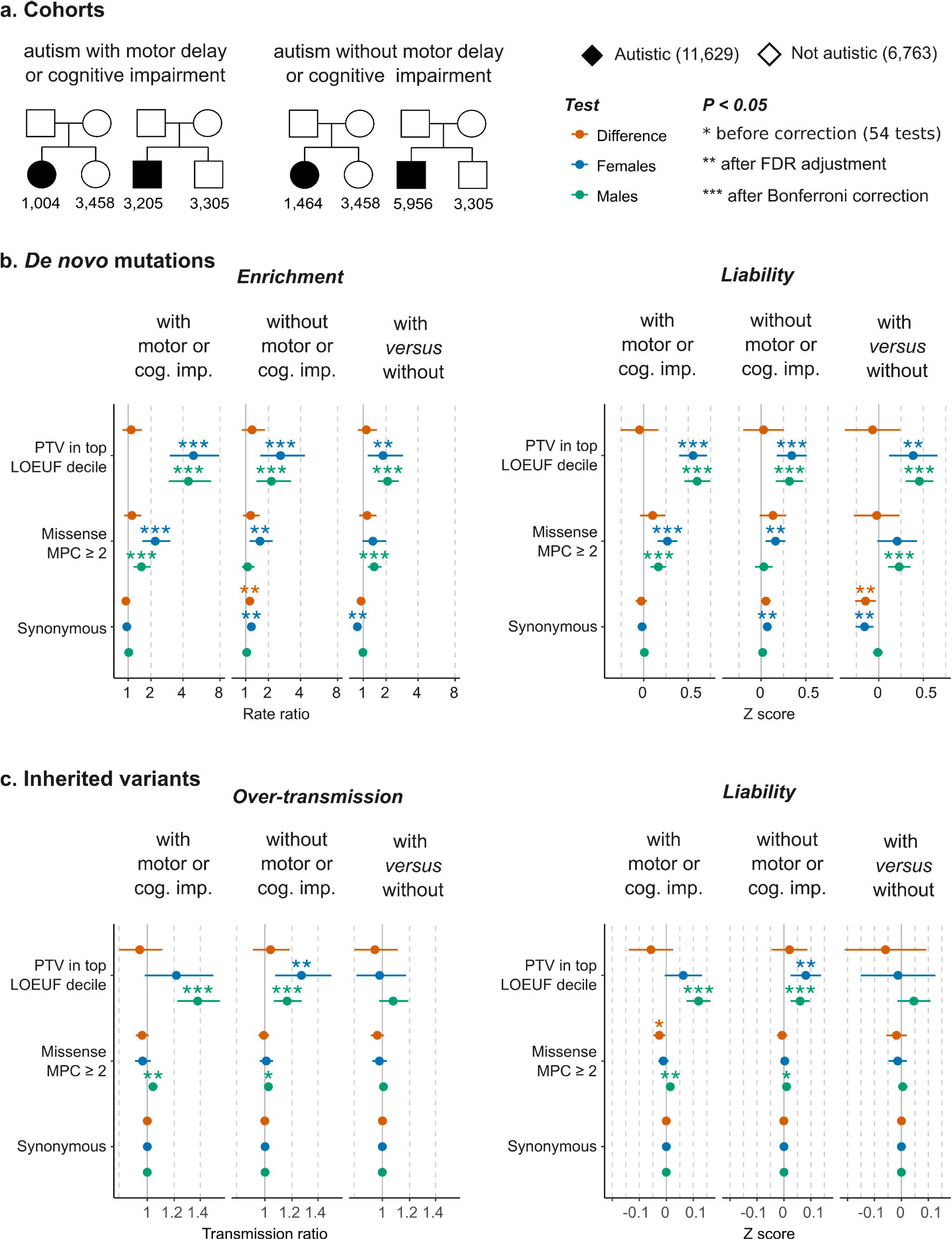
Rare variant burden in autistic individuals with and without cognitive impairment or motor delay in SPARK trio-sequenced cohort. **a**, The sample size in two sub-cohorts of trio-sequenced individuals from the Simons Foundation Powering Autism Research for Knowledge study (SPARK) divided based on the presence of co-occurring motor developmental delays or cognitive impairment. **b**, Sex-stratified observed *de novo* mutation rates (left) and average liability (right) (see Methods). In addition to the comparisons versus siblings, autistic probands with motor or cognitive difficulties were compared directly to sex-matched autistic individuals without these co-occurring conditions (‘with *versus* without’). **c**, Over-transmission of rare inherited variants (left) and their average liability (right) (see Methods). The results depicted in this figure are available in Supplementary Table S6 (see the Extended Tables). See section 4.1.3 of the Supplementary Results (see the Supplementary Note) for details on the imbalance of synonymous variants. Limiting the analysis in **b** to ultra-rare DNMs in ancestry-matched autistic females and siblings showed well-balanced synonymous DNM burden (p = 0.42; Figure S13).

There was no significant sex difference in terms of effect sizes attributed to these protein-truncating *de novo* mutations on the liability scale, neither among autistic individuals with co-occurring cognitive impairment or motor delay (Z_Sex-Difference_ = −0.045; 95% CI = −0.26 - 0.17; p = 0.68), among those without these co-occurring conditions (Z_Sex-Difference_ = −0.069; 95% CI= −0.38 - 0.24; p = 0.66), nor between those with *versus* without cognitive or motor impairment (Z_Sex-Difference_ = 0.026; 95% CI = −0.20 - 0.25; p =0.82). The effect sizes of damaging *de novo* missense mutations and damaging inherited protein-truncating and missense variants were not significantly different between the two sexes after correcting for multiple testing (Figure 2b). These comparisons are detailed in section 4 of the Supplementary Results (Supplementary Note).

### Average liability of rare variants in genes with sex-biased expression in the cortex

In the ASC cohort, which contains a mix of individuals with and without cognitive impairment or motor delay, females had a significantly increased burden of damaging PTVs in 856 genes (including 226 genes in the most-constrained decile) with male-biased expression in the fetal cortex (Figure 3a) that was higher than the burden in males (Rate Ratio_Sex-Difference_ = 2.15; 95% CI = 1.38 - 3.28; p = 7.1×10^-4^; FDR-adjusted p = 0.018; Bonferroni-corrected p = 0.13). However, this did not translate into a sex difference in the liability conferred by these variants (Z_Sex-Difference_ = 0.10; 95% CI = −0.46 - 0.67; p = 0.72; Figure 3b). In SPARK, the sex difference in *de novo* enrichment was less pronounced (Rate Ratio_Sex-Difference_ = 1.72; 95% CI = 1.04 - 2.77; p = 0.024; FDR-adjusted p = 0.2; Bonferroni-corrected p = 1) and not significant even after less conservative correction for multiple testing (FDR-adjusted p = 0.20). Similar to the ASC, there was no significant change in liability (Z_Sex-Difference_ = 0.13; 95% CI = −0.46 - 0.71; p = 0.67).

**Figure 3:**
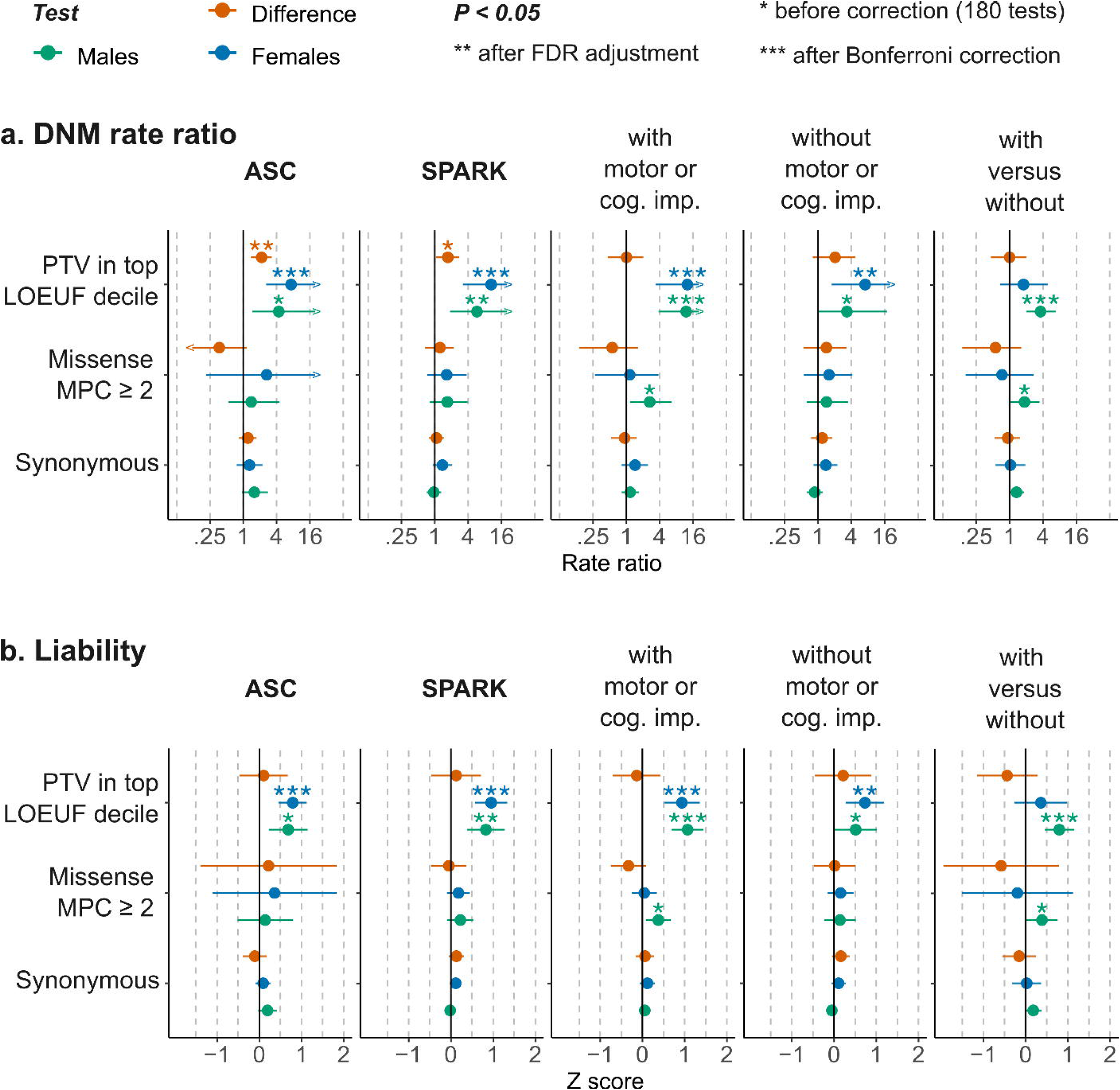
Burden and liability of damaging protein truncating de novo mutations in 856 genes with male-biased expression in the human fetal cortex. **a**, The relative risk (rate ratio) attributed to *de novo* mutations (DNM), examined along with rare inherited variants (not shown), in trio-sequenced individuals from the Autism Sequencing Consortium (ASC) and the Simons Foundation Powering Autism Research for Knowledge (SPARK) cohorts, in two SPARK sub-cohorts of autistic individuals ascertained to have autism with or without co-occurring developmental delay or cognitive impairment (versus siblings), and compared directly between these two groups. **b**, The corresponding average liability attributed to DNMs (effect size on the liability scale). See Methods for details, Figures 1 and 2 for the sample sizes, and Figure S17 in section 6.2 of the Supplementary Methods (Supplementary Note) for a comparison against matched genes. The complete results of DNM and rare inherited variants (180 tests) are presented in Supplementary Table S7 (Extended Tables).

When stratified by motor and cognitive difficulties, autistic individuals with cognitive or motor impairment were significantly enriched for *de novo* protein truncating mutations in genes with male-biased expression in the fetal cortex, with effect sizes that did not differ significantly between males and females (Z_Sex-Difference_ = −0.031; 95% CI = −0.7 - 0.43; p = 0.64). Among autistic individuals without cognitive or motor impairment, females but not males showed significant burden in this gene set, albeit only after less conservative correction for multiple testing (FDR-adjusted *p*-value in females = 0.034); the two sex-stratified estimates were not significantly different (Z_Sex-Difference_ = 0.22; 95% CI = −0.54 - 0.89; p = 0.53).

Within these genes with male-biased expression in the fetal cortex, damaging missense DNMs as well as damaging inherited protein-truncating and missense variants were not significantly enriched in autistic individuals after correction for multiple testing (Figure 3). In none of the sub-cohorts or variant classes examined was the sex-stratified burden significantly higher (after multiple testing correction) than what is expected for a gene set matched for constraint and other metrics (Figure S17 in section 6.2 of the Supplementary Results, see the Supplementary Note). Genes with female-biased expression in the fetal cortex (n=794; including 52 genes in the most-constrained decile) did not show any significant enrichment in damaging *de novo* protein-truncating mutations (Supplementary Table S18; see Extended Tables). Also, genes that show male-biased (n=303; 32 genes in the most-constrained decile) or female-biased (n=426; 61 genes in the most-constrained decile) expression in adult cortical tissues were not substantially enriched for damaging protein-truncating DNMs (Supplementary Tables S19 and S20; Extended Tables).

## Discussion

We examined the sex differences in rare autosomal coding variant rates both exome-wide and in specific gene sets in 47,061 autistic individuals from two large autism cohorts, showing that the average liability attributed to damaging rare variants exome-wide and in genes with sex-biased expression in fetal or adult cortex is not statistically significantly different between males and females.

The sex differences in rare variant enrichment on the observed scale are likely caused by the differences in the prevalence of autism and co-occurring cognitive impairment in males and females, as these did not translate into differences in variant liability between sexes (Figure 1) and were driven by autism-predisposition genes known to increase the chance of other developmental disorders affecting motor and cognitive skills (Figure S7 in section 2.1 of the Supplementary Results). Indeed, comparisons stratified by these co-occurring conditions in SPARK did not show significant sex differences in exome-wide burden and liability, but did confirm that carriers of protein-truncating *de novo* mutations in haploinsufficient genes have an increased likelihood of motor and cognitive impairment (Figure 2). Therefore, the sex differences in damaging DNM rate ratios seen when examining a mix of individuals with and without motor and cognitive difficulties may reflect a difference in the relative frequency of these endo-phenotypes between the sexes in the study cohort. This, in turn, is likely driven by there being a higher likelihood of co-occurring cognitive impairment amongst autistic females than autistic males.

Genes with male-biased expression in fetal brain were enriched for damaging DNMs in autistic individuals compared to controls but did not show sex differences in rare variant liability after controlling for multiple testing (Figure 3). Unlike known autism-predisposition genes, which have a significantly higher DNM burden (on the observed scale) compared to matched genes with comparable constraint and brain expression (Figure S16 in section 6.1 of the Supplementary Results), the enrichment in these sex-biased gene sets is largely attributed to their overlap with LoF-intolerant genes and genes with high expression in brain (Figure S17 in section 6.2 of the Supplementary Results).

Protein-disrupting alterations confer the highest predisposition for autism among rare short coding variants.^1,12,14^ Since genes implicated through *de novo* association are generally developmental disorder genes (e.g., SFARI high-confidence and syndromic genes), it is unclear whether they increase the predisposition for autism *per se*. Under a Liability Threshold Model, in which autism predisposition is assumed to be additive and normally distributed (Figure S4 in section 4 of the Supplementary Methods), an autism prevalence of 2.5% in the general population puts the threshold for autism diagnosis at 1.96 standardized units. We estimate that protein-truncating *de novo* mutations (in highly LoF-constrained genes) increase the chance of having autism without co-occurring motor or cognitive impairment by 0.32 units in males (95% CI = 0.16 - 0.47) and 0.34 units in females (95% CI = 0.17 - 0.51), which alone is not sufficient to reach the threshold for autism diagnosis in the absence of other factors such as, for example, a high polygenic score for autism. We have previously shown that the association between autism diagnosis and an autism polygenic score capturing a proportion of the predisposition from common variants is more pronounced in autistic individuals with few motor and cognitive developmental difficulties than in those with several developmental disabilities.^22^

On the other hand, the stratified analysis we carried out in SPARK suggests that these damaging protein-truncating *de novo* mutations have an average effect size that conveys enough risk to cause co-occurring motor or cognitive difficulties in autistic individuals. About a third of autistic individuals in the population^4^ and 35% (males) to 40% (females) in SPARK have autism with motor or cognitive impairment. Assuming that the genetic predisposition for these co-occurring conditions amongst the individuals already diagnosed with autism is also additive and normally distributed, the threshold needed so that one third of autistic individuals would have motor or cognitive difficulties is ∼0.4 units above the mean of those without co-occurring conditions. The observed two-fold enrichment of protein-truncating *de novo* mutations in autistic individuals with *versus* without motor or cognitive impairment translates to 0.39 units in liability in females (95% CI = 0.11 - 0.66) and 0.45 units in males (95% CI = 0.30 - 0.66), which is sufficient to reach this threshold. Taken together with the findings in those without co-occurring conditions, autism is likely an incompletely penetrant, complex phenotype within the phenotypic spectrum of these developmental genes.

A strength of this study is that we included samples from diverse populations. In theory, since our core analyses were focused on within-family analyses of *de novo* mutations and inherited variants, the inclusion of individuals of different ancestries should not create spurious stratification effects. However, in practice this resulted in a subtle association of synonymous DNMs with autism diagnosis in SPARK (Figure 1a), discussed in sections 2.1.3 and 4.1.3 of the Supplementary Results. The imbalance in synonymous DNMs was most prominent between autistic females without motor delay or cognitive impairment and sex-matched siblings not diagnosed with autism (Figure 2b). However, it is unlikely that this subtle imbalance biased the outcomes for damaging protein-truncating DNMs.

Our analysis relied on a standard Liability Threshold Model assuming equal variance of the liability distribution in males and females.^9,10^ This model is easily interpretable and allows direct comparisons between the two sexes on the same scale. Alternative models with less restrictive assumptions, e.g., higher variance in males,^4,23^ may better capture the true underlying distribution of autism predisposition in the population. Another limitation of our study is that we only examined rare variants on the autosomes, ignoring the sex chromosomes. We note that recent large-scale gene association studies from the ASC did not include sex chromosomes,^1,2^ so the contribution of sex-linked genes may be underestimated. Nonetheless, it seems unlikely that large-effect rare variants on the sex chromosomes are a major driver of the sex difference in autism, at least amongst those with co-occurring motor and cognitive impairments, since our previous work in the Deciphering Developmental Disorders cohort found that rare Mendelian-acting coding variants in the X chromosome contributed similarly in males and females and didn’t explain the observed 1.6:1 male-bias.^24^

To summarize, deleterious *de novo* and rare inherited autosomal coding variants confer similar liability for autism in females and males. These variants, particularly *de novo* protein-truncating mutations, increase the liability for co-occurring motor or cognitive impairment significantly more so than autism with otherwise typical motor and cognitive development. Autosomal *de novo* mutations with large effect sizes are therefore unlikely to explain the observed sex differences in autism prevalence. Future studies with larger sample sizes, considering the contribution of both autosomal and sex-linked alleles across the frequency spectrum of rare and common variants, may capture additional predisposing variants with small effect sizes that contribute to the sex differences in autism.

## Data Availability

The data analyzed in this work were obtained from the Simons Foundation Autism Research Initiative (SFARI) and the Autism Sequencing Consortium (ASC). The Simons Foundation Powering Autism Research for Knowledge study (SPARK) phenotypes and exome data are available for approved users through SFARI Base (https://www.sfari.org/resource/sfari-base/). The ASC data used in this study are available for approved users at NHGRI AnVIL (https://anvilproject.org/data) with the accession ID: phs000298. The data produced in the present work are contained in the manuscript and the Supplementary Information.

## Supplementary Information

SPARK phenotypes and exome data are available for approved users through SFARI Base (https://www.sfari.org/resource/sfari-base/). The ASC data used in this study are available for approved users at NHGRI AnVIL (https://anvilproject.org/data) with the accession ID: phs000298. The Supplementary Note contains the affiliations of the ASC and APEX consortia members, Supplementary Methods (including Supplementary Tables S1-S4 and figures S1-S4), Supplementary Results, and related figures (Figure S5-S17). Enrichment analysis results including the input used to create the main and supplementary figures are available as Extended Tables.

## Acknowledgements

We thank the participants and investigators who contributed to the datasets of the Simons Powering Autism Research for Knowledge project, the Autism Sequencing Consortium, the Simons Simplex Collection, and the Lundbeck Foundation Initiative for Integrative Psychiatric Research (iPSYCH) project. This work was supported by the Simons Foundation Autism Research Initiative (SFARI) through grant RNAG/669 G10 9280 to H.M., V.W. and other principal investigators of the Autism Prenatal Sex Differences Consortium (APEX). The Autism Sequencing Consortium (ASC) received support from: SFARI (574598 to S.J.S., 575097 to B.D. and K.R., 573206 to M.E.T. and M.J.D., 571009 to J.D., 736613 and 647371 to S.J.S., 606362 and 608540 to M.E.T., M.J.D., J.D.B., B.D., K.R. and S.J.S.), NHGRI (HG008895 to M.J.D., S.G. and M.E.T.), NIMH (MH115957 and MH123155 to M.E.T., MH111658 and MH057881 to B.D., MH097849, MH111661 and MH100233 to J.D.B., MH109900 and MH123184 to K.R., MH111660 and MH129722 to M.J.D., MH111662 and MH100027 to S.J.S.), NICHD (HD081256 and HD096326 to M.E.T.), AMED (JP21WM0425007 to N.O.), and the Beatrice and Samuel Seaver Foundation.

## Authors Information

### Autism Sequencing Consortium

Branko Aleksic, Mykyta Artomov, Mafalda Barbosa, Elisa Benetti, Catalina Betancur, Monica Biscaldi-Schafer, Anders D. Børglum, Harrison Brand, Alfredo Brusco, Joseph D. Buxbaum, Gabriele Campos, Simona Cardaropoli, Diana Carli, Angel Carracedo, Marcus C. Y. Chan, Andreas G. Chiocchetti, Brian H. Y. Chung, Brett Collins, Ryan L. Collins, Edwin H. Cook, Hilary Coon, Claudia I. S. Costa, Michael L. Cuccaro, David J. Cutler, Mark J. Daly, Silvia De Rubeis, Bernie Devlin, Ryan N. Doan, Enrico Domenici, Shan Dong, Chiara Fallerini, Montserrat Fernández-Prieto, Giovanni Battista Ferrero, Christine M. Freitag, Jack M. Fu, J. Jay Gargus, Sherif Gerges, Elisa Giorgio, Ana Cristina Girardi, Stephen Guter, Emily Hansen-Kiss, Gail E. Herman, Irva Hertz-Picciotto, David M. Hougaard, Christina M. Hultman, Suma Jacob, Miia Kaartinen, Lambertus Klei, Alexander Kolevzon, Itaru Kushima, So Lun Lee, Terho Lehtimäki, Lindsay Liang, Carla Lintas, Alicia Ljungdahl, Caterina Lo Rizzo, Yunin Ludena, Patricia Maciel, Behrang Mahjani, Nell Maltman, Marianna Manara, Dara S. Manoach, Gal Meiri, Idan Menashe, Judith Miller, Nancy Minshew, Matthew Mosconi, Rachel Nguyen, Norio Ozaki, Aarno Palotie, Mara Parellada, Maria Rita Passos-Bueno, Lisa Pavinato, Minshi Peng, Margaret Pericak-Vance, Antonio M. Persico, Isaac N. Pessah, Kaija Puura, Abraham Reichenberg, Alessandra Renieri, Kathryn Roeder, Stephan J. Sanders, Sven Sandin, F. Kyle Satterstrom, Stephen W. Scherer, Sabine Schlitt, Rebecca J. Schmidt, Lauren Schmitt, Katja Schneider-Momm, Paige M. Siper, Laura Sloofman, Moyra Smith, Christine R. Stevens, Pål Suren, James S. Sutcliffe, John A. Sweeney, Michael E. Talkowski, Flora Tassone, Karoline Teufel, Elisabetta Trabetti, Slavica Trajkova, Maria del Pilar Trelles, Brie Wamsley, Jaqueline Y. T. Wang, Lauren A. Weiss, Mullin H. C. Yu and Ryan Yuen.

### APEX Consortium

Deep Adhya, Carrie Allison, Bonnie Ayeung, Rosie Bamford, Simon Baron-Cohen, Richard Bethlehem, Tal Biron-Shental, Graham Burton, Wendy Cowell, Jonathan Davies, Dori Floris, Alice Franklin, Lidia Gabis, Daniel Geschwind, David M. Greenberg, Yuanjun Gu, Alexandra Havdahl, Alexander Heazell, Rosemary Holt, Matthew Hurles, Yumnah Khan, Meng-Chuan Lai, Madeline Lancaster, Michael Lombardo, Hilary Martin, Jose Gonzalez Martinez, Jonathan Mill, Mahmoud K. Musa, Kathy Niakan, Adam Pavlinek, Lucia Dutan Polit, Marcin Radecki, David Rowitch, Laura Sichlinger, Deepak Srivastava, Alexandros Tsompanidis, Florina Uzefovsky, Varun Warrier, Elizabeth Weir, Xinhe Zhang.

## Author contributions

Study design: HM, VW, MK, APEX Consortium. Samples and data generation: Autism Sequencing Consortium. Quality control and data preparation: MK and FKS. Analysis: MK. Writing: MK, HM, VW. Study direction and supervision: HM, VW.

## References

1. Fu, J. M. et al. Rare coding variation provides insight into the genetic architecture and phenotypic context of autism. Nat. Genet. 54, 1320–1331 (2022).

2. Satterstrom, F. K. et al. Large-Scale Exome Sequencing Study Implicates Both Developmental and Functional Changes in the Neurobiology of Autism. Cell 180, 568–584.e23 (2020).

3. De Rubeis, S. et al. Synaptic, transcriptional and chromatin genes disrupted in autism. Nature 515, 209–215 (2014).

4. Dougherty, J. D. et al. Can the “female protective effect” liability threshold model explain sex differences in autism spectrum disorder? Neuron 110, 3243–3262 (2022).

5. Zeidan, J. et al. Global prevalence of autism: A systematic review update. Autism Res. 15, 778–790 (2022).

6. Chaste, P., Roeder, K. & Devlin, B. The Yin and Yang of Autism Genetics: How Rare De Novo and Common Variations Affect Liability. Annu. Rev. Genomics Hum. Genet. 18, 167–187 (2017).

7. Cirnigliaro, M. et al. The contributions of rare inherited and polygenic risk to ASD in multiplex families. Proc. Natl. Acad. Sci. 120, e2215632120 (2023).

8. Klei, L. et al. How rare and common risk variation jointly affect liability for autism spectrum disorder. Mol. Autism 12, 66 (2021).

9. Carter, C. O. THE INHERITANCE OF CONGENITAL PYLORIC STENOSIS. Br. Med. Bull. 17, 251–253 (1961).

10. Falconer, D. S. The inheritance of liability to certain diseases, estimated from the incidence among relatives. Ann. Hum. Genet. 29, 51–76 (1965).

11. Zhu, C. et al. Amplification is the primary mode of gene-by-sex interaction in complex human traits. Cell Genomics 3, 100297 (2023).

12. Antaki, D. et al. A phenotypic spectrum of autism is attributable to the combined effects of rare variants, polygenic risk and sex. Nat. Genet. 54, 1284–1292 (2022).

13. Chan, A. J. S. et al. Genome-wide rare variant score associates with morphological subtypes of autism spectrum disorder. Nat. Commun. 13, 6463 (2022).

14. Zhou, X. et al. Integrating de novo and inherited variants in 42,607 autism cases identifies mutations in new moderate-risk genes. Nat. Genet. 54, 1305–1319 (2022).

15. Loomes, R., Hull, L. & Mandy, W. P. L. What Is the Male-to-Female Ratio in Autism Spectrum Disorder? A Systematic Review and Meta-Analysis. J. Am. Acad. Child Adolesc. Psychiatry 56, 466–474 (2017).

16. Ratto, A. B. et al. What About the Girls? Sex-Based Differences in Autistic Traits and Adaptive Skills. J. Autism Dev. Disord. 48, 1698–1711 (2018).

17. Maenner, M. J. et al. Prevalence and Characteristics of Autism Spectrum Disorder Among Children Aged 8 Years — Autism and Developmental Disabilities Monitoring Network, 11 Sites, United States, 2018. MMWR Surveill. Summ. 70, 1–16 (2021).

18. Abrahams, B. S. et al. SFARI Gene 2.0: a community-driven knowledgebase for the autism spectrum disorders (ASDs). Mol. Autism 4, 36 (2013).

19. Shu, C., Green Snyder, L., Shen, Y., Chung, W. K., & SPARK Consortium. Imputing cognitive impairment in SPARK, a large autism cohort. Autism Res. Off. J. Int. Soc. Autism Res. 15, 156–170 (2022).

20. O’Brien, H. E. et al. Sex Differences in Gene Expression in the Human Fetal Brain. http://biorxiv.org/lookup/doi/10.1101/483636 (2018) doi:10.1101/483636.

21. Fass, S. B. et al. Relationship between Sex Biases in Gene Expression and Sex Biases in Autism and Alzheimer’s Disease. http://medrxiv.org/lookup/doi/10.1101/2023.08.29.23294773 (2023) doi:10.1101/2023.08.29.23294773.

22. Warrier, V. et al. Genetic correlates of phenotypic heterogeneity in autism. Nat. Genet. 54, 1293–1304 (2022).

23. Falconer, D. S. The inheritance of liability to diseases with variable age of onset, with particular reference to diabetes mellitus. Ann. Hum. Genet. 31, 1–20 (1967).

24. Martin, H. C. et al. The contribution of X-linked coding variation to severe developmental disorders. Nat. Commun. 12, 627 (2021).

