## Supplementary Note for "Contribution of autosomal rare and *de novo* variants to sex differences in autism"

#### Consortia and affiliations

***Autism Sequencing Consortium:***

Branko Alexie: Department of Psychiatry, Graduate School of Medicine, Nagoya University, Nagoya, Japan

Mykyta Artomov: Center for Genomic Medicine, Massachusetts General Hospital, Boston, MA, USA; Program in Medical and Population Genetics, Broad Institute of MIT and Harvard, Cambridge, MA, USA; Stanley Center for Psychiatric Research, Broad Institute of MIT and Harvard, Cambridge, MA, USA; Harvard Medical School, Boston, MA, USA

Mafalda Barbosa: The Mindich Child Health and Development Institute, Icahn School of Medicine at Mount Sinai, New York, NY, USA; Department of Genetics and Genomic Sciences, Icahn School of Medicine at Mount Sinai, New York, NY, USA

Elisa Benetti: Med Biotech Hub and Competence Center, Department of Medical Biotechnologies, University of Siena, Siena, Italy; Medical Genetics, , University of Siena, Siena, Italy

Catalina Betancur: Sorbonne Université, INSERM, CNRS, Neuroscience Paris Seine, Institut de Biologie Paris Seine, Paris, France

Monica Biscaldi-Schafer: Department of Child and Adolescent Psychiatry, Psychosomatics and Psychotherapy, Goethe University Frankfurt, Frankfurt, Germany

Anders D. Børglum: The Lundbeck Foundation Initiative for Integrative Psychiatric Research, iPSYCH, Aarhus, Denmark; Department of Biomedicine—Human Genetics, Aarhus University, Aarhus, Denmark; Center for Genomics and Personalized Medicine, Aarhus, Denmark; Bioinformatics Research Centre, Aarhus University, Aarhus, Denmark

Harrison Brand: Center for Genomic Medicine, Massachusetts General Hospital, Boston, MA, USA; Program in Medical and Population Genetics, Broad Institute of MIT and Harvard, Cambridge, MA, USA; Department of Neurology, Massachusetts General Hospital and Harvard Medical School, Boston, MA,

USA; Pediatric Surgical Research Laboratories, Department of Surgery, Massachusetts General Hospital, Boston, MA, USA

Alfredo Brusco: Department of Medical Sciences, University of Torino, Turin, Italy; Medical Genetics Unit, 'Città della Salute e della Scienza' University Hospital, Turin, Italy

Joseph D. Buxbaum: Seaver Autism Center for Research and Treatment, Icahn School of Medicine at Mount Sinai, New York, NY, USA; Department of Psychiatry, Icahn School of Medicine at Mount Sinai, New York, NY, USA; The Mindich Child Health and Development Institute, Icahn School of Medicine at Mount Sinai, New York, NY, USA; Department of Genetics and Genomic Sciences, Icahn School of Medicine at Mount Sinai, New York, NY, USA; Friedman Brain Institute, Icahn School of Medicine at Mount Sinai, New York, NY, USA; Department of Neuroscience, Icahn School of Medicine at Mount Sinai, New York, NY, USA

Gabriele Campos: Centro de Pesquisas sobre o Genoma Humano e Células tronco, Instituto de Biociências, Universidade de São Paulo, São Paulo, Brazil

Simona Cardaropoli: Department of Public Health and Pediatrics, University of Torino, Turin, Italy

Diana Carli: Department of Public Health and Pediatrics, University of Torino, Turin, Italy

Angel Carracedo: Grupo de Medicina Xenómica, Centro de Investigación en Red de Enfermedades Raras (CIBERER), CIMUS, Universidade de Santiago de Compostela, Santiago de Compostela, Spain; Fundación Pública Galega de Medicina Xenómica, Servicio Galego de Saúde (SERGAS), Santiago de Compostela, Spain

Marcus C. Y. Chan: Department of Pediatrics and Adolescent Medicine, Duchess of Kent Children's Hospital, The University of Hong Kong, Hong Kong Special Administrative Region, China

Andreas G. Chiocchetti: Department of Psychiatry, Graduate School of Medicine, Nagoya University, Nagoya, Japan

Brian H. Y. Chung: Department of Pediatrics and Adolescent Medicine, Duchess of Kent Children's Hospital, The University of Hong Kong, Hong Kong Special Administrative Region, China

Brett Collins: Seaver Autism Center for Research and Treatment, Icahn School of Medicine at Mount Sinai, New York, NY, USA; Department of Psychiatry, Icahn School of Medicine at Mount Sinai, New York, NY, USA; The Mindich Child Health and Development Institute, Icahn School of Medicine at Mount Sinai, New York, NY, USA

Ryan L. Collins: Center for Genomic Medicine, Massachusetts General Hospital, Boston, MA, USA; Program in Medical and Population Genetics, Broad Institute of MIT and Harvard, Cambridge, MA, USA; Department of Neurology, Massachusetts General Hospital and Harvard Medical School, Boston, MA, USA; Program in Bioinformatics and Integrative Genomics, Harvard Medical School, Boston, MA, USA

Edwin H. Cook: Institute for Juvenile Research, Department of Psychiatry, University of Illinois at Chicago, Chicago, IL, USA

Hilary Coon: Department of Internal Medicine, University of Utah, Salt Lake City, UT, USA; Department of Psychiatry, Huntsman Mental Health Institute, University of Utah, Salt Lake City, UT, USA

Claudia I. S. Costa: Centro de Pesquisas sobre o Genoma Humano e Células tronco, Instituto de Biociências, Universidade de São Paulo, São Paulo, Brazil

Michael L. Cuccaro: The John P Hussman Institute for Human Genomics, The University of Miami Miller School of Medicine, Miami, FL, USA

David J. Cutler: Department of Human Genetics, Emory University School of Medicine, Atlanta, GA, USA

Mark J. Daly: Center for Genomic Medicine, Massachusetts General Hospital, Boston, MA, USA; Program in Medical and Population Genetics, Broad Institute of MIT and Harvard, Cambridge, MA, USA; Stanley Center for Psychiatric Research, Broad Institute of MIT and Harvard, Cambridge, MA, USA; Analytic and Translational Genetics Unit, Department of Medicine, Massachusetts General Hospital, Boston, MA, USA; Harvard Medical School, Boston, MA, USA; Institute for Molecular Medicine Finland (FIMM), University of Helsinki, Helsinki, Finland

Silvia De Rubeis: Seaver Autism Center for Research and Treatment, Icahn School of Medicine at Mount Sinai, New York, NY, USA; Department of Psychiatry, Icahn School of Medicine at Mount Sinai, New York, NY, USA; The Mindich Child Health and Development Institute, Icahn School of Medicine at Mount Sinai, New York, NY, USA; Friedman Brain Institute, Icahn School of Medicine at Mount Sinai, New York, NY, USA

Bernie Devlin: Department of Psychiatry, University of Pittsburgh School of Medicine, Pittsburgh, PA, USA

Ryan N. Doan: Division of Genetics and Genomics, Boston Children’s Hospital, Boston, MA, USA

Enrico Domenici: Department of Cellular, Computational and Integrative Biology, , University of Trento, Trento, Italy

Shan Dong: Department of Psychiatry, UCSF Weill Institute for Neurosciences, University of California San Francisco, San Francisco, CA, USA

Chiara Fallerini: Med Biotech Hub and Competence Center, Department of Medical Biotechnologies, University of Siena, Siena, Italy; Medical Genetics, , University of Siena, Siena, Italy

Montserrat Fernández-Prieto: Grupo de Medicina Xenómica, Centro de Investigación en Red de Enfermedades Raras (CIBERER), CIMUS, Universidade de Santiago de Compostela, Santiago de Compostela, Spain; Neurogenetics group, Instituto de Investigación Sanitaria de Santiago (IDIS-SERGAS), Santiago de Compostela, Spain

Giovanni Battista Ferrero: Department of Public Health and Pediatrics, University of Torino, Turin, Italy

Christine M. Freitag: Department of Psychiatry, Graduate School of Medicine, Nagoya University, Nagoya, Japan

Jack M. Fu: Center for Genomic Medicine, Massachusetts General Hospital, Boston, MA, USA; Program in Medical and Population Genetics, Broad Institute of MIT and Harvard, Cambridge, MA, USA; Department of Neurology, Massachusetts General Hospital and Harvard Medical School, Boston, MA, USA

J. Jay Gargus: Center for Autism Research and Translation, University of California Irvine, Irvine, CA, USA

Sherif Gerges: Center for Genomic Medicine, Massachusetts General Hospital, Boston, MA, USA; Program in Medical and Population Genetics, Broad Institute of MIT and Harvard, Cambridge, MA, USA; Stanley Center for Psychiatric Research, Broad Institute of MIT and Harvard, Cambridge, MA, USA; Harvard Medical School, Boston, MA, USA

Elisa Giorgio: Department of Medical Sciences, University of Torino, Turin, Italy

Ana Cristina Girardi: Centro de Pesquisas sobre o Genoma Humano e Células tronco, Instituto de Biociências, Universidade de São Paulo, São Paulo, Brazil

Stephen Guter: Institute for Juvenile Research, Department of Psychiatry, University of Illinois at Chicago, Chicago, IL, USA

Emily Hansen-Kiss: Department of Diagnostic and Biomedical Sciences, University of Texas Health Science Center at Houston, School of Dentistry, Houston, TX, USA

Gail E. Herman: The Research Institute at Nationwide Children’s Hospital, Columbus, OH, USA

Irva Hertz-Picciotto: MIND (Medical Investigation of Neurodevelopmental Disorders) Institute, University of California Davis, Davis, CA, USA

David M. Hougaard: Department of Child and Adolescent Psychiatry, Psychosomatics and Psychotherapy, Goethe University Frankfurt, Frankfurt, Germany; Center for Neonatal Screening, Department for Congenital Disorders, Statens Serum Institut, Copenhagen, Denmark

Christina M. Hultman: Department of Medical Epidemiology and Biostatistics, Karolinska Institutet, Stockholm, Sweden

Suma Jacob: Institute for Juvenile Research, Department of Psychiatry, University of Illinois at Chicago, Chicago, IL, USA

Miia Kaartinen: Department of Child Psychiatry, Tampere University and Tampere University Hospital, Tampere, Finland

Lambertus Klei: Department of Psychiatry, University of Pittsburgh School of Medicine, Pittsburgh, PA, USA

Alexander Kolevzon: Seaver Autism Center for Research and Treatment, Icahn School of Medicine at Mount Sinai, New York, NY, USA; Department of Psychiatry, Icahn School of Medicine at Mount Sinai, New York, NY, USA; Department of Pediatrics, Icahn School of Medicine at Mount Sinai, New York, NY, USA

Itaru Kushima: The Lundbeck Foundation Initiative for Integrative Psychiatric Research, iPSYCH, Aarhus, Denmark; Medical Genomics Center, Nagoya University Hospital, Nagoya, Japan

So Lun Lee: Department of Pediatrics and Adolescent Medicine, Duchess of Kent Children’s Hospital, The University of Hong Kong, Hong Kong Special Administrative Region, China

Terho Lehtimäki: Department of Clinical Chemistry, Fimlab Laboratories and Finnish Cardiovascular Research Center-Tampere, Faculty of Medicine and Health Technology, Tampere University, Tampere, Finland

Lindsay Liang: Department of Psychiatry, UCSF Weill Institute for Neurosciences, University of California San Francisco, San Francisco, CA, USA

Carla Lintas: Service for Neurodevelopmental Disorders, University Campus Bio-medico of Rome, Rome, Italy

Alicia Ljungdahl: Department of Psychiatry, UCSF Weill Institute for Neurosciences, University of California San Francisco, San Francisco, CA, USA

Caterina Lo Rizzo: Medical Genetics, , University of Siena, Siena, Italy; Genetica Medica, Azienda Ospedaliera Universitaria Senese, Siena, Italy

Yunin Ludena: MIND (Medical Investigation of Neurodevelopmental Disorders) Institute, University of California Davis, Davis, CA, USA

Patricia Maciel: Life and Health Sciences Research Institute, School of Medicine, University of Minho, Braga, Portugal

Behrang Mahjani: Seaver Autism Center for Research and Treatment, Icahn School of Medicine at Mount Sinai, New York, NY, USA; Department of Psychiatry, Icahn School of Medicine at Mount Sinai, New York, NY, USA; Department of Medical Epidemiology and Biostatistics, Karolinska Institutet, Stockholm, Sweden

Nell Maltman: Institute for Juvenile Research, Department of Psychiatry, University of Illinois at Chicago, Chicago, IL, USA

Marianna Manara: Medical Genetics, , University of Siena, Siena, Italy; Genetica Medica, Azienda Ospedaliera Universitaria Senese, Siena, Italy

Dara S. Manoach: Department of Psychiatry, Massachusetts General Hospital and Harvard Medical School, Boston, MA, USA

Gal Meiri: The Azrieli National Center for Autism and Neurodevelopment Research, Ben-Gurion University of the Negev, Beer-Sheva, Israel; Pre-School Psychiatry Unit, Soroka University Medical Center, Beer Sheva, Israel

Idan Menashe: Department of Public Health, Ben-Gurion University of the Negev, Beer-Sheva, Israel; National Autism Research Center of Israel, Ben-Gurion University of the Negev, Beer-Sheva, Israel

Judith Miller: Children’s Hospital of Philadelphia, Philadelphia, PA, USA; Department of Psychiatry, University of Utah, Salt Lake City, UT, USA

Nancy Minshew: Department of Psychiatry, University of Pittsburgh School of Medicine, Pittsburgh, PA, USA

Matthew Mosconi: Life Span Institute and Kansas Center for Autism Research and Training, University of Kansas, Lawrence, KS, USA

Rachel Nguyen: Center for Autism Research and Translation, University of California Irvine, Irvine, CA, USA

Norio Ozaki: The Lundbeck Foundation Initiative for Integrative Psychiatric Research, iPSYCH, Aarhus, Denmark; Institute for Glyco-core Research (iGCORE), Nagoya University, Nagoya, Japan

Aarno Palotie: Program in Medical and Population Genetics, Broad Institute of MIT and Harvard, Cambridge, MA, USA; Analytic and Translational Genetics Unit, Department of Medicine, Massachusetts General Hospital, Boston, MA, USA; Institute for Molecular Medicine Finland (FIMM), University of Helsinki, Helsinki, Finland; Psychiatric & Neurodevelopmental Genetics Unit, Department of Psychiatry, Massachusetts General Hospital, Boston, MA, USA

Mara Parellada: Department of Child and Adolescent Psychiatry, Hospital General Universitario Gregorio Marañón, IiSGM, CIBERSAM, School of Medicine Complutense University, Madrid, Spain

Maria Rita Passos-Bueno: Centro de Pesquisas sobre o Genoma Humano e Células tronco, Instituto de Biociências, Universidade de São Paulo, São Paulo, Brazil

Lisa Pavinato: Department of Medical Sciences, University of Torino, Turin, Italy

Minshi Peng: Department of Statistics and Data Science, Carnegie Mellon University, Pittsburgh, PA, USA;

Margaret Pericak-Vance: The John P Hussman Institute for Human Genomics, The University of Miami Miller School of Medicine, Miami, FL, USA

Antonio M. Persico: Interdepartmental Program 'Autism 0-90', 'Gaetano Martino' University Hospital, University of Messina, Messina, Italy

Isaac N. Pessah: MIND (Medical Investigation of Neurodevelopmental Disorders) Institute, University of California Davis, Davis, CA, USA; Department of Molecular Biosciences, University of California Davis, School of Veterinary Medicine, Davis, CA, USA

Kaija Puura: Department of Child Psychiatry, Tampere University and Tampere University Hospital, Tampere, Finland

Abraham Reichenberg: Seaver Autism Center for Research and Treatment, Icahn School of Medicine at Mount Sinai, New York, NY, USA; Department of Psychiatry, Icahn School of Medicine at Mount Sinai, New York, NY, USA; The Mindich Child Health and Development Institute, Icahn School of Medicine at Mount Sinai, New York, NY, USA; Department of Environmental Medicine and Public Health, Icahn School of Medicine at Mount Sinai, New York, NY, USA

Alessandra Renieri: Med Biotech Hub and Competence Center, Department of Medical Biotechnologies, University of Siena, Siena, Italy; Medical Genetics, , University of Siena, Siena, Italy; Genetica Medica, Azienda Ospedaliera Universitaria Senese, Siena, Italy

Kathryn Roeder: Department of Statistics and Data Science, Carnegie Mellon University, Pittsburgh, PA, USA; Computational Biology Department, Carnegie Mellon University, Pittsburgh, PA, USA

Stephan J. Sanders: Department of Psychiatry, UCSF Weill Institute for Neurosciences, University of California San Francisco, San Francisco, CA, USA

Sven Sandin: Seaver Autism Center for Research and Treatment, Icahn School of Medicine at Mount Sinai, New York, NY, USA; Department of Psychiatry, Icahn School of Medicine at Mount Sinai, New York, NY, USA; Department of Medical Epidemiology and Biostatistics, Karolinska Institutet, Stockholm, Sweden

F. Kyle Satterstrom: Program in Medical and Population Genetics, Broad Institute of MIT and Harvard, Cambridge, MA, USA; Stanley Center for Psychiatric Research, Broad Institute of MIT and Harvard, Cambridge, MA, USA; Analytic and Translational Genetics Unit, Department of Medicine, Massachusetts General Hospital, Boston, MA, USA

Stephen W. Scherer: Program in Genetics and Genome Biology, The Centre for Applied Genomics, The Hospital for Sick Children, Toronto, Ontario, Canada; Department of Molecular Genetics and McLaughlin Centre, University of Toronto, Toronto, Ontario, Canada

Sabine Schlitt: Department of Psychiatry, Graduate School of Medicine, Nagoya University, Nagoya, Japan

Rebecca J. Schmidt: MIND (Medical Investigation of Neurodevelopmental Disorders) Institute, University of California Davis, Davis, CA, USA

Lauren Schmitt: Institute for Juvenile Research, Department of Psychiatry, University of Illinois at Chicago, Chicago, IL, USA

Katja Schneider-Momm: Department of Psychiatry, Graduate School of Medicine, Nagoya University, Nagoya, Japan

Paige M. Siper: Seaver Autism Center for Research and Treatment, Icahn School of Medicine at Mount Sinai, New York, NY, USA; Department of Psychiatry, Icahn School of Medicine at Mount Sinai, New York, NY, USA; The Mindich Child Health and Development Institute, Icahn School of Medicine at Mount Sinai, New York, NY, USA

Laura Sloofman: Seaver Autism Center for Research and Treatment, Icahn School of Medicine at Mount Sinai, New York, NY, USA; Department of Psychiatry, Icahn School of Medicine at Mount Sinai, New York, NY, USA; The Mindich Child Health and Development Institute, Icahn School of Medicine at Mount Sinai, New York, NY, USA

Moyra Smith: Center for Autism Research and Translation, University of California Irvine, Irvine, CA, USA

Christine R. Stevens: Program in Medical and Population Genetics, Broad Institute of MIT and Harvard, Cambridge, MA, USA; Stanley Center for Psychiatric Research, Broad Institute of MIT and Harvard, Cambridge, MA, USA; Analytic and Translational Genetics Unit, Department of Medicine, Massachusetts General Hospital, Boston, MA, USA

Pål Suren: Norwegian Institute of Public Health, Oslo, Norway

James S. Sutcliffe: Department of Molecular Physiology & Biophysics and Psychiatry, Vanderbilt University School of Medicine, Nashville, TN, USA; Vanderbilt

Genetics Institute, Vanderbilt University School of Medicine, Nashville, TN, USA
John A. Sweeney: Department of Psychiatry, University of Cincinnati, Cincinnati, OH, USA
Michael E. Talkowski: Center for Genomic Medicine, Massachusetts General Hospital, Boston, MA, USA; Program in Medical and Population Genetics, Broad Institute of MIT and Harvard, Cambridge, MA, USA; Department of Neurology, Massachusetts General Hospital and Harvard Medical School, Boston, MA, USA; Stanley Center for Psychiatric Research, Broad Institute of MIT and Harvard, Cambridge, MA, USA; Program in Bioinformatics and Integrative Genomics, Harvard Medical School, Boston, MA, USA
Flora Tassone: MIND (Medical Investigation of Neurodevelopmental Disorders) Institute, University of California Davis, Davis, CA, USA; Department of Biochemistry and Molecular Medicine, University of California Davis, School of Medicine, Sacramento, CA, USA
Karoline Teufel: Department of Psychiatry, Graduate School of Medicine, Nagoya University, Nagoya, Japan
Elisabetta Trabetti: Department of Neurosciences, Biomedicine and Movement Sciences, Section of Biology and Genetics, University of Verona, Verona, Italy
Slavica Trajkova: Department of Medical Sciences, University of Torino, Turin, Italy
Maria del Pilar Trelles: Seaver Autism Center for Research and Treatment, Icahn School of Medicine at Mount Sinai, New York, NY, USA; Department of Psychiatry, Icahn School of Medicine at Mount Sinai, New York, NY, USA
Brie Wamsley: Program in Neurogenetics, Department of Neurology, David Geffen School of Medicine, University of California Los Angeles, Los Angeles, CA, USA
Jaqueline Y. T. Wang: Centro de Pesquisas sobre o Genoma Humano e Células tronco, Instituto de Biociências, Universidade de São Paulo, São Paulo, Brazil
Lauren A. Weiss: Department of Psychiatry, UCSF Weill Institute for Neurosciences, University of California San Francisco, San Francisco, CA, USA
Mullin H. C. Yu : Department of Pediatrics and Adolescent Medicine, Duchess of Kent Children’s Hospital, The University of Hong Kong, Hong Kong Special Administrative Region, China
Ryan Yuen: Program in Genetics and Genome Biology, The Centre for Applied Genomics, The Hospital for Sick Children, Toronto, Ontario, Canada

***Autism Prenatal and Sex Differences (APEX):***

Dwaipayan Adhya: Department of Psychiatry, Autism Research Centre, University of Cambridge, Cambridge, Cambridgeshire, CB2 8AH, UK; Department of Basic and Clinical Neuroscience, Maurice Wohl Clinical Neuroscience Institute, Institute of Psychiatry, Psychology and Neuroscience, King's College London, London, UK

Carrie Allison: Department of Psychiatry, Autism Research Centre, University of Cambridge, Cambridge, Cambridgeshire, CB2 8AH, UK

Bonnie Ayeung: Department of Psychiatry, Autism Research Centre, University of Cambridge, Cambridge, Cambridgeshire, CB2 8AH, UK

Rosie Bamford: University of Exeter Medical School, Exeter, Devon, EX2 5DW, UK

Simon Baron-Cohen: Department of Psychiatry, Autism Research Centre, University of Cambridge, Cambridge, Cambridgeshire, CB2 8AH, UK

Richard Bethlehem: Department of Psychiatry, Autism Research Centre, University of Cambridge, Cambridge, Cambridgeshire, CB2 8AH, UK; Department of Psychiatry, Brain Mapping Unit, University of Cambridge, Cambridge, Cambridgeshire, CB2 8AH, UK

Tal Biron-Shental: Department of Obstetrics and Gynecology, Meir Medical Center, Kfar Saba, Israel; Sackler Faculty of Medicine, Tel Aviv University, Tel Aviv, Israel

Graham Burton: Centre for Trophoblast Research, Department of Physiology, Development and Neuroscience, University of Cambridge, Cambridge, CB2 3EG, UK

Wendy Cowell: University of Exeter Medical School, Exeter, Devon, EX2 5DW, UK

Jonathan Davies: University of Exeter Medical School, Exeter, Devon, EX2 5DW, UK

Dori Floris: Department of Cognitive Neuroscience, Donders Institute for Brain Cognition and Behaviour, Radboud University Medical Centre, 6525EN Nijmegen, The Netherlands; Methods of Plasticity Research, Department of Psychology, University of Zurich, Zurich, Switzerland

Alice Franklin: University of Exeter Medical School, Exeter, Devon, EX2 5DW, UK

Lidia Gabis: Tel Aviv University, Wolfson Hospital and Maccabi healthcare, Tel Aviv, Israel

Daniel Geschwind: Program in Neurobehavioral Genetics, Center for Autism Research and Treatment, Semel Institute, David Geffen School of Medicine, University of California, Los Angeles, Los Angeles, CA 90095, USA; Department of Neurology, David Geffen School of Medicine, University of California, Los Angeles, 695 Charles E. Young Drive South, Los Angeles, CA 90095, USA; Department of Psychiatry, Semel Institute, David Geffen School of Medicine, University of California, Los Angeles, 695 Charles E. Young Drive South,

Los Angeles, CA 90095, USA; Department of Human Genetics, David Geffen School of Medicine, University of California, Los Angeles, CA 90095, USA

David M. Greenberg: Interdisciplinary Department of Social Sciences, Bar-Ilan University, Ramat Gan, Israel; Department of Music, Bar-Ilan University, Ramat Gan, Israel; Autism Research Centre, Department of Psychiatry, University of Cambridge, Cambridge, UK

Yuanjun Gu: Department of Psychiatry, Autism Research Centre, University of Cambridge, Cambridge, Cambridgeshire, CB2 8AH, UK

Alexandra Havdahl: Center for Genetic Epidemiology and Mental Health, Norwegian Institute of Public Health, Oslo, Norway; Nic Waals Institute, Lovisenberg Diakonale Hospital, Oslo, Norway; PROMENTA Research Center, Department of Psychology, University of Oslo, Oslo, Norway; MRC Integrative Epidemiology Unit, University of Bristol, Bristol, UK

Alexander Heazell: Maternal and Fetal Health Research Centre, School of Medical Sciences, University of Manchester, Manchester, UK; Saint Mary's Hospital, Manchester University NHS Foundation Trust, Manchester, UK

Rosemary Holt: Department of Psychiatry, Autism Research Centre, University of Cambridge, Cambridge, Cambridgeshire, CB2 8AH, UK

Matthew Hurles: Wellcome Sanger Institute, Hinxton, Cambridgeshire, CB10 1SA, UK

Yumnah Khan: Department of Psychiatry, Autism Research Centre, University of Cambridge, Cambridge, Cambridgeshire, CB2 8AH, UK

Meng-Chuan Lai: Department of Psychiatry, Autism Research Centre, University of Cambridge, Cambridge, Cambridgeshire, CB2 8AH, UK; The Margaret and Wallace McCain Centre for Child, Youth & Family Mental Health and Azrieli Adult Neurodevelopmental Centre, Campbell Family Mental Health Research Institute, Centre for Addiction and Mental Health, Toronto, Canada; Department of Psychiatry and Autism Research Unit, The Hospital for Sick Children, Toronto, Canada; Department of Psychiatry, Temerty Faculty of Medicine, University of Toronto, Toronto, Canada; Department of Psychiatry, National Taiwan University Hospital and College of Medicine, Taipei, Taiwan

Madeline Lancaster: MRC Laboratory for Molecular Biology, University of Cambridge, Cambridge, UK

Michael Lombardo: Laboratory for Autism and Neurodevelopmental Disorders, Center for Neuroscience and Cognitive Systems, Istituto Italiano di Tecnologia, Rovereto, Italy

Hilary Martin: Wellcome Sanger Institute, Hinxton, Cambridgeshire, CB10 1SA, UK
Jose Gonzalez Martinez: MRC Laboratory for Molecular Biology, University of Cambridge, Cambridge, UK
Jonathan Mill: University of Exeter Medical School, Exeter, Devon, EX2 5DW, UK
Mahmoud Koko Musa: Wellcome Sanger Institute, Hinxton, Cambridgeshire, CB10 1SA, UK
Kathy Niakan: Cambridge Reproduction, University of Cambridge, Cambridge, UK; The Centre for Trophoblast Research, Department of Physiology, Development and Neuroscience, University of Cambridge, Cambridge, UK; Human Embryo and Stem Cell Laboratory, The Francis Crick Institute, London, UK; Wellcome Trust-Medical Research Council Stem Cell Institute, University of Cambridge, Jeffrey Cheah Biomedical Centre, Cambridge, UK; Epigenetics Programme, Babraham Institute, Cambridge, UK
Adam Pavlinek: Department of Basic and Clinical Neuroscience, Institute of Psychiatry, Psychology and Neuroscience, King's College London, London, UK; MRC Centre for Neurodevelopmental Disorders, King's College London, London, UK
Lucia Dutan Polit: Department of Basic and Clinical Neuroscience, Institute of Psychiatry Psychology and Neuroscience, King's College London, London, UK; MRC Centre for Neurodevelopmental Disorders, King's College London, London, UK
Marcin Radecki: Department of Psychiatry, Autism Research Centre, University of Cambridge, Cambridge, Cambridgeshire, CB2 8AH, UK; Social and Affective Neuroscience Group, IMT School for Advanced Studies Lucca, Lucca 55100, Italy
David Rowitch: Wellcome-MRC Cambridge Stem Cell Institute, University of Cambridge, Cambridge CB2 0AW, UK; Department of Paediatrics, University of Cambridge, Cambridge, UK
Laura Sichlinger: Department of Basic and Clinical Neuroscience, The Maurice Wohl Clinical Neuroscience Institute, Institute of Psychiatry, Psychology & Neuroscience, King's College London, London, UK; MRC Centre for Neurodevelopmental Disorders, Institute of Psychiatry, Psychology & Neuroscience, King's College London, London, UK
Deepak Srivastava: Department of Basic and Clinical Neuroscience, Institute of Psychiatry Psychology and Neuroscience, King's College London, London, UK; MRC Centre for Neurodevelopmental Disorders, King's College London, London, UK

Alexandros Tsompanidis: Department of Psychiatry, Autism Research Centre, University of Cambridge, Cambridge, Cambridgeshire, CB2 8AH, UK
Florina Uzefovsky: Ben-Gurion University of the Negev, Beer-Sheva, 84105, Israel
Varun Warrier: Department of Psychiatry, Autism Research Centre, University of Cambridge, Cambridge, Cambridgeshire, CB2 8AH, UK
Elizabeth Weir: Department of Psychiatry, Autism Research Centre, University of Cambridge, Cambridge, Cambridgeshire, CB2 8AH, UK
Xinhe Zhang: Department of Psychiatry, Autism Research Centre, University of Cambridge, Cambridge, Cambridgeshire, CB2 8AH, UK

### Supplementary Methods

#### 1. SPARK dataset preparation

##### 1.1. Exome Sequencing and phenotypic data

We downloaded the Simons Foundation Powering Autism Research for Knowledge (SPARK) integrated whole-exome sequencing data[1] release version 2 through Globus.[2] This release (jointly called VCF v2.2023_01, Deep Variant/GLNexus) encompassed 106,744 samples from five sequencing batches (WES1: 27,177 samples, WES2: 15,671, WES3: 16,476, WES4: 11,340, WES5: 36,080), sequenced using two capture systems (WES1-4: IDT, WES5: TWIST). The dataset included 44,304 autistic individuals (33,335 males and 10,969 females) and 62,440 individuals without an autism diagnosis (24,783 males and 37,657 females), including 27,775 father-child-pairs (19,977 autistic individuals) and 46,343 mother-child pairs (34,011 autistic individuals). These formed 25,386 trios (18,172 autistic individuals), 23,346 samples with one sequenced parent (17,644 autistic individuals), and 58,012 samples without parental WES (8,488 autistic individuals). The phenotypic data for the autistic individuals were downloaded from SFARI's Genotypes and Phenotypes in Families (GPF) database.[3] These phenotypic data were available for 40,346 autistic individuals with WES data.

##### 1.2. Variant annotation

The provided dataset included 10,202,547 sites (11,236,834 after splitting multiallelic sites). These sites were annotated using Variant Effect Predictor[4] (VEP) Ensembl release 108. The following annotations were added: consequences on Ensembl genes and transcripts, exon/intron numbers, allele frequencies based on gnomAD exomes (r2.1.1) and gnomAD genomes (r3), MPC scores, Loftee predictions, NMD plugin predictions.[5] The consequences were annotated on 'MANE Plus Clinical' transcripts (Matched Annotation between NCBI and Ensembl v1). For variants with consequences on more than one MANE transcript, one consequence was prioritized based on the severity of the predicted effects or gene constraint. We produced a working dataset in which we split multiallelic sites, set any variants with depth zero to missing, and filtered for variants that were in the exonic regions or the adjacent splice regions of MANE/Clinical transcripts.

#### 1.3. Ancestry inference

SPARK provided ancestry labels based on the 1000 Genomes super-populations (Africans [AFR], Admixed Americans [AMR], East Asians [EAS], Europeans [EUR], and South Asians [SAS]), along with subpopulation labels, and probabilities for being assigned to these sub-populations. A group of 2,783 samples were labeled as having an unknown super-population (Table S1). To confirm these labels and to classify the samples in the 'Unknown' group to the nearest population when possible, we projected SPARK iWES2 samples onto a principal components (PCs) space based on 2,536 samples from the 1000 Genomes Project[6] calculated using PLINK.[7] PC analysis of the 1000 Genomes reference data (10 PCs) leveraged 8,718 pruned variants with allele frequency > 1% that are present in at least one sample in SPARK samples, limited to exonic regions of protein coding transcripts described above. SPARK samples were then projected into the 1000 Genomes PC space. The clustering of these samples was contrasted with the pre-defined ancestry labels. We reclassified the samples where the superpopulation was labeled as 'unknown' to one of the 1000 Genomes superpopulations if these had a probability exceeding 0.8 of belonging to that group (calculated as the sum of probabilities of belonging to the sub-populations under that group, provided by SPARK), or clustered with that group on each of the first four PCs. Last, we defined a group of admixed individuals that did not cluster closely with their corresponding groups on PC1-4 (Table S2 and Figure S1).

#### 1.4. Sex inference

To infer the ploidy of sex chromosomes from sequencing data, we evaluated the depth and genotypes in the hemizygous regions of chrX and chrY, namely, (1) the read depth in chrX normalized by the median chrX depth in samples labeled as males, (2) the read depth in chrY normalized by the median chrY depth in samples labeled as males, (3) the fraction of missing calls on chrY, and (4) the F statistic (the difference between the expected and observed heterozygosity in the hemizygous region of chrX; PLINK). Male sex was inferred for samples with normalized chrY depth between 0.5 and 2, fraction of missing genotypes in chrY < 50%, normalized chrX depth < 2, and F statistic > 0.8. Female sex was inferred for samples with normalized chrX depth between 1 and 3, F statistic > -0.6 and < 0.6, normalized chrY depth < 0.15, and fraction of missing genotypes on chrY > 50%. 122 male and 88 female samples were classified as having ambiguous inferred sex or having a mismatch between reported and inferred sex, and were removed from the burden analyses.

Table S1: SPARK samples per population before QC.

| Sequencing | Group | Sub-group | AFR | AMR | EAS | EUR | SAS | UNK | | Total | |
|---|---|---|---|---|---|---|---|---|---|---|---|
| | | **Fathers** | 1087 | 1877 | 467 | 13258 | 549 | 317 | 17555 | 47698 | 58012 |
| | **Parents** | **Mothers** | 2395 | 3182 | 781 | 22709 | 566 | 510 | 30143 | | |
| | | **Males** | 551 | 576 | 127 | 3670 | 85 | 142 | 5151 | 7841 | |
| | **Probands** | **Females** | 240 | 219 | 77 | 2072 | 24 | 58 | 2690 | | |
| | | **Males** | 86 | 127 | 40 | 843 | 21 | 33 | 1150 | 2473 | |
| **Single sample** | **Siblings** | **Females** | 126 | 153 | 32 | 962 | 21 | 29 | 1323 | | |
| | | **Fathers** | 0 | 1 | 0 | 10 | 0 | 0 | 11 | 35 | 23346 |
| | **Parents** | **Mothers** | 2 | 2 | 0 | 20 | 0 | 0 | 24 | | |
| | | **Males** | 1768 | 1714 | 156 | 9479 | 156 | 418 | 13691 | 17611 | |
| | **Probands** | **Females** | 502 | 427 | 46 | 2807 | 36 | 102 | 3920 | | |
| **Child-parent** | | **Males** | 285 | 331 | 28 | 1985 | 29 | 85 | 2743 | 5700 | |
| **pair** | **Siblings** | **Females** | 315 | 364 | 26 | 2138 | 30 | 84 | 2957 | | |
| | | **Fathers** | 0 | 0 | 0 | 8 | 0 | 0 | 8 | 19 | 25386 |
| | **Parents** | **Mothers** | 0 | 0 | 0 | 11 | 0 | 0 | 11 | | |
| | | **Males** | 893 | 1770 | 302 | 10306 | 427 | 553 | 14251 | 18156 | |
| | **Probands** | **Females** | 240 | 463 | 63 | 2867 | 93 | 179 | 3905 | | |
| | | **Males** | 167 | 388 | 61 | 2715 | 86 | 141 | 3558 | 7211 | |
| **Trio** | **Siblings** | **Females** | 204 | 442 | 57 | 2739 | 79 | 132 | 3653 | | |
| **All** | | | 8861 | 12036 | 2263 | 78599 | 2202 | 2783 | 106744 | | |

AFR: Africans, AMR: Admixed Americans, EAS: East Asians, EUR: Europeans, SAS: South Asians, UNK: Unknown (admixed).

Table S2: SPARK samples per population after PCA.

| Sequencing strategy | Group | Sub-group | AFR | AMR | EAS | EUR | SAS | UNK |
|---|---|---|---|---|---|---|---|---|
| **Before PCA** | | | 8861 | 12036 | 2263 | 78599 | 2202 | 2783 |
| | | **Fathers** | 1079 | 2177 | 471 | 13258 | 540 | 30 |
| | **Parents** | **Mothers** | 2384 | 3654 | 797 | 22709 | 558 | 41 |
| | | **Males** | 547 | 713 | 130 | 3670 | 83 | 8 |
| | **Probands** | **Females** | 238 | 272 | 80 | 2072 | 23 | 5 |
| | | **Males** | 85 | 160 | 40 | 843 | 21 | 1 |
| **Single sample** | **Siblings** | **Females** | 126 | 182 | 32 | 962 | 21 | 0 |
| | | **Fathers** | 0 | 1 | 0 | 10 | 0 | 0 |
| | **Parents** | **Mothers** | 2 | 2 | 0 | 20 | 0 | 0 |
| | | **Males** | 1764 | 2113 | 160 | 9479 | 153 | 22 |
| | **Probands** | **Females** | 501 | 525 | 47 | 2807 | 36 | 4 |
| | | **Males** | 285 | 412 | 29 | 1985 | 27 | 5 |
| **Child-parent pair** | **Siblings** | **Females** | 315 | 445 | 27 | 2138 | 30 | 2 |
| | | **Fathers** | 0 | 0 | 0 | 8 | 0 | 0 |
| | **Parents** | **Mothers** | 0 | 0 | 0 | 11 | 0 | 0 |
| | | **Males** | 890 | 2304 | 307 | 10306 | 417 | 27 |
| | **Probands** | **Females** | 237 | 638 | 63 | 2867 | 89 | 11 |
| | | **Males** | 166 | 527 | 62 | 2715 | 83 | 5 |
| **Trio** | **Siblings** | **Females** | 204 | 568 | 57 | 2739 | 78 | 7 |
| **All** | | | 8823 | 14693 | 2302 | 78599 | 2159 | 168 |

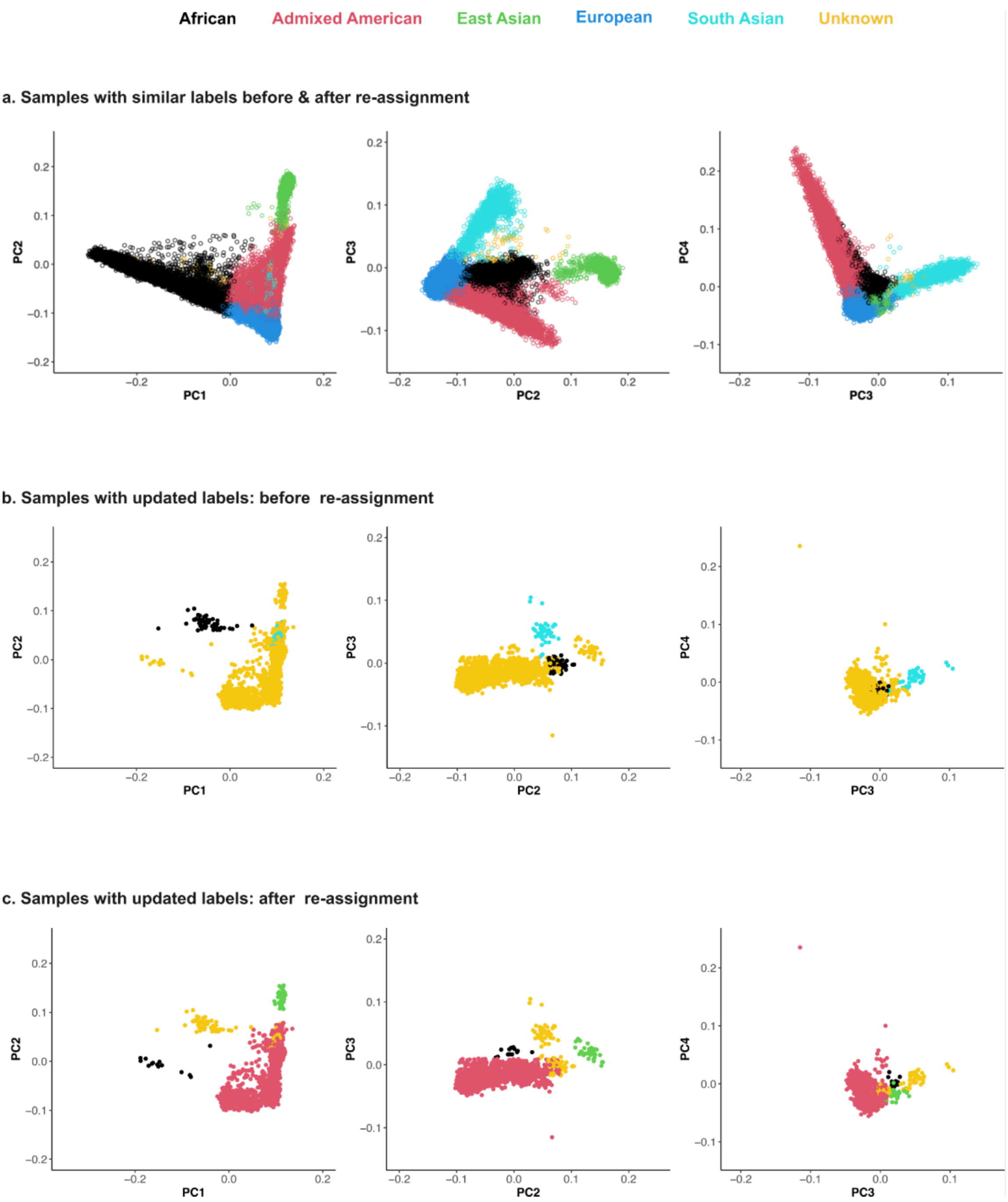


Figure S1. Population labels validation.

Principal component analysis (PCA) was used to check the validity of the pre-assigned population labels from SPARK against the top principal components projected on the 1000 Genomes space (reference samples not shown). Most samples clustered as expected (**a**). Samples with unknown labels and those not clustering with their respective pre-assigned groups (**b**) were re-classified (**c**). See section 1.3. 'Ancestry Inference' and Tables S1 & S2 for details.

#### 1.5. Relatedness inference

The provided meta-data indicated the presence of 27,775 father-child-pairs and 46,343 mother-child pairs (25,386 complete trios). To verify the parental relationships, kinship was estimated using PLINK based on KING's robust algorithm.[8] We required the parent-child pairs to have a kinship estimate between 0.3 and 0.1 and a proportion of SNPs with Identity By State (IBS) sharing < $5x10^{-5}$. Ten parent-child pairs could not be verified based on kinship. A set of maximally unrelated probands (N=39,020) was selected by incrementally removing the samples with the highest number of related autistic individuals (kinship > 0.0884) while prioritizing those with sequenced parents and females in that order. Ties were resolved by retaining the sample with the older identifier. Similar sets of unrelated individuals were defined for the siblings (N=15,056) and parents (N=47,525). We estimated the allele frequencies, Hardy Weinberg Equilibrium *p*-values, and Excess Heterozygosity *p*-values separately in these three sets of unrelated samples (per population) and used the largest *p*-value (i.e. the least significant) as an input for variant quality control (QC).

#### 1.6. Variant QC

A Random Forest was trained to identify low quality variant call sites. It was applied separately on SNVs and INDELs and the training sites were selected from variants heterozygous in one or more samples. Before running the variant QC, we applied basic genotype filtering by setting the genotypes that had any of the following to missing: (1) depth equal to zero, (2) genotype quality (GQ) equal to zero, (3) a conflict between the called genotype and the phred-scaled genotype likelihood, or (4) a *p*-value from a binomial test for allele balance less that $1x10^{-10}$ (for heterozygous calls). Negative training sites were then defined as those that met one of the following criteria, excluding multi-allelic sites: (1) more than 50% of the samples with non-reference (heterozygous or homozygous) genotypes that had GQ < 20, (2) more than 50% of samples with a heterozygous genotype had variant allele fraction (VAF) below 0.2, or (3) the Hardy-Weinberg Equilibrium test *p*-value (calculated for separately for unrelated parents, siblings and probands per population, taking the largest estimate across all populations) was lower than $10^{-9}$. The positive training sites were the high confidence sites from the Broad bundle resource for (SNVs: Omni, Axiom, HapMap; Indels: Axiom, Mills) used in the Genome Analysis Toolkit (GATK) best practices[9]). Since these sites are enriched for common variants, we annotated an equal number of randomly selected high confidence transmitted singleton variants (seen in a single child-parent pair, with Allele Quality > 35) as positive training sites. To balance the number of positive (SNVs: 388,268; INDELs: 17,732) and negative (SNVs: 98,676; INDELs: 3,961) training sites, the positive training set was down-sampled to match the size of the negative training set, resulting in a balanced set of 205,274 variants (SNV = 197,352, 2.8% of all SNVs, INDEL=7,922, 2.3% of all INDELs). The remaining sites were annotated as test sites (SNVs= 6,731,744, INDELs= 310,362).

We used the `ranger` package[10] in `R 4.1.0` to train random forest models on SNVs and INDELs separately, with 500 trees and probabilities as an output. The features used to train the random forest, and their relative importance, are depicted in Figure S2a. The training sites did not include any missing values in these features, and missing values among the test sites were set to the median of the respective feature across all sites of similar type (SNV or INDEL). The random forest model was then applied on all training and test sites to obtain probabilistic scores (range 0-1) indicating the probability of being a truly variable site. These scores were scaled to the range 0-100. The performance of the random forest was evaluated using precision and recall against the training sites using the `caret` package[11] in `R`. We then examined the fraction of retained sites from the total dataset and the transmission ratio of synonymous variants with incremental random forest cut-offs. Random Forest score cut-offs of 97 for SNVs (recall = 0.97, precision = 1) and 92 for indels (recall = 0.975, precision = 1.0) were selected, and sites with scores higher than these were retained. The transmitted/untransmitted ratio of autosomal synonymous variants was 0.5 (Figure S2b). Next, the variants were filtered to those with a minimum genotyping rate (per sequencing batch; across five sequencing batches) exceeding 95%. After this QC, 6,345,636 out of 7,273,162 sites were retained (12.8% filtered). These variants spanned 18,959 MANE-Plus transcripts (18,902 protein coding genes; including 57 genes with two transcripts).

#### 1.7. Genotype filtering

We applied genotype filters based on genotype quality, allele depth, variant allele fraction, by setting the genotypes to missing if they were not consistent with the observed phred-scaled genotype likelihoods (PLs), had GQ < 10, had DP < 10, were reference genotypes with VAF > 0.25, were homozygous with VAF < 0.75, or were heterozygous with VAF < 0.2 or > 0.8.

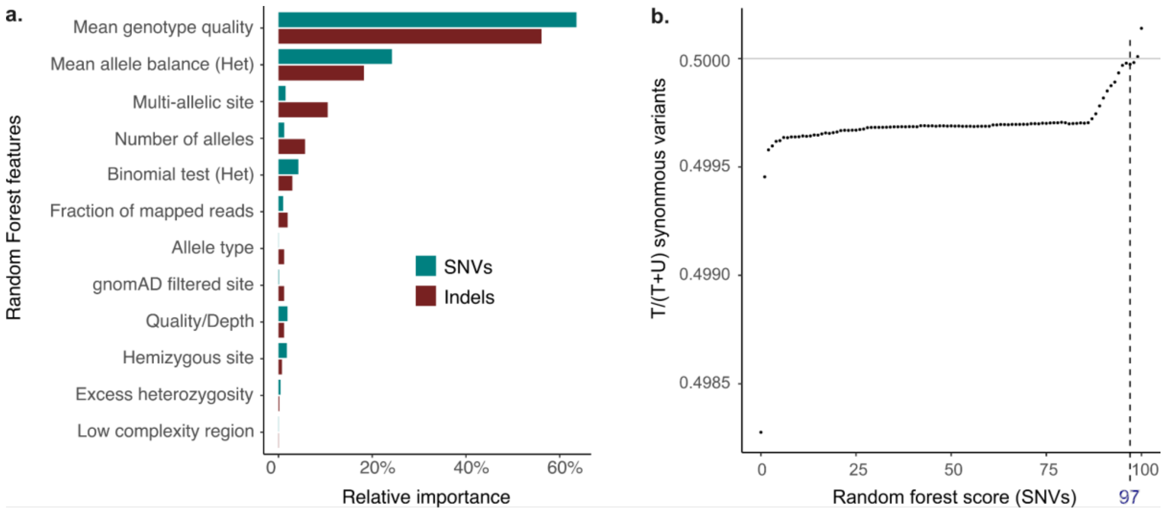


Figure S2. Assigning variant quality scores using Random Forest.

A Random Forest was trained using 12 features (**a**) to assign a variant-level quality score between 0 (lowest quality) and 100 (highest quality). The average genotype quality per variant (all genotypes) and mean allele balance per variant (heterozygous genotypes) had the highest relative importance (y-axis). The fraction of the transmitted (T) alleles from the total transmitted & untransmitted (T+U) parental synonymous variants achieved after filtering with increasing stringency (higher Random Forest scores) is shown in **b**. Single Nucleotide Variants (SNVs) with scores > 97 (dotted line) and Insertions-Deletions (Indels) with scores > 92 were retained. See section 1.6. (Variant QC) for further details.

### 1.8. Sample QC

We calculated the following sample-level metrics: total number of SNVs, total number of INDELs, total number of private variants (seen in one family), transition-transversion ratio, insertion-deletion ratio, and heterozygous-homozygous variants ratio. We regressed these on the population label, four principal components and the number of sequenced samples per family, to account for the population structure and the differences in variant-calling sensitivity in related individuals. We filtered all samples that were more than six standard deviations from the mean of the residuals for any metric, removing 275 samples (AFR: 180, EUR: 55, AMR: 22, EAS: 3, SAS: 6, Unknown/Admixed: 9). Parents with autism and parents and siblings with motor delay or cognitive impairment were filtered (see 1.9). Subsequently, sets of maximally unrelated probands and siblings were selected. Note that this unrelated dataset of probands and siblings was used for the formal analysis of rare variant enrichment, and is prepared independently from the unrelated set presented in section 1.5, which was used to estimate the allele frequency for variant QC. Table S3 shows the sample size per population group after the sample QC.

#### 1.9. Motor delay and cognitive impairment

We used the following phenotypic data fields to stratify the samples based on the presence of a motor developmental disorder, cognitive impairment or an intellectual disability diagnosis:

- ‘cognitive_impairment_at_enrollment’ and ‘cognitive_impairment_latest’: provided information on cognitive impairment diagnosis (identical values). Probands with a diagnosis were coded as “True” and those without diagnosis were coded as “False”. Missing values were coded as “-”.
- 'dev_id' and 'dev_motor': provided information on cognitive impairment (Intellectual disability, cognitive impairment, global developmental delay, or borderline intellectual functioning, reported professional diagnosis) and motor development (Motor delay, e.g., delay in walking, or developmental coordination disorder; reported professional diagnosis). Diagnoses were coded as “1”. Typical development and missing values were coded as “-”.
- ‘cog_test_score’ and the ‘reported_cog_test_score’: provided binned full-scale IQ measurements (identical values). Missing values were coded as “-”.

We classified the autistic individuals in three phenotypic groups:

1. Autism with motor delay or cognitive impairment:
   ‘Cognitive_impairment_at_enrollment’, *or*
   ‘Cognitive_impairment_latest’ = “True”, *or*
   ‘dev_motor’ or ‘dev_id’ = 1, *or*
   ‘Cog_test_score’, *or*
   ‘reported_cog_test_score’ = ‘24_below’,‘25_39’, ‘40_54’, ‘55_69’, or ‘70_79’.
2. Autism without motor or cognitive impairment:
   ‘Cognitive_impairment_at_enrollment’, *and*
   ‘Cognitive_impairment_latest’ = “False”, *and*
   IQ bin equivalent to an IQ >= 80 or missing IQ data, *and*
   missing values in the ‘dev_motor’ field, *and*
   missing values in the ‘dev_id’ field.

The remaining probands with missing values in all fields were considered unclassified.

Table S3A: SPARK trio-sequenced samples (after sample QC).

| Group | | AFR | AMR | EAS | EUR | SAS | UNK | Total | |
|---|---|---|---|---|---|---|---|---|---|
| **Before sample QC** | | 1497 | 4037 | 489 | 18646 | 667 | 50 | 25386 | |
| **Parents** | **Fathers** | 0 | 0 | 0 | 2 | 0 | 0 | 2 | 3 |
| | **Mothers** | 0 | 0 | 0 | 1 | 0 | 0 | 1 | |
| **Siblings** | **Males** | 148 | 492 | 60 | 2519 | 81 | 5 | 3305 | 6763 |
| | **Females** | 181 | 543 | 56 | 2596 | 76 | 6 | 3458 | |
| **Probands** | **Males** | 646 | 1706 | 237 | 7536 | 340 | 17 | 10482 | 13473 |
| | **Females** | 190 | 504 | 51 | 2163 | 77 | 6 | 2991 | |
| **With motor/cog. imp.** | **Males** | 202 | 488 | 48 | 2369 | 97 | 1 | 3205 | 4209 |
| | **Females** | 55 | 154 | 20 | 750 | 23 | 2 | 1004 | |
| **No motor/cog. imp.** | **Males** | 365 | 1017 | 150 | 4217 | 197 | 10 | 5956 | 7420 |
| | **Females** | 101 | 285 | 23 | 1008 | 44 | 3 | 1464 | |
| **Unknown** | **Males** | 79 | 201 | 39 | 950 | 46 | 6 | 1321 | 1844 |
| | **Females** | 34 | 65 | 8 | 405 | 10 | 1 | 523 | |
| **All** | | 1165 | 3245 | 404 | 14817 | 574 | 34 | 20239 | |

Table S3B: SPARK samples with one sequenced parent (after sample QC).

| Group | | AFR | AMR | EAS | EUR | SAS | UNK | Total | |
|---|---|---|---|---|---|---|---|---|---|
| **Before sample QC** | | 2867 | 3498 | 263 | 16439 | 246 | 33 | 23346 | |
| **Parents** | **Fathers** | 0 | 0 | 0 | 0 | 0 | 0 | 0 | 2 |
| | **Mothers** | 0 | 0 | 0 | 2 | 0 | 0 | 2 | |
| **Siblings** | **Males** | 258 | 395 | 29 | 1849 | 26 | 5 | 2562 | 5381 |
| | **Females** | 301 | 427 | 27 | 2032 | 30 | 2 | 2819 | |
| **Probands** | **Males** | 1342 | 1600 | 135 | 7186 | 129 | 19 | 10411 | 13435 |
| | **Females** | 386 | 421 | 39 | 2146 | 28 | 4 | 3024 | |
| **With motor/cog. imp.** | **Males** | 521 | 544 | 42 | 2575 | 39 | 3 | 3724 | 4741 |
| | **Females** | 152 | 135 | 13 | 707 | 8 | 2 | 1017 | |
| **No motor/cog. imp.** | **Males** | 677 | 901 | 73 | 3793 | 74 | 7 | 5525 | 6938 |
| | **Females** | 181 | 223 | 19 | 975 | 14 | 1 | 1413 | |
| **Unknown** | **Males** | 144 | 155 | 20 | 818 | 16 | 9 | 1162 | 1756 |
| | **Females** | 53 | 63 | 7 | 464 | 6 | 1 | 594 | |
| **All** | | 2287 | 2843 | 230 | 13215 | 213 | 30 | 18818 | |

Table S3C: SPARK samples without sequenced parents after sample QC.

| Group | | AFR | AMR | EAS | EUR | SAS | UNK | Total | |
|---|---|---|---|---|---|---|---|---|---|
| **Before sample QC** | | 4459 | 7158 | 1550 | 43514 | 1246 | 85 | 58012 | |
| **Parents** | **Fathers** | 1038 | 2149 | 468 | 12993 | 534 | 28 | 17210 | 46673 |
| | **Mothers** | 2301 | 3593 | 789 | 22193 | 551 | 36 | 29463 | |
| **Siblings** | **Males** | 79 | 154 | 40 | 828 | 20 | 1 | 1122 | 2436 |
| | **Females** | 120 | 178 | 32 | 961 | 22 | 1 | 1314 | |
| **Probands** | **Males** | 441 | 580 | 111 | 3094 | 72 | 8 | 4306 | 6646 |
| | **Females** | 209 | 234 | 67 | 1805 | 22 | 3 | 2340 | |
| **With motor/cog. imp.** | **Males** | 168 | 206 | 25 | 901 | 28 | 2 | 1330 | 1813 |
| | **Females** | 64 | 66 | 24 | 324 | 5 | 0 | 483 | |
| **No motor/cog. imp.** | **Males** | 187 | 259 | 54 | 1202 | 28 | 6 | 1736 | 2181 |
| | **Females** | 54 | 65 | 16 | 295 | 12 | 3 | 445 | |
| **Unknown** | **Males** | 86 | 115 | 32 | 991 | 16 | 0 | 1240 | 2652 |
| | **Females** | 91 | 103 | 27 | 1186 | 5 | 0 | 1412 | |
| **All** | | 4188 | 6888 | 1507 | 41874 | 1221 | 77 | 55755 | |

### 2. *De novo* and rare inherited variants in trios

As introduced in the main text, *de novo* mutations (DNMs) were studied in a total of 30,274 trios from the two cohorts by comparing DNM rates in 21,501 autistics (ASC: 8,028; SPARK: 13,473) to those seen in 8,773 siblings (ASC: 2,460; SPARK: 6,763). Over-transmission analysis of rare inherited variants was performed by evaluating the counts of rare parental alleles transmitted to 21,043 autistic individuals (ASC: 7,570; SPARK: 13,473) *versus* untransmitted alleles. This section outlines the curation of DNMs and inherited alleles in each cohort.

#### 2.1. ASC

DNMs in this dataset were obtained from previous ASC studies (see 'Methods: ASC cohort'). These were curated in 9,929 trios (7,570 autistics, 2,359 siblings) as described in detail in the Supplementary Note of Ref.[12] Briefly, the `de_novo()` function from `Hail 0.2` python library[13] was used to identify candidate DNMs, with priors from population allele frequencies. These candidates were then filtered for rare alleles with internal (within ASC dataset) and external (gnomAD) allele frequencies < 0.1%, with additional filtering on genotype depth (allele balance and parent/child depth ratio), variant quality ('ExcessHet' filter and GATK variant quality score log-odds), as well as the number of candidate DNMs per sample. Additional published DNMs were included, bringing the total sample size to 10,488 trios (8,028 autistics, 2,460 siblings). Transmitted and untransmitted rare parental alleles were counted in 7,570 trios (autistic probands) with available exome data.

#### 2.2. SPARK

Following the variant, genotype and sample QC described in section 1 above (see 'SPARK dataset preparation'), we evaluated the genotypes of 20,236 trios (13,473 autistics and 6,763 siblings) for potential DNMs (see counts for probands and siblings in Table S3). We first filtered for candidate DNMs based on the parental genotypes and used the '`trio-dnm2`' plugin from `bcftools`[14] 1.17 to estimate the error probability of *de novo* inheritance based on allele depth (adDNM) and genotype likelihoods (plDNM). Then for each candidate DNM, we transformed the probabilities of it being a *de novo mutation* based on these two methods to a phred-scaled error probability ($-10*\log_{10}(\mathrm{minimum}(\mathrm{plDNM},\mathrm{adDNM}))$). We then defined putative DNMs as those with phred-scaled error probability ≤ 60 having internal & gnomAD

MAF < 0.1%, with an Allele Quality (AQ) score ≥ 30, and that had GQ ≥ 30 in all members of the trio. Finally, we selected one DNM per gene (worst consequence) per individual. This resulted in a dataset of 23,332 DNMs in 13,664 individuals (68% trios with DNMs, average rate of 1.15 DNMs per individual). Between 20% and 22% of the probands and siblings carried rare synonymous DNMs, compared to 23%-25% in the previous analysis of SPARK[12] (WES1), suggesting that our DNM callset was of similar quality to the published one. To ensure that the sensitivity to identify DNMs was not biased by sample sex, we calculated the odds ratio of carrying a rare *de novo* synonymous (all autosomal genes) or protein-truncating DNM (most-constrained LOEUF decile) among male versus female siblings, and found that it did not differ significantly from one (Figure S3b).

We also evaluated the transmission of rare variants (MAF < 0.1% in SPARK parents and gnomAD) in the same set of trio-sequenced autistic individuals. Similar to DNMs, we considered variants remaining after basic variant, genotype and sample QC. In instances where a single gene had multiple variants, the variant with the worst consequence was retained. We then filtered for heterozygous variants seen in one parent that had a GQ > 25 in all three samples in the trio to ensure the transmission ratio of synonymous variants was not significantly different from 1.

#### 2.3. Additional filtering of *de novo* mutations in SPARK

De novo mutations, especially damaging ones, are typically ultra-rare, usually seen in a few individuals or not seen at all in the general population. DNM calling from genotypes (see '2.2 SPARK' above) does not leverage allele frequencies as priors, and is best coupled with stringent allele frequency filtering by removing DNMs seen in more than a few individuals in the dataset or in the general population. Given that the ASC DNM dataset was prepared using a different pipeline and filtered for rare alleles (MAF < 0.1%), we adopted a similar cutoff in SPARK to have comparable call sets, thus facilitating fixed-effect meta-analysis of risk ratios. We then performed an additional analysis of ultra-rare DNMs in SPARK to ensure that the conclusions are the same in a call-set of high-confidence DNMs.

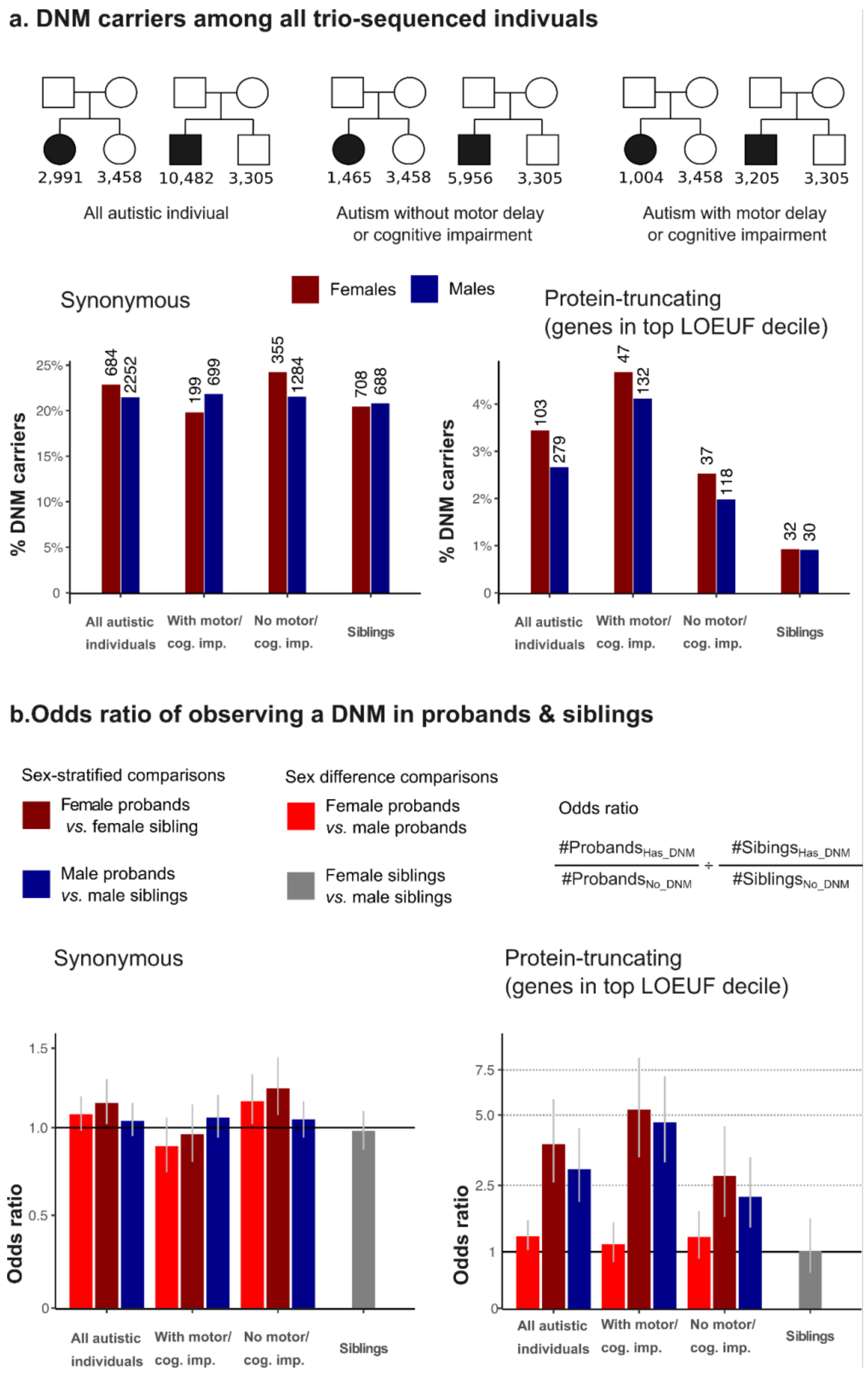


Figure S3. Odds of carrying a rare synonymous and damaging protein-truncating *de novo* mutation in SPARK trio-sequenced individuals.

Among 20,236 trios, about 20%-22% of the siblings had a synonymous de novo mutation (DNM), and less than 1% had a damaging protein-truncating DNM (**a**). There was no sex difference (measured using odds ratios as shown in **b**) in the sensitivity of detecting synonymous and protein-truncating DNMs among the siblings. The percentage of DNM carriers among the probands is shown for comparison. Autistic females without cognitive impairment had a slightly higher percentage of synonymous DNM carriers compared to autistic males in the same group. See the Supplementary Results (section 4.1.3) for more details.

To define ultra-rare DNMs, we further dropped DNMs if the individual had more than one DNM in the same gene, those seen as *de novo* in more than 3 individuals in SPARK, DNMs with MAF ≥ 0.005% in SPARK parents or gnomAD, and DNMs seen in gnomAD and two of the three SPARK cohorts of unrelated individuals used for estimating allele frequency (probands, siblings, parents). This final dataset included 15,072 ultra-rare DNMs, with an average rate of 0.74 ultra-rare DNMs per sample.

#### 2.4. Matching samples on principal components

As presented in the main text (Figure 2) and Supplementary Results (Figure S13; section 4.1.3), *de novo* synonymous mutations showed a spurious association with autism when comparing autistic females without motor or cognitive impairment to sex-matched probands with motor or cognitive impairment or when compared to female siblings. This likely reflects the greater diversity among autistic females without motor or cognitive impairment (69% of samples with European genetic ancestry) relative to the other two groups (75% of samples with European genetic ancestry in each; see Table S3 in section 1.8, 'Sample QC'). We performed an additional sensitivity analysis of rare and ultra-rare *de novo mutation* enrichment in a stringently matched set of samples to ensure that the results obtained for protein-truncating DNMs were not biased. Starting with 1,008 autistic females without cognitive impairment (`P`) and 2,596 female siblings (`S`) of European genetic ancestry (Table S3), we calculated the distance between all pairs of probands and siblings in the first four principal components (see section 1.3). Specifically, we subtracted the first four PCA eigenvectors for each proband `p` in `P` from the eigenvectors of each sibling `s` in `S`, then used the `norm(type="2")` function in `R` to calculate a Euclidean-type spectral norm representing the distance between the two samples in the PC space. We then took the minimum value for each sample in `P` and `S`, which represents the nearest neighboring sample from the opposite cohort. Samples with large values don't have any neighboring samples from the other group and are likely to be poorly-matched on ancestry. We excluded the outliers on this distance metric from the probands and siblings (> 2 median absolute deviations), leaving 868 autistic females and 2,235 siblings. See section 4.1.3 of the Supplementary Results for the outcomes of DNM enrichment analysis in this subset.

### 3. Ultra-rare variants

A substantial part of the ASC & SPARK datasets is formed of children with sequence data from one parent (N=18,816) or without any sequenced parents (N=23,114). Including these in analyses could potentially increase power. Specifically, they can be leveraged to study ultra-rare inherited alleles (when one parental exome is available) or the average effect of a mix of ultra-rare alleles of undetermined origin (case-control analysis), as detailed here. Similarly, ultra-rare variants ascertained in individuals without sequencing data from their parents ('case-control' cohorts) can be used to study the average effects of damaging *de novo* and inherited variants.

#### 3.1. Ultra-rare inherited variants in child-parent pairs in SPARK

To be able to leverage the exome data from parents not sequenced as complete trios (Table S3B), we studied the transmission of parental alleles in child-parent pairs, treating trios as two separate pairs rather than following the standard approach of comparing the transmission of rare variants in trios only. Studying transmission in parent-child pairs when sequence data is present from one parent only assumes the parents are not consanguineous (therefore unlikely to carry the same ultra-rare variant), and precludes the analysis of variants that are low-frequency but not extremely rare (as their transmission status cannot be determined reliably). Here, we examined ultra-rare parental alleles seen in one parent, in one family, and not seen in gnomAD. These were filtered for high-confidence calls (GQ > 32) to balance the transmission rates of synonymous variants.

#### 3.2. Case-control variants in the ASC & SPARK

We evaluated the enrichment & liability of ultra-rare variants in the ASC case-control dataset as a supplementary analysis. This dataset contained samples from the Danish iPSYCH cohort[15] and Swedish PAGES samples.[16] The processing and a previous sex-averaged analysis of these cohorts is described elsewhere.[12] Rare variants in iPSYCH were defined as those with an allele count ≤ 5 in the combined set of gnomAD non-Finnish Europeans (nonpsychiatric subset) and iPSYCH data (allele frequency ≤ 0.0043%). Rare variants in PAGES were defined as those with an allele count ≤ 5 in ExAC r0.3 (nonpsychiatric subset; allele frequency ≤ 0.0055%) and the autism cohort in Satterstrom (allele frequency ≤ 0.014%).

For SPARK, we analyzed ultra-rare variant enrichment in the remaining individuals (6,646 probands and 2,436 siblings) who did not have sequence data from any parent (Table S3C) - and as such were not included in all the previous analyses described above. This design approximates the case-control analysis in ASC. Whereas the ASC case-control dataset consisted of unrelated individuals, this SPARK sub-cohort included pairs of probands and siblings from the same family (like the trio-based analysis). We included variants seen with an allele frequency < 0.005% (in gnomAD and SPARK unrelated individuals' cohorts) that are seen in up to three individuals in the 'case-control' cohort of 9,082 individuals remaining after QC (allele frequency < 0.015%). We dropped 156 individuals with ultra-rare synonymous variants counts exceeding four median absolute deviations (> 33 variants), and compared the variant rates in the remaining 8,926 children (6,534 probands *versus* 2,392 siblings).

### 4. Measuring rare variant enrichment

In summary, we processed exome sequencing data from autism probands, siblings and parents from the latest release of SPARK and combined these data with data from the SSC and other smaller cohorts previously curated by the ASC. We then evaluated the enrichment of synonymous, damaging missense and damaging protein-truncating variants in a total of 131,970 individuals encompassing 47,601 autistic probands or autism cases, 25,593 non-autistic siblings or autism controls, and 59,316 parents (Table S4). Specifically, we performed these sex-stratified rare variant enrichment analyses:

1. Enrichment of *de novo* and rare inherited alleles in 20,501 autistic trios (*versus* 9,223 siblings).
2. A supplementary analysis over-transmission of ultra-rare variants in 13,435 autistic individuals and 5,381 siblings with one sequenced parent.
3. A supplementary case-control analysis of ultra-rare variants in 12,125 autism cases/probands *versus* 10,989 controls/siblings.

Table S4. Overview of the total sample size of autistic individuals, parents, siblings, and autism controls used in three sets of analyses in this study.

| | | Parents | | | Children | | | |
|---|---|---|---|---|---|---|---|---|
| Analysis | Cohort | Fathers | Mothers | Both | Probands Cases | Siblings Controls | Both | All |
| | **SPARK** | 14,569 | 14,568 | 29,137 | 13,473 | 6,763 | 20,236 | |
| | **ASC** | 7899 | 7898 | 15,797 | 8,028 | 2,460 | 10,488 | |
| **De novo & rare inherited** | **Both** | 22,468 | 22,466 | 44,934 | 21,501 | 9,223 | 30,724 | 75,658 |
| **Ultra-rare inherited** | **SPARK** | 1,566 | 12,816 | 14,382 | 13,435 | 5,381 | 18,816 | 33,198 |
| | **SPARK** | 0 | 0 | 0 | 6,534 | 2,392 | 8,926 | |
| | **ASC** | 0 | 0 | 0 | 5,591 | 8,597 | 14,188 | |
| **Ultra-rare ‘case-control’** | **Both** | 0 | 0 | 0 | 12,125 | 10,989 | 23,114 | 23,114 |
| **All samples** | | 24,034 | 35,282 | 59,316 | 47,061 | 25,593 | 72,654 | 131,970 |

#### 4.1. Variant rates

We calculated the *de novo* and inherited variant rates for synonymous, damaging missense and protein-truncating variants (most-constrained LOEUF decile) in each cohort (ASC, SPARK) by dividing the total number of variants by the sample size. The 95% Confidence Intervals (CI) were estimated in `R` as follows:

$$95\%\ CI_{lower} = qchisq(0.025, \frac{\#Variants}{2 * \#Samples})$$

$$95\%\ CI_{upper} = qchisq(0.975, \frac{\#Variants + 1}{2 * \#Samples})$$

#### 4.2. Sex-stratified comparisons

##### 4.2.1. De novo mutations

We used the ratio between the rate of DNMs in the probands and the siblings (or female and male probands in direct comparisons) to evaluate the enrichment of a specific class of DNMs in a gene set or exome-wide:

$$DNM\ rate\ ratio = \frac{\#DNM\ in\ probands\ /\ \#probands}{\#\ DNM\ in\ siblings\ /\ \#\ siblings}$$

To test for significance, we compared the counts DNMs in the probands to the counts in their siblings in `R`:

```
binom.test(x= #DNMs_in_probands, n= (#DNMs_in_probands +
#DNMs_in_siblings), p= #probands/(#probands + #siblings),
alternative="two.sided")
```

We note that the measure used to test the relative enrichment in the binomial test is the ***fraction*** of DNMs in ***probands*** among ***all*** DNMs.

$$Fraction = \frac{\#DNM\ in\ probands}{\#\ DNM\ in\ siblings} = \frac{\#DNM\ in\ probands}{\#\ Total\ DNMs - \#\ DNMs\ in\ probands}$$

It is different from the DNM ***rate ratio*** (a risk ratio) between the ***probands*** and ***siblings***.

$$Rate\ Ratio = \frac{\#DNM\ in\ probands\ *\ \#\ siblings}{\#\ DNM\ in\ siblings\ *\ \#\ probands}$$

Here, we report the enrichment as rate ratios as these are easy to interpret and can be used for DNMs, inherited and case-control variants to express fold-enrichment. It is possible to convert between the rate ratio and fraction as follows:

$$Rate\ Ratio\ = \frac{\#DNM\ in\ probands\ *\ \#\,siblings}{(\#\,Total\ DNMs\ -\ \#\,DNM\ in\ probands)\ *\ \#\,probands}$$

$$Rate\ Ratio = \frac{\#DNM\ in\ probands\ *\ \#\,siblings}{(\#\,Total\ DNMs\ *\ \#\,probands)\ -\ (\#\,DNM\ in\ probands\ *\ \#\,probands)}$$

$$Rate\ Ratio = \frac{(\#DNM\ in\ probands\ /\ \#\,Total\ DNMs)\ *\ \#\,siblings}{\#\,probands\ -\ (\#\,DNM\ in\ probands/\ \#\,Total\ DNMs)\ *\ \#\,probands}$$

$$Rate\ Ratio\ = \frac{(\#DNM\ in\ probands\ /\ \#\,Total\ DNMs)\ *\ \#\,siblings}{(1\ -\ (\#\,DNM\ in\ probands/\ \#\,Total\ DNMs)\ )\ *\ \#\,probands}$$

$$Rate\ Ratio\ = \frac{Fraction\ *\ \#\,siblings}{(1\ -\ Fraction\ )\ *\ \#\,probands}$$

Although we calculated the rate ratios directly from the observed counts, the above equation is useful for estimating the confidence intervals with the same test used to calculate the *p*-values; the binomial test returns the upper and lower bounds of the 95% CI of the observed fraction. We converted these bounds to equivalent rate ratios by plugging them into this formula:

$$DNM\ ratio\ CI = \frac{Fraction\ CI_{binomial\ test}\ *\ \#\,siblings}{(1\ -\ Fraction\ CI_{binomial\ test})\ *\ \#\,probands}$$

##### 4.2.2. Over-transmission

To assess over-transmission in the probands, we calculated the ratio between transmitted untransmitted parental alleles:

$$Transmitted/\ Untransmitted\ ratio = \frac{\#\,parental\ alleles\ transmitted\ to\ the\ probands}{\#\,untransmitted\ parental\ alleles}$$

We then compared the significance of the difference in the counts of transmitted and untransmitted alleles:

```
binom.test(
x=#variants_transmitted_to_probands,
n=#variants_transmitted_to_probands + #untransmitted_variants,
p=0.5,
alternative=”two.sided”)
```

We scaled the 95% CIs from the binomial test as follows:

$$Tr./\,Ut.ratio\ CI\ = \frac{Fraction\ CI_{binomial\ test}}{1\ -\ Fraction\ CI_{binomial\ test}}$$

##### 4.2.3. Case-control comparisons

Variants were compared between cases and controls in a similar manner to DNMs, i.e., using the sample size to derive the expected fraction of variants in the cases. These case-control comparisons are more sensitive to differences in population architecture than comparisons of *de novo* mutations and transmitted-untransmitted alleles. When testing the enrichment of ultra-rare variants in cases versus controls in a regression framework, synonymous variant counts can be used as a covariate along with principal components. When using a binomial test, the expected variant rates can be adjusted for population differences by using the synonymous variants. Therefore, the damaging variant comparisons were additionally adjusted by the synonymous variant counts as follows:

$$Case/Control\ rate\ ratio = \frac{\#\ damaging\ variants\ in\ cases\ /\ \#\ synonymous\ variants\ in\ cases}{\#\ damaging\ variants\ in\ controls\ /\ \#\ synonymous\ variants\ in\ controls}$$

```
binom.test(x= #damaging_in_cases, n= (#damaging_in_cases +
#damaging_in_ctrls), p= #syn_in_cases/(#syn_in_cases +
#syn_in_ctrls), alternative=”two.sided”)
```

$$Case/Ctrl\ CI = \frac{Fraction\ CI_{binomial\ test}\ *\ \#synonymous\ in\ controls}{(1\ -\ Fraction\ CI_{binomial\ test})\ *\ \#synonymous\ in\ cases}$$

#### 4.3. Sex differences in enrichment

Sex differences were assessed by comparing autistic females to autistic males in the same manner as we compared autistic individuals to their siblings. Rate ratios > 1 indicate higher variant rates in autistic females compared to autistic males. To test for significant differences in de novo mutation counts, we compared the DNM counts in females and males:

```
binom.test(x= #DNMs_in_females,
n= (#DNMs_in_males + #DNMs_in_females),
p= #females/(#males + #females), alternative=”two.sided”)
```

$$DNM\ ratio\ CI = \frac{Fraction\ CI_{binomial\ test}\ *\ \#\ Females}{(1\ -\ Fraction\ CI_{binomial\ test})\ *\ \#\ Males}$$

For inherited variants, we compared the counts of parental alleles transmitted to females and males:

```
binom.test(x=#variants_transmitted_to_females,
n= #variants_transmitted_to_males_or_females,
p= #all_parental_alleles_in_females/#all_parental_alleles,
alternative="two.sided")
```

$$Tr./Ut.ratio\ CI = \frac{Fraction\ CI_{binomial\ test}\ *\ \#\ parental\ alleles\ in\ females}{(1\ -\ Fraction\ CI_{binomial\ test}\ )\ *\ \#\ parental\ alleles\ in\ males}$$

Case-control variants were tested in the same manner as DNMs.

#### 4.4. Meta-analysis between ASC & SPARK cohorts

We performed a fixed-effect inverse-variance-weighted meta-analysis of the risk ratios (rate ratios or transmitted/untransmitted ratios) in ASC & SPARK using `metagen` function from `meta` package[17] in `R`:

```
metagen(sm = "RR",
      fixed = TRUE,
      studlab = Cohort,
      TE = Risk_Ratio,
      level.ci = 0.95,
      lower = Risk_Ratio_CIL,
      upper = Risk_Ratio_CIU,
      pval = Risk_Ratio_Pval,
      method.tau = "PM")
```

#### 4.5. Correcting for multiple testing

We adjusted the p-values from each experiment (e.g. 54 tests when performing exome-wide enrichment testing) for the Family-wise (Experiment-wise) Error Rate using Bonferroni correction. We also performed more lenient adjustment for False Discovery Rate using Benjamini-Hochberg method. Both were performed in R:

```
      p.adjust(method='bonferroni')
```

```
p.adjust(method='BH')
```

### 5. Measuring effect sizes on the liability scale

We compared the change in trait liability for each variant class to reflect the deviation from population mean liability that would result from carrying a variant in that class. The relative differences in variant frequency between the probands and siblings was converted to Z scores on the liability scale, assuming the liability is normally distributed and centered around zero in the population. The mathematical derivation is explained in Ref[16]. Here, we summarize the concept behind this procedure for those less familiar with the statistical concepts:

- In a Liability Threshold Model assuming additive genetic risk that is normally distributed in the population, the threshold is the point that forms the boundary of an area under the normal distribution curve that is equivalent to the trait prevalence (Figure S4). For example, an autism prevalence of 0.025 indicates that the distance between the population mean risk and the threshold is 1.96 standardized units, putting 2.5% of the population in the right tail.
- We can work out the distance (in standardized units) between the population mean (zero, given how the model is defined) and the threshold using the inverse of the standard normal cumulative distribution function ($\Phi^{-1}$), or '*norminv*' in short. The threshold in the population equals the average liability in the population (i.e., zero) + *norminv*(Prevalence) as shown in Figure S4. The inverse normal function $\Phi^{-1}$ is implemented in `R` using the function `qnorm`, which returns the lower tail by default, whereas the prevalence reflects the upper tail (fraction of individuals *above* the threshold). We can get the upper tail by passing an argument to the function ('`qnorm(Prevalence,lower.tail=FALSE)`) or simply by using the complement of the prevalence, `qnorm(1 - Prevalence)`.
- Now we move to the carriers of a certain class of variants, for instance damaging PTVs. The liability in this sub-population can also be approximated by a normal distribution, and the mean of this distribution will be the *average PTV liability.* This average variant liability is a measure of the average effect size of PTVs on the liability scale. PTV carriers who are autistic will form the upper tail above a certain threshold, and non-autistic PTV carriers will be in the lower tail (Figure S4).
- Whereas the threshold in the population liability distribution reflects the prevalence in the population, the threshold in the PTV liability distribution will reflect autism *prevalence among PTV carriers* in the general population. In accordance with the original derivation in [17], we refer to this fraction as the 'penetrance'.

- Similar to the prevalence, the penetrance is a cumulative density and as such can be converted to equivalent standardized units in the PTV liability distribution using the inverse normal cumulative density distribution function ($\Phi^{-1}$). The threshold here equals the (unknown) average PTV liability + *norminv*(Penetrance). We can get this is `R` using `qnorm(1 - Penetrance)`.
- By leveraging the fact that the threshold in both distributions is the same, we can now estimate the distance between the population mean (zero) and the (unknown) mean protein-truncating variant liability. As shown in Figure S4, we can write:

  threshold = `qnorm(1 - Prevalence)` = *average PTV liability* + `qnorm(1 - Penetrance)`
- The prevalence in the general population is a known parameter. Autism is diagnosed in 1 in 40 male individuals (a prevalence of 2.5% in males), with a 1:4 male-to-female ratio (prevalence in females = 0.25 x 2.5% = 0.625%). Previous estimates suggested that about one third of autistic individuals in the population have profound deficits with cognitive impairment [18,19]. Among 11,630 autistic individuals in SPARK who could be classified, ~ 36% fell in the autism with motor or cognitive impairment group (35% in males & 40% in females). For estimating liability, we scaled the sex-specific population prevalence using these percentages, i.e., using a prevalence estimate of 0.88% in males (35% x 2.5%) and 0.25% in females (40% x 0.625%) for liability calculations in the autism with motor or cognitive impairment group; for the autism without motor or cognitive impairment group, we used a prevalence of 1.62% in males (2.5% - 0.88%) and 0.38% in females (0.625% - 0.25%). For directly comparing those with motor or cognitive impairment to those without these co-occurring difficulties, we used a prevalence of 0.4 in females and 0.35 in males.
- The prevalence among PTV carriers, on the other hand, is not known. We can, however, convert the PTV carrier rates in the study cohort to population estimates, and convert these population estimates to penetrance estimates. Specifically, the penetrance is the ratio between PTV rate among autistic individuals and the overall frequency of PTVs in the population. The overall population frequency of PTVs can, in turn, be estimated from the study cohort, namely by summing the frequency of PTVs in the probands weighed (multiplied) by their fraction in the population (trait prevalence) and the frequency of PTVs in the siblings weighed by their relative fraction as well (1 minus the trait prevalence).
- Now that we have the prevalence estimates both in the general population and among PTV carriers (penetrance), we estimate the average PTV liability as `qnorm(1 - Prevalence)- qnorm(1 - Penetrance)`.

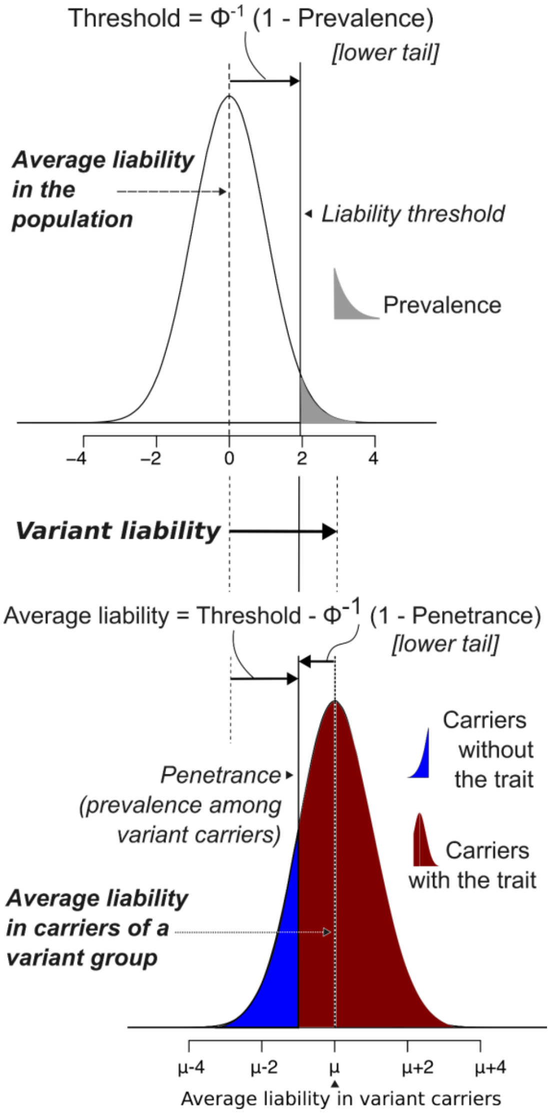


Figure S4: Estimating rare variant liability from population prevalence and carrier rates of variant groups.

The Liability Threshold Model postulates that risk factors act additively and underlie a normally distributed liability distribution in the general population (top), where the threshold determines the trait prevalence. When examining carriers of a group of rare variants (bottom), the average liability will reflect the effect size of these variants. If the prevalence of the trait among rare variant carriers is known, the two distributions can be compared to get an estimate of the distance between their means. See the ‘Variant liability’ section for a detailed description.

- The liability stratified by sex was measured in `R` using these formulas:

$$Rate_{Population} = Prevalence * Rate_{Probands} + (1 - Prevalence) * Rate_{Siblings}$$

$$Penetrance = \frac{Rate_{Probands}}{Rate_{Population}} * Prevalence$$

$$Z = qnorm(1 - Prevalence) - qnorm(1 - Penetrance)$$

- The *p*-values were those obtained from the binomial tests detailed above. The standard error, and then the confidence intervals around Z, were estimated from these *p*-values:

$$Q = qnorm(1 - \frac{Binomial\ test\ P\ value}{2})$$

$$SE = \frac{abs(Z)}{Q}$$

$$95\%\ CI = Z \pm (1.96 * SE)$$

- The difference between males and females liability estimates was measured as follows:

$$Z_{Difference} = Z_{Females} - Z_{Males}$$

$$SE_{Difference} = sqrt(SE^2{}_{Females} + SE^2{}_{Males})$$

$$Q_{Difference} = \frac{abs(Z_{Difference})}{SE_{Difference}}$$

$$P_{Difference} = 2 * (1 - pnorm(Q_{Difference}))$$

- The inverse-variance-weighted average from both sexes and its confidence interval was measured as follows:

$$Z_{Both} = (\frac{Z_{Females}}{SE^2{}_{Females}} + \frac{Z_{Males}}{SE^2{}_{Males}}) \div (\frac{1}{SE^2{}_{Females}} + \frac{1}{SE^2{}_{Males}})$$

$$SE_{Both} = 1 \div sqrt(\frac{1}{SE^2{}_{Females}} + \frac{1}{SE^2{}_{Males}})$$

$$95\%\ CI_{Both} = Z_{Both} \pm (1.96 * SE_{Both})$$

- The meta-analysis *p*-value was obtained using Fisher's method implemented in the `metap` package[17]:

$$P_{Both} = metap::sumlog(P_{Females}, P_{Males})$$

- The *p*-values for each experiment were corrected for Family-wise Error Rate (Bonferroni correction) and False Discovery Rate (Benjamini-Hochberg correction), as shown for the risk ratios.

- We did an additional analysis in which we removed a set of high confidence autism predisposition genes to assess the residual exome-wide liability in unknown risk genes. The Simons Foundation Autism Research Initiative (SFARI) database provides curated scores for 1,172 genes that reflect the strength of evidence linking each gene to autism risk. There are 3,58 autosomal and 47 X-linked SFARI Category 1 (high-confidence) & S (syndromic) genes with strong evidence of association with autism (354 autosomal genes included in the current analysis cohort after QC). We tested the enrichment of DNMs and rare inherited variants after removing these genes as well as in this gene set only.

### 6. Gene set enrichment

#### 6.1. Genes with sex-biased expression

Genes with sex-biased expression in the fetal cortex (FDR < 0.1) were obtained from Supplementary Table S1 of a study by O'Brien and colleagues.[20] Autosomal male-biased genes (n=856) were defined as those with fold-difference (Male/Female) > 1, and female-biased genes (n=794) as those with fold-difference < 1. Genes with significant sex-biased expression in the adult human cortex (FDR < 0.05) were obtained from Supplementary Table S1 of a recent study by Fass and colleagues.[21] Autosomal male-biased genes (n=303) were defined as those with fold-change in the cortex (logFC_Cortex < 1), and female-biased genes (n=426) as those with fold-difference > 1. We note that the genes were limited to autosomal genes annotated in our dataset; the FDR cutoffs were those used by the authors in the source publications.

#### 6.2. Gene set enrichment versus matched genes

For each tested gene set, we selected a matched set (see 6.3) and counted DNMs, transmitted and untransmitted variants; we repeated this procedure 10,000 times with replacement and took the average ratio (rate ratio between DNM counts in probands and siblings or transmitted to untransmitted ratio in the probands); we then used this ratio as the expected ratio in a binomial test as described above. Specifically, we tested the difference between the rate of DNMs between probands and siblings against the permutation-averaged expected ratio for this gene set (instead of the sample size ratio used in the exome-wide analyses), and similarly tested rare variant over-transmission against the permutation-averaged expected transmitted-to-untransmitted ratio for the given gene set (instead of 0.5 as used in the exome-wide analysis). We also used the average variant rates across these 10,000 permutations instead of the rate in siblings to estimate the variant liability attributed to a gene set in excess of what is expected for matched genes.

#### 6.3. Selecting random sets of matching genes

We followed a procedure similar to that previously used by Ouwenga and Dougherty[22] to select sets of control genes for gene set burden analysis. We used a multi-dimensional kernel density estimator (KDE) from the `ks` package[23] in `R` to select these control genes. First, we created a table of features (coding length, brain expression level, and LOEUF bins) and used it to build a 3D KDE. Next, we used the test gene set to build a density distribution and evaluated the remaining genes (not in the gene set) on this distribution. We then evaluated these remaining genes on a density distribution built using all genes rather than the gene set genes. The ratio of the two estimates was then used as a sampling weight. Here we show the `R` code used to implement this:

```
kde_set_genes = ks::kde(
                x = features_set_genes,
                eval.points = features_remaining_genes
                )
                # feature_x_genes: table gene x length, expr., loeuf
kde_all_genes = ks::kde(
                x = features_all_genes,
                eval.points = features_remaining_genes
                )
kde_weights =  kde_set_genes$estimate ÷ kde_all_genes$estimate
genes_random_idx = sample(
                1:length(remaining_genes),
                length(set_genes),
                prob =kde_weights
                )
genes_randoms = remaining_genes[genes_random_idx]
```

### 7. References

1. Zhou, X. *et al.* Integrating de novo and inherited variants in 42,607 autism cases identifies mutations in new moderate-risk genes. *Nat. Genet.* **54**, 1305–1319 (2022).
2. Foster, I. Globus Online: Accelerating and Democratizing Science through Cloud-Based Services. *IEEE Internet Comput.* **15**, 70–73 (2011).
3. Chorbadjiev, L. *et al. The Genotype and Phenotypes in Families (GPF) Platform Manages the Large and Complex Data at SFARI*. http://biorxiv.org/lookup/doi/10.1101/2024.02.08.579330 (2024) doi:10.1101/2024.02.08.579330.
4. McLaren, W. *et al.* The Ensembl Variant Effect Predictor. *Genome Biol.* **17**, 122 (2016).
5. Genome Aggregation Database Consortium *et al.* The mutational constraint spectrum quantified from variation in 141,456 humans. *Nature* **581**, 434–443 (2020).
6. The 1000 Genomes Project Consortium. A global reference for human genetic variation. *Nature* **526**, 68–74 (2015).
7. Chang, C. C. *et al.* Second-generation PLINK: rising to the challenge of larger and richer datasets. *GigaScience* **4**, 7 (2015).
8. Manichaikul, A. *et al.* Robust relationship inference in genome-wide association studies. *Bioinformatics* **26**, 2867–2873 (2010).
9. Van der Auwera, G. A. *et al.* From FastQ Data to High-Confidence Variant Calls: The Genome Analysis Toolkit Best Practices Pipeline: The Genome Analysis Toolkit Best Practices Pipeline. in *Current Protocols in Bioinformatics* (eds. Bateman, A., Pearson, W. R., Stein, L. D., Stormo, G. D. & Yates, J. R.) 11.10.1-11.10.33 (John Wiley & Sons, Inc., Hoboken, NJ, USA, 2013).
10. Wright, M. N. & Ziegler, A. **ranger** : A Fast Implementation of Random Forests for High Dimensional Data in *C++* and *R*. *J. Stat. Softw.* **77**, (2017).
11. Kuhn, M. Building Predictive Models in *R* Using the **caret** Package. *J. Stat. Softw.* **28**,

(2008).

12. Fu, J. M. *et al.* Rare coding variation provides insight into the genetic architecture and phenotypic context of autism. *Nat. Genet.* **54**, 1320–1331 (2022).
13. Hail Team. Hail. (2022).
14. Danecek, P. *et al.* Twelve years of SAMtools and BCFtools. *GigaScience* **10**, giab008 (2021).
15. Satterstrom, F. K. *et al.* Autism spectrum disorder and attention deficit hyperactivity disorder have a similar burden of rare protein-truncating variants. *Nat. Neurosci.* **22**, 1961–1965 (2019).
16. Satterstrom, F. K. *et al.* Large-Scale Exome Sequencing Study Implicates Both Developmental and Functional Changes in the Neurobiology of Autism. *Cell* **180**, 568-584.e23 (2020).
17. Balduzzi, S., Rücker, G. & Schwarzer, G. How to perform a meta-analysis with R: a practical tutorial. *Evid. Based Ment. Health* **22**, 153–160 (2019).
18. Dougherty, J. D. *et al.* Can the “female protective effect” liability threshold model explain sex differences in autism spectrum disorder? *Neuron* **110**, 3243–3262 (2022).
19. Zeidan, J. *et al.* Global prevalence of autism: A systematic review update. *Autism Res.* **15**, 778–790 (2022).
20. O’Brien, H. E. *et al. Sex Differences in Gene Expression in the Human Fetal Brain*. http://biorxiv.org/lookup/doi/10.1101/483636 (2018) doi:10.1101/483636.
21. Fass, S. B. *et al. Relationship between Sex Biases in Gene Expression and Sex Biases in Autism and Alzheimer’s Disease*. http://medrxiv.org/lookup/doi/10.1101/2023.08.29.23294773 (2023) doi:10.1101/2023.08.29.23294773.
22. Ouwenga, R. L. & Dougherty, J. Fmrp targets or not: long, highly brain-expressed genes tend to be implicated in autism and brain disorders. *Mol. Autism* **6**, 16 (2015).
23. Duong, T. ks: Kernel Smoothing. R package version 1.14.0. (2022).

### Supplementary Results

#### 1. DNM and inherited variant rates

The *de novo* and rare inherited variant rates observed in the siblings in the current SPARK release (iWES2) were comparable to, albeit slightly lower than, the variant rates seen in the ASC cohort (Figure S5). Small differences are expected given the different *de novo* calling pipelines (see section 2 of the Supplementary Methods). The rare inherited variant rates (MAF<0.1%) in SPARK iWES2 were higher than those seen in the ASC cohort and this difference was most prominent in damaging missense variants. Average rare inherited variant counts are sensitive to population differences, partly because different ancestral groups differ in their demographic histories and hence allele frequency spectra, and also because of how the variants are filtered based on in-sample frequencies. While the ASC cohort is predominantly European (composed of the Simons Simplex Collection and other smaller cohorts), SPARK has more diversity, being only 73% European ancestry in iWES2 (see Figure S1 in the Supplementary Methods). To have a more informative comparison unconfounded by potential artifactual differences in processing, we explored the variant rates in the previously analyzed SPARK cohort (Pilot and first sequencing wave WES1; 78% European ancestry), which was processed using the same pipeline as the ASC. The rare inherited variant rates in SPARK Pilot/WES1 were also higher than in ASC (Figure S1). This suggests that these differences in rare variant rates are a reflection of the diverse genetic ancestry of SPARK samples more so than mere technical differences. Ultra-rare variant rates (i.e. seen in one parent in the dataset, absent from gnomAD) in the remaining cohorts (one sequenced parent, without sequenced parents) are shown in Figure S6.

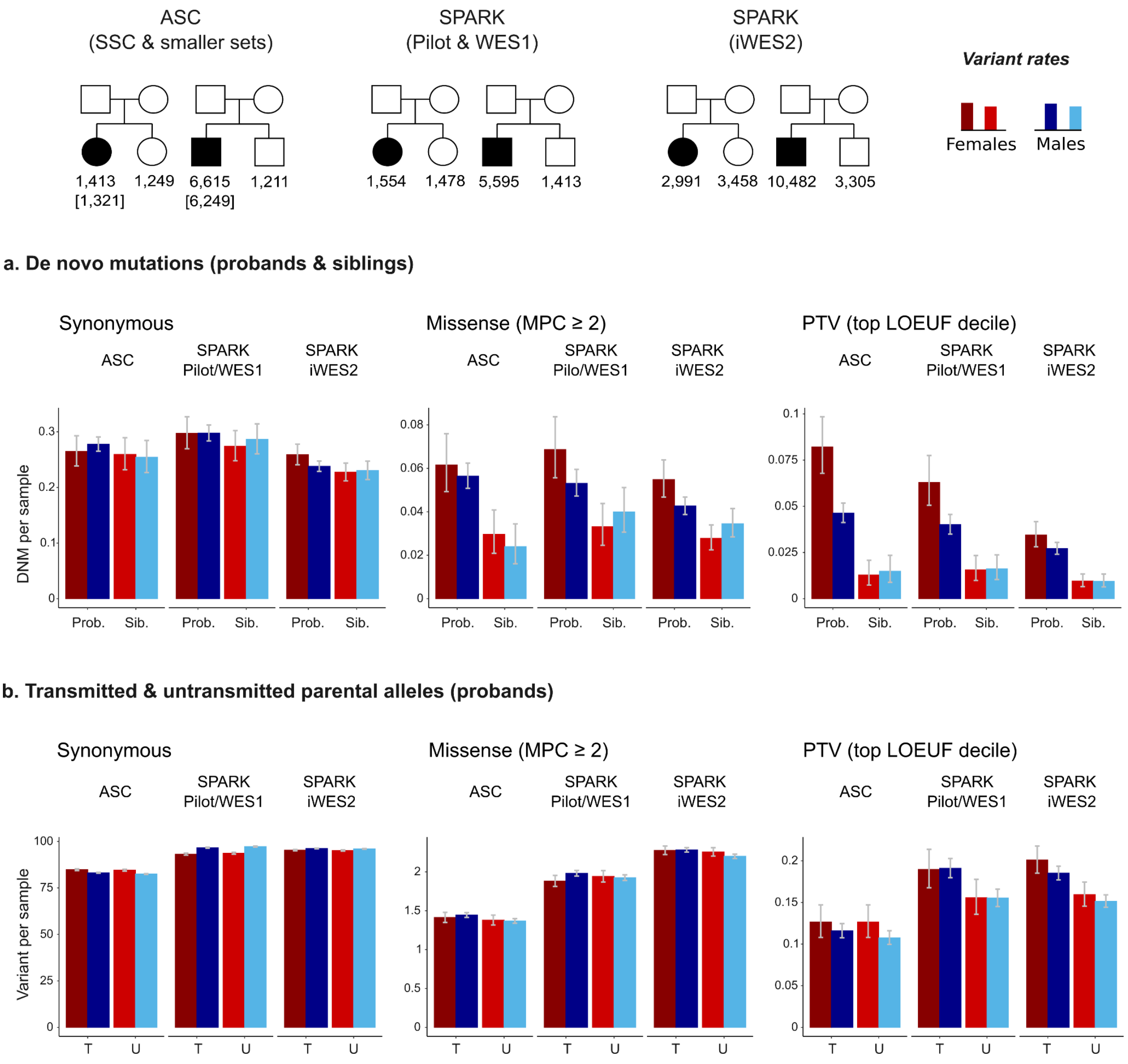


Figure S5: Rare *de novo* and inherited variant rates in the ASC & SPARK trio-sequenced cohorts.

**a**, *De novo* mutation rates in the probands (prob.) and siblings (sib.) in the Autism Sequencing Consortium (ASC) cohort and the Simons Foundation Powering Autism Research for Knowledge (SPARK) cohort. **b**, Counts of transmitted (T) and untransmitted (U) parental alleles at rare variants (MAF<0.1%). The ASC and SPARK iWES2 were used for the enrichment analysis. SPARK Pilot/WES1 is shown for comparison, as it was processed using the same pipeline used for the ASC. The pedigrees at the top show the sample size used for calculating *de novo* rates in each cohort (sample size for inherited variants given between brackets as some individuals in the ASC cohort did not have information on inherited alleles). The error bars indicate the 95% confidence intervals of the carrier rates.

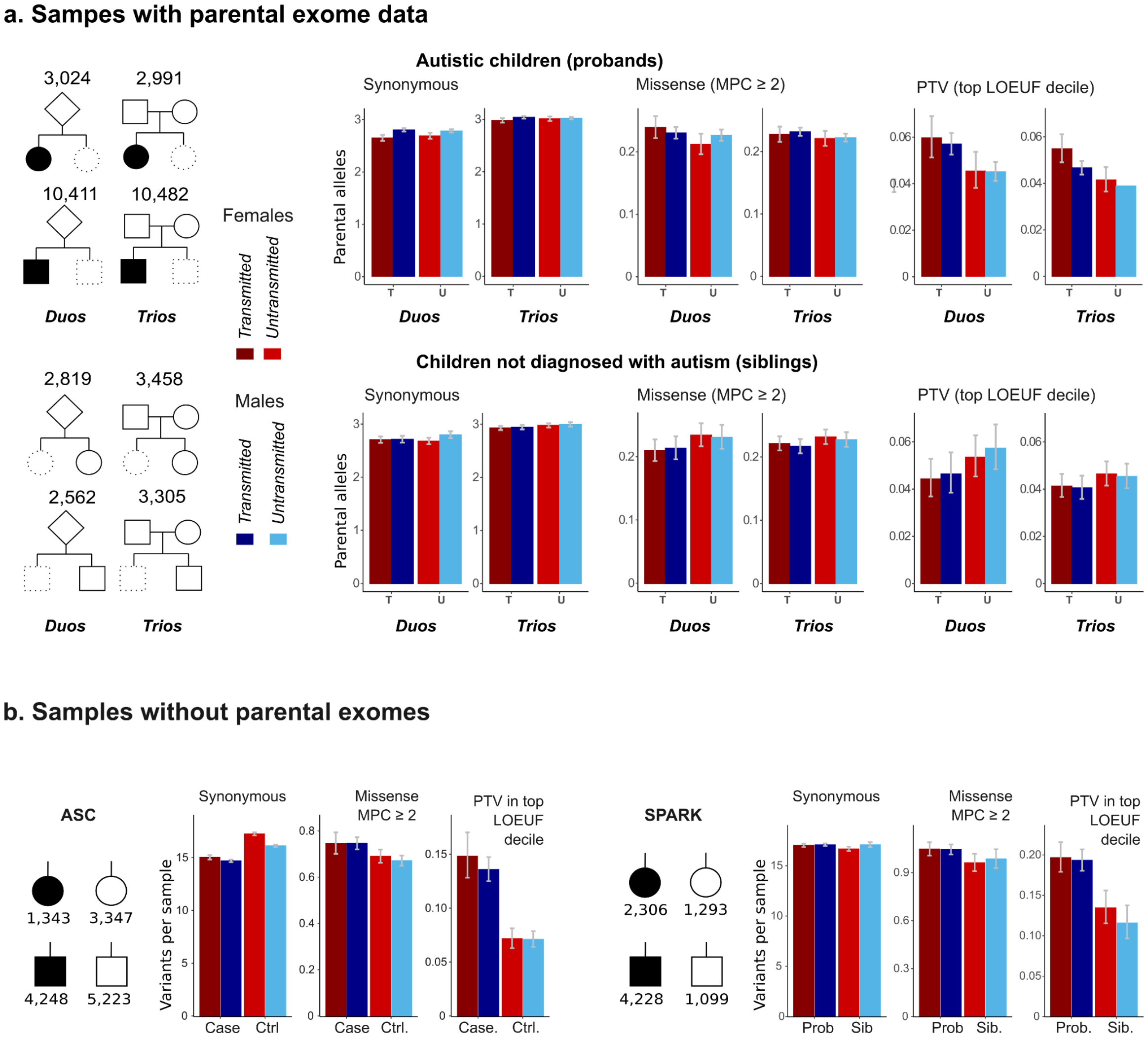


Figure S6 Ultra-rare inherited and case-control variant rates in the remaining cohorts.

**a**, The transmission rates of ultra-rare parental alleles (seen in one parent and not in gnomAD) in probands and siblings with exome data from one parent in the Simons Foundation Powering Autism Research for Knowledge (SPARK) cohort. Transmission rates of ultra-rare variants in trio-sequenced probands and siblings are shown for comparison. **b**, Ultra-rare variants (allele frequency < 0.005%) in autism cases and controls from the Autism Sequencing Consortium (ASC) case-control cohorts and probands and siblings from SPARK who did not have parental sequence data. The pedigrees show the sex-stratified sample size in each cohort. The error bars indicate the 95% confidence intervals of the carrier rates.

### 2. Sex differences in autism likelihood conferred by exome-wide rare variants

Here we describe in more detail the results shown in Figure 1.

#### 2.1. *De novo* mutations

##### 2.1.1 Damaging protein-truncating DNMs

Damaging protein-truncating DNM rates in autistic males were three times higher than sex-matched siblings, and were comparable in SPARK and ASC; we found a 2.9 fold-enrichment in SPARK (95% CI = 2 - 4.3; p = $2.1x10^{-10}$; Bonferroni-corrected p =$1.1x10^{-8}$) and a 3.1 fold-enrichment in ASC (95% CI = 1.9 - 5.3 ; p = $5.1x10^{-8}$; Bonferroni-corrected p = $2.8x10^{-6}$). The enrichment in females *versus* sex-matched siblings was slightly stronger in SPARK (risk ratio = 3.6; 95% CI = 2.4 - 5.5; p = $4.8x10^{-12}$; Bonferroni-corrected p = $2.6x10^{-10}$) and much more pronounced in the ASC (risk ratio = 6.4, 95% CI =3.8 - 11.6 ; p = $1.7x10^{-17}$; Bonferroni-corrected p = $9.4x10^{-16}$). The most notable sex difference was seen in *de novo* damaging PTV rates in the ASC cohort, where DNMs rates in autistic females were 1.8 times higher than autistic males (95% CI = 1.4 - 2.2; p = $5.1x10^{-7}$; Bonferroni-corrected p = $2.7x10^{-5}$). The difference in SPARK was lower (risk ratio = 1.3; 95% CI = 1 - 1.6) and significant only before correction for multiple testing (p = 0.04).

On the liability scale, *de novo* damaging PTVs increased the liability by 0.5 standard deviation units in SPARK - similarly for both sexes (95% CI: 0.3 - 0.6 ; Bonferroni-corrected p < $3x10^{-9}$). In the ASC cohort, they also increased the liability by 0.5 units in males (95% CI = 0.3 - 0.7; Bonferroni-corrected p = $3x10^{-7}$), and caused a larger shift in females (Z = 0.75; 95% CI = 0.6 - 0.9 ; Bonferroni-corrected p = $4.7x10^{-16}$), albeit not significantly different from males (p = 0.09). SFARI genes showed the same general patterns of protein-truncating DNM enrichment with larger effect sizes. Similar patterns were observed when removing these known autism predisposition genes, with considerably lower effect sizes (Figure S7a). We note here that the discovery of SFARI genes was based, in part, on these datasets.

##### 2.1.2 Damaging missense DNMs

Damaging missense mutations showed comparable enrichment in females *versus* siblings in SPARK (risk ratio = 2; 95% CI = 1.5 - 2.6 ; p = $7x10^{-8}$; Bonferroni-corrected p = $3.8x10^{-6}$) and ASC (risk ratio = 2.1; 95% CI = 1.4 - 3.1; p = 0.00013; Bonferroni-corrected p = 0.0075). The enrichment in males was slightly higher in the ASC (risk ratio = 2.4; 95% CI =

1.6 - 3.6 ; p = $8 \times 10^{-7}$; Bonferroni-corrected p = $4.3 \times 10^{-5}$) but considerably lower in SPARK (risk ratio = 1.2; 95% CI = 1 - 1.5 ; p = 0.04; not significant after FDR or Bonferroni correction). In terms of direct comparisons, damaging missense DNMs were slightly more frequent in autistic females *versus* autistic males in SPARK (risk ratio = 1.3 , 1.1 - 1.5; p = 0.0073; FDR-adjusted p = 0.017; FDR-adjusted p = ; Bonferroni-corrected p = 0.39) but did not show a significant sex difference in ASC (p = 0.45).

Females in both ASC and SPARK had comparable variant liability attributed to damaging missense DNMs (ASC = 0.27; 95% CI = 0.13 - 0.4; SPARK = 0.25; 95% CI = 0.16 - 0.24; Bonferroni-corrected p < 0.00044). In males, these missense DNMs showed different estimates between ASC (Z = 0.38; 95% CI = 0.23 - 0.53) and SPARK (0.09 units; 95% CI = 0.003 - 0.18), and this difference between cohorts was significant ($Z_{Difference}$ = 0.29; 95% CI: 0.12 - 0.46; p = 0.0011). In terms of sex differences, damaging missense DNMs had similar average liability in ASC but significantly lower liability in males in SPARK ($Z_{Difference}$ = -0.16; 95% CI: -0.28 - 0.034; p = 0.013; FDR-adjusted p = 0.031; Bonferroni-corrected p = 0.70). However, these sex-stratified estimates were more congruent between males and females when examining ultra-rare DNMs (see 2.2 below), suggesting that the effect sizes are indeed similar. This is in line with the findings from meta-analysis, where the estimates were not significantly different between males and females (Figure 1b). As seen with protein-truncating PTVs, damaging missense DNMs in SFARI genes and the remaining autosomal genes showed similar patterns compared to that obtained with exome-wide analysis, with larger and smaller effect sizes, respectively (Figure S7).

##### 2.1.3 Synonymous DNMs

We note that rare synonymous DNMs showed a small but significant liability in autistic females in SPARK (Z = 0.046, 95% CI = 0.01 - 0.082; p = 0.011; FDR-adjusted p = 0.029). This likely reflects the higher genetic diversity in autistic females (72% individuals of European genetic ancestry) compared to sex-matched siblings (75% individuals of European genetic ancestry). This imbalance, however, did not persist after meta-analyzing the two cohorts (Z = 0.036; 95% CI = 0.0055 - 0.067; p = 0.052). A stringently-defined set of ultra-rare DNMs in SPARK (allele frequency < 0.005%) showed well-balanced synonymous mutation counts between female probands and sex-matched siblings while having the same protein-truncating DNM liability (see 2.1.4 and Figure S8 below). Therefore, it is unlikely that the meta-analyzed estimates of damaging DNM liability presented in Figure 1 are biased.

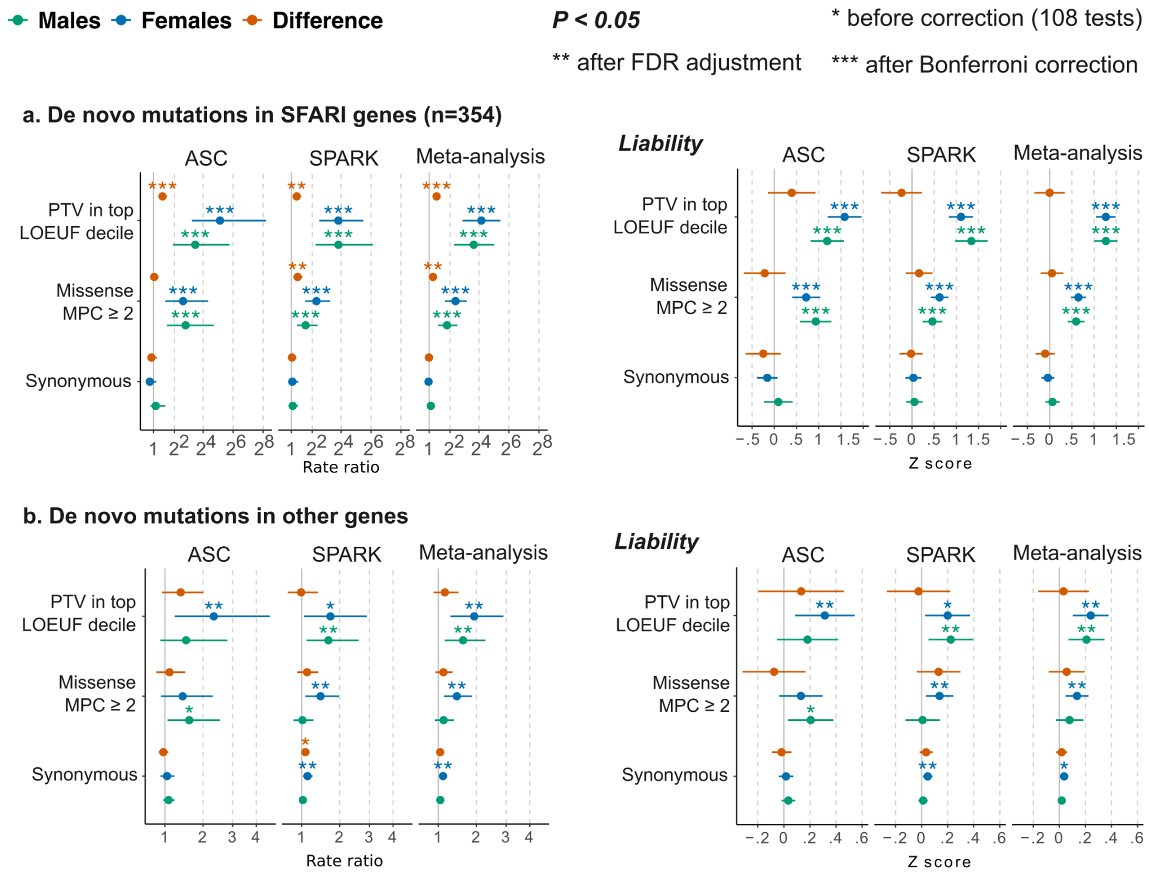


Figure S7. Enrichment of *de novo* mutations in SFARI genes versus all other genes.

Sex-stratified *de novo* mutation rate ratios (left) and liability (right) (see Methods) in SFARI genes (**a**) and all other genes (**b**). For sex differences, a rate ratio > 1 indicates that females show a higher enrichment; a Z score > 0 indicates that females show a higher effect size on the liability scale.Error bars show 95% confidence intervals. Similar to Figure 1b.

##### 2.1.4 Ultra-rare *de novo* mutations in SPARK

Almost all protein-truncating DNMs and most damaging missense DNMs were ultra-rare, whereas only half of the synonymous variants were in this group (Figure S8). Consequently, the liability of ultra-rare and rare protein-truncating DNMs were similar; the liability of ultra-rare damaging missense DNMs in males (Z = 0.16; 95% CI = 0.06 - 0.25; p = 0.0015) was higher than that seen with rare missense DNMs (Z = 0.09; 95% CI = 0.003 - 0.18; p = 0.04). There was no significant sex difference in ultra-rare DNM liability (p = 0.065) in contrast to what is seen with rare DNMs (p = 0.012). Ultra-rare synonyms DNMs were not enriched in the probands *versus* siblings, nor did they show a significant sex difference.

All in all, this analysis showed a stronger enrichment in ultra-rare damaging DNMs along with better-balanced ultra-rare synonymous DNM burden, which suggests that the imbalance in synonymous DNMs seen in the rare variant analysis is not accompanied by an inflation of the estimates for damaging mutations particularly protein-truncating mutations, which showed equal or rather higher risk ratios in ultra-rare DNM compared to rare DNMs.

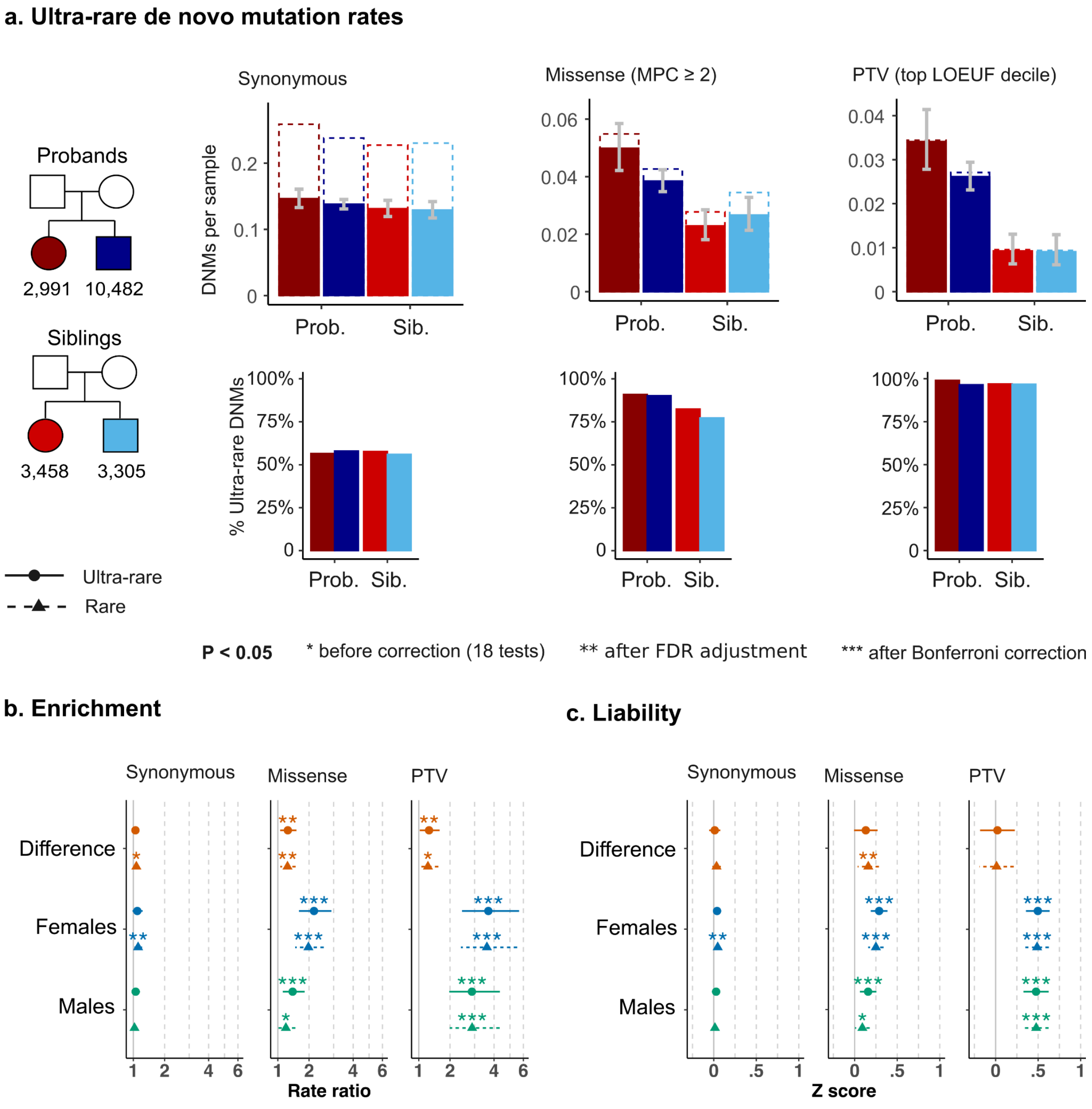


Figure S8. Enrichment of ultra-rare de novo mutations in SPARK trio-sequenced individuals.

**a**, Rates of ultra-rare de novo mutations (DNM) (filled bars) compared to all rare DNMs (dotted lines). Most damaging protein-truncating and missense *de novo* mutations (DNMs) were ultra-rare compared to about half of the synonymous DNMs. **b,** Ultra-rare DNM rate ratios between probands and siblings, compared to rare DNMs. **c**, The liability of ultra-rare and rare DNMs. There was no significant difference in the average liability of ultra-rare synonymous DNM, in contrast to the difference seen in rare DNMs. Error bars show 95% confidence intervals.

#### 2.2. Inherited variants

##### 2.2.1. Damaging protein-truncating variants

Damaging PTVs showed evidence of over-transmission from non-autistic parents to autistic individuals in SPARK only - both to female (transmission ratio = 1.3 ; 95% CI = 1.1 - 1.4; p = 0.00018; Bonferroni-corrected p = 0.0095) and males (T/U = 1.22; 95% CI = 1.14 - 1.3; p = $3x10^{-9}$; Bonferroni-corrected p = $1.6x10^{-7}$). Similar to the observed over-transmission patterns, inherited damaging PTVs conveyed approximately the same liability in males (Z = 0.085 units; 95% CI = 0.057 - 0.11) and females (Z = 0.082; 95% CI = 0.04 - 0.13).

There was still a significant over-transmission and inherited liability in SPARK when excluding SFARI genes, suggesting that a substantial portion of the genes driving this association are not in the SFARI gene set. In particular, females showed significant over-transmission in the remaining genes rather than SFARI genes, whereas males showed significant over-transmission in both. In the ASC, only SFARI genes showed significant PTV over-transmisisonl, both in males and in females. These findings are shown in Figure S9.

Ultra-rare inherited PTVs ascertained in a cohort of autistic probands with sequence data from a single parent in females (T/U = 1.3; 95% CI = 1.05 - 0.65 ; p = 0.018; FDR-adjusted p = 0.14; Bonferroni-corrected p = 0.99) and males (T/U = 1.26; 95% CI = 1.12 - 1.43; p = 0.00016; FDR-adjusted p = 0.0021; Bonferroni-corrected p = 0.0084). Accordingly, these variants conveyed similar liabilities in females (Z = 0.068; 95% CI = 0.017 - 0.18) and males (Z = 0.10; 95% CI = 0.048 - 0.15). The effect size of ultra-rare PTVs were similar in trio-sequenced probands, albeit slightly less prominent in males (Figure S10a).

##### 2.2.2. Damaging missense variants

Damaging missense variants showed comparable over-transmission to autistic males but not to females in both cohorts; the transmission ratios were significantly higher than 1 in autistic males in ASC (T/U = 1.06; 95% CI = 1.02 - 1.09; p = 0.00033; Bonferroni-corrected p = 0.018) and in SPARK (T/U = 1.04; 95% CI = 1.02 - 1.06; p = $7.4x10^{-5}$; Bonferroni-corrected p = 0.004) but not in females in both (T/U in ASC = 1.02; 95% CI = 0.96 - 1.09; p = 0.47; T/U in SPARK = 1.01; 95% CI = 0.98 - 1.04; p = 0.61). Despite this difference in sex-stratified estimates, there was no statistically significant over-transmission in autistic females when compared directly to autistic males (T/U in ASC = 0.99; 95% CI = 0.94 - 1.04; p = 0.58; T/U in SPARK = 0.99; 95% CI = 0.96 - 1.01; p = 0.32).

The cohort-level liability attributed to inherited damaging missense variants mirrored the over-transmission patterns, where it was significant in males in ASC (Z = 0.023; 95% CI = 0.010 - 0.035) and SPARK (Z = ; 95% CI = 0.015 - 0.0077) but not females, without a significant sex difference (p > 0.093). As noted in the main text, this sex difference was nominally significant in the meta-analyzed cohort (Figure 1). The liability attributed to inherited damaging missense variants in SFARI genes did not differ significantly from the population mean. Consequently, the liability in the remaining genes mirrored that seen exome-wide (Figure S9).

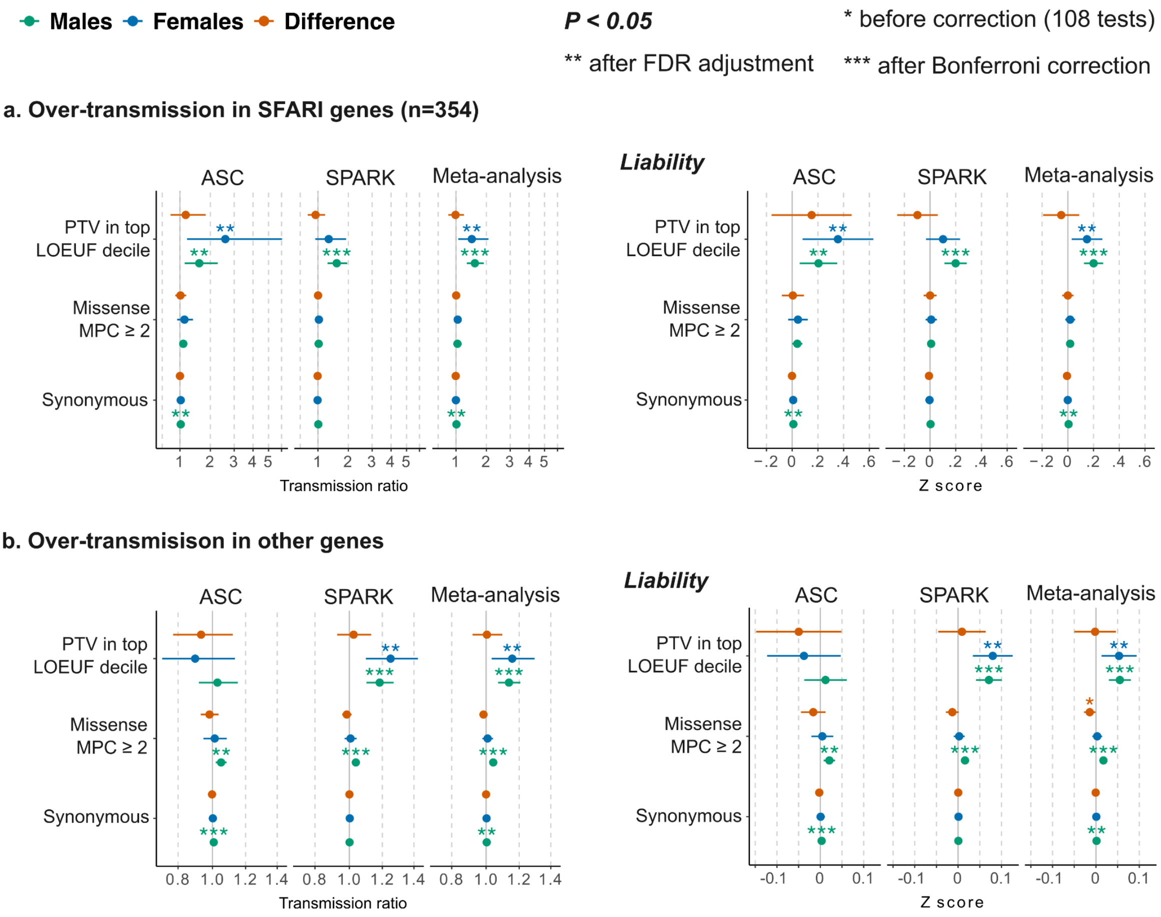


Figure S9. Over-transmission of rare inherited variants in SFARI genes.

Over-transmission (left) and liability (right) of rare inherited variants in SFARI genes (**a**) versus all other genes (**b**) (see Methods). Error bars show 95% confidence intervals. Similar to Figure 1c.

When evaluating ultra-rare variants in the remaining child-parent pairs (Figure S10), damaging missense variants showed over-transmission to females (T/U = 1.13; 95% CI =

1.013 - 1.26; p = 0.028; FDR-adjusted p = 0.19; Bonferroni-corrected p = 1) but not males (T/U = 1.017; 95% CI = 0.096 - 1.077; p = 0.57). This was different from ultra-rare variants in trio-sequenced probands, in whom these variants were over-transmitted to males (T/U = 1.042; 95% CI = 1.0010 - 1.085; p = 0.045; FDR-adjusted p = 0.20; Bonferroni-corrected p = 1) but not females (T/U = 1.030; 95% CI = 0.95 - 1.11; p = 0.46); this was shown above for rare variants in the same individuals. As a consequence, the liability was significantly higher than zero in females in the one cohort (Z = 0.043; 95% CI = 0.0046 - 0.081) and in males in the other  (Z = 0.017; 95% CI = 0.00042 - 0.034), but not significantly higher than zero in a meta-analysis of the two (In males: Z = 0.014; 95% CI = 0.000096 - 0.028; p = 0.12; In females: Z = 0.021 ; 95% CI = -0.00095 - 0.044; p = 0.07). There was no significant sex difference in any of these ultra-rare variant comparisons. Siblings showed nominal under-transmission of PTVs that was not significantly different from zero on the liability scale (Figure S10b).

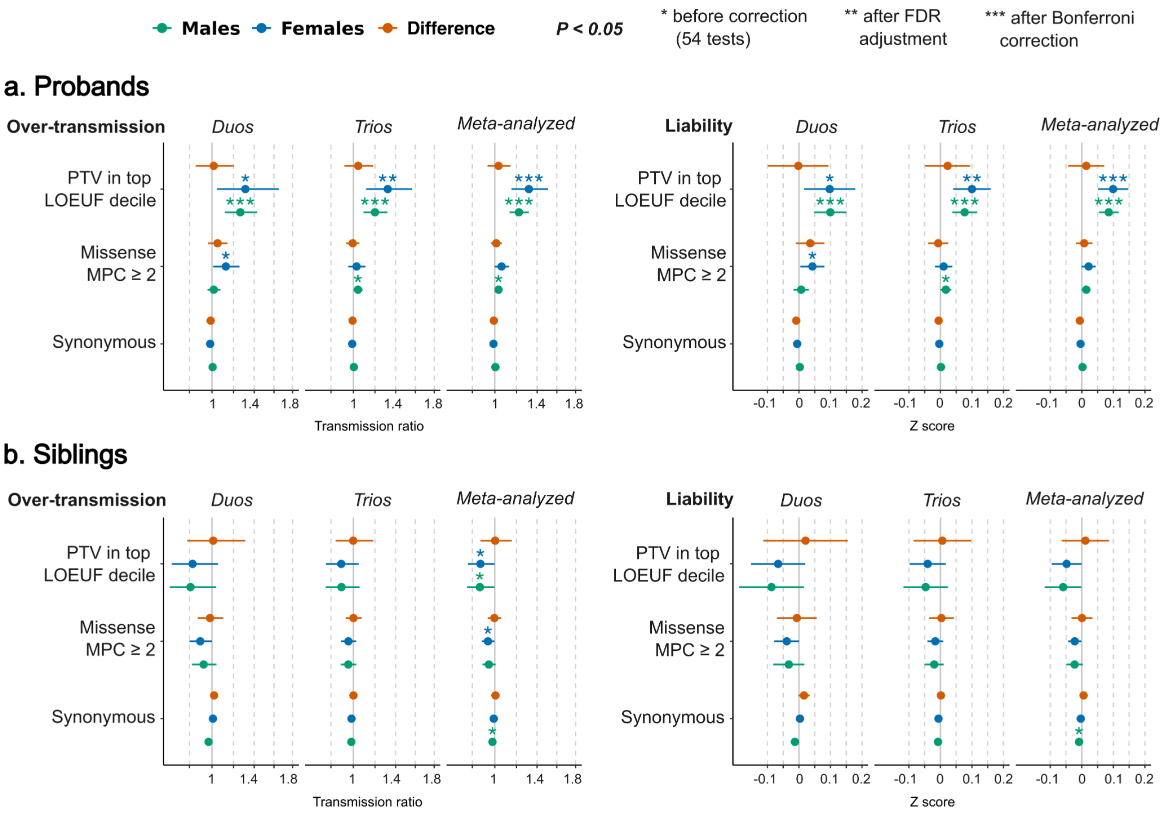


Figure S10. Over-transmission of ultra-rare inherited variants in SPARK.

Over-transmission of ultra-rare variants (i.e. seen in one parent in the dataset, absent from gnomAD) was studied in child-parent pairs with one (duos) or two sequenced parents (trios), then meta-analyzed. This analysis was performed in probands (**a**) and siblings (**b**). Error bars show 95% confidence intervals. See Figure 1c for over-transmission analysis of rare (MAF<0.1%) variants.

##### 2.2.3. Synonymous variants

Inherited parental allele counts in SPARK were not significantly different from untransmitted alleles (Figure 1c). There was an imbalance in transmitted and untransmitted synonymous variants in autistic males in the ASC cohort (Transmitted alleles = 519,579; Untransmited = 515,406; Rate ratio = 1.0081) corresponding to a small effect size on the liability scale (Z = 0.0034; 95% CI = 0.0018 - 0.005; p = $4.12 \times 10^{-5}$; Bonferroni-corrected p = 0.0022). This difference is likely to be due to the greater sensitivity to call rare variants that are transmitted (i.e. occur at least twice in the dataset) than those that are not transmitted. It was persistent after meta-analysis across cohorts but was extremely small (Z = 0.0016; 95% CI = 0.00070 - 0.0026; p = 0.0001; Bonferroni-corrected p = 0.0056) and thus unlikely to have major implications for the key conclusions.

#### 2.3. Ultra-rare variants in cases and controls

Case-control cohorts (including individuals recruited in family-based studies but currently without sequencing data from their parents) form a substantial portion of available autism cohorts. These constitute valuable independent datasets to study sex differences in the enrichment of ultra-rare variants. To leverage these data, we compared the enrichment of ultra-rare variants in 12,125 autistic individuals *versus* 10,962 controls or siblings not diagnosed with autism (Figure S6b). This analysis showed significant enrichment and liability attributed to damaging PTVs, and to a lesser extent damaging missense variants, without significant sex differences (Figure S11). Since the mode of inheritance of alleles in these cohorts is unknown, this analysis captures the combined effect size of *de novo* mutations and inherited variants. On the liability scale, the effect sizes attributed to damaging PTVs in females (Z=0.21; 95% CI = 0.16 - 0.25; p =$3.81 \times 10^{-21}$; Bonferroni-corrected p = $1.71 \times 10^{-19}$) and males (Z=0.28; 95% CI = 0.22 - 0.33; p = $8.03 \times 10^{-36}$; Bonferroni-corrected p = $3.61 \times 10^{-34}$) were not significantly different ($Z_{Difference}$ = -0.072; 95% CI = -0.14 - 0.0018 ; p =0.056).

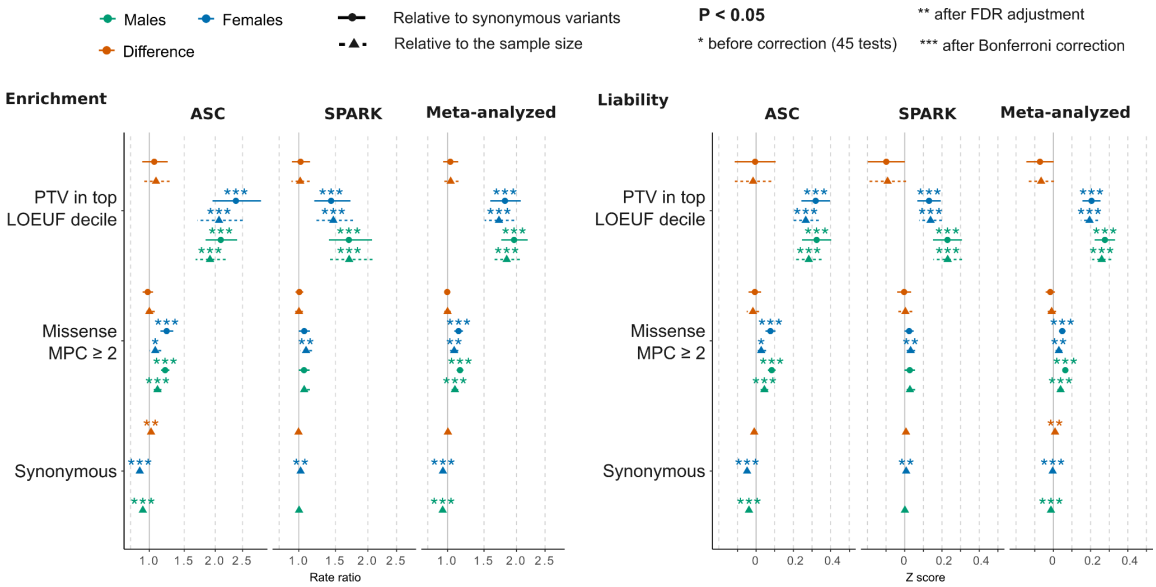


Figure S11. Enrichment of ultra-rare variants the case-control cohorts.

Ultra-rare variant rates in autism cases (ASC) and autistic probands without parental sequence data (SPARK) were compared to autism controls (ASC) or siblings not diagnosed with autism (SPARK). The error bars show the 95% confidence intervals of the effect size on the observed scale (left) and liability scale (left). The enrichment and liability were assessed relative to the sample size of the case/control cohorts (i.e. assuming that the expected rare variant burden per sample is similar across cohorts). To account for the differences in ultra-rare variant counts arising from the differences in ancestry, damaging missense and protein truncating variants were also compared to the expected rate ratio from synonymous variants (i.e. normalizing the average variant rates using synonymous variant counts).

### 3. Exome-wide burden in autistic individuals with *versus* without cognitive and motor difficulties

Here, we describe in more detail the findings presented in Figure 2 and related analysis in the remaining cohorts.

#### 4.1. DNM burden

##### 4.1.1. Damaging protein-truncating DNMs:

The effect size attributed to protein-truncating DNMs in autistic individuals with co-occurring motor or cognitive impairment was 0.60 in males (95% CI: 0.46 - 0.74; $p = 2.3 \times 10^{-17}$; Bonferroni corrected $p = 1.3 \times 10^{-15}$) and 0.55 in females (95% CI = 0.40 - 0.71; $p = 3.2 \times 10^{-12}$; Bonferroni corrected $p = 1.1 \times 10^{-10}$), without a significant difference (p = 0.67). *De novo* protein-truncating mutations increased the liability to autism without cognitive impairment or motor delay similarly (p = 0.82) in males (Z = 0.32; 95% CI: 0.16 - 0.47; $p = 5.9 \times 10^{-5}$; Bonferroni corrected p = 0.0032) and females (Z = 0.34; 95% CI: 0.17 - 0.51; $p = 6.2 \times 10^{-5}$; Bonferroni corrected p = 0.0034). When comparing autistic individuals with motor delay or cognitive impairment to those without these co-occurring difficulties, these DNMs had similar liabilities (p = 0.66) in males (Z = 0.45; 95% CI = 0.30 - 0.61; $p = 7.3 \times 10^{-9}$; Bonferroni-corrected $p = 4 \times 10^{-7}$) and females (Z = 0.39; 95% CI = 0.11 - 0.66; p = 0.0053; FDR-adjusted p = 0.019).

As mentioned in section 2.1, we found that SFARI genes but not the remaining autosomal genes show significant sex-differences in DNM rate ratios (Figure S7 in section 2.1 of the supplementary results). Since these genes are known to cause multiple developmental difficulties with high penetrance including cognitive impairment and motor delays, this alluded to the sex difference on the observed scale being driven by differences in the proportions of autistic females and males with these co-occurring difficulties. Indeed, we find that among autistics with motor and cognitive impairment (Figure S12), protein-truncating DNMs were enriched in autistic males (Rate ratio = 23.72; 95% CI = 7.78 - 117.82; $p = 6.3 \times 10^{-18}$; Bonferroni-corrected $p = 1.1 \times 10^{-15}$) and females (Rate ratio = 22.04; 95% CI 8.53 - 72.47; $p = 2.4 \times 10^{-16}$; Bonferroni-corrected $p = 4.3 \times 10^{-14}$) but not significantly different between the two sexes (p = 0.79).

Amongst those without these co-occurring conditions, protein-truncating DNMs were also significantly enriched in autistic males (Rate ratio = 9.43; 95% CI = 3.05 - 47.25; $p = 4.4 \times 10^{-7}$; Bonferroni-corrected $p = 7.9 \times 10^{-5}$) and females (Rate ratio = 7.56; 95% CI = 2.65 -

26.38; p = 1.5x10$^{-5}$; Bonferroni-corrected p = 0.0027) without a significant difference between the sexes (p = 0.36). As expected for this these known developmental genes DNMs were substantially enriched in those with motor or cognitive impairment *versus* those without co-occurring difficulties, in males (Rate ratio = 2.51; 95% CI = 1.73 - 3.68; p = 6.6x10$^{-7}$; Bonferroni-corrected p = 0.00012) and females (Rate ratio = 2.92; 95% CI = 1.55 - 5.69; p = 0.00034; FDR-adjusted p = 0.0020; Bonferroni-corrected p = 0.060). These findings are highlighted in Figure S12.

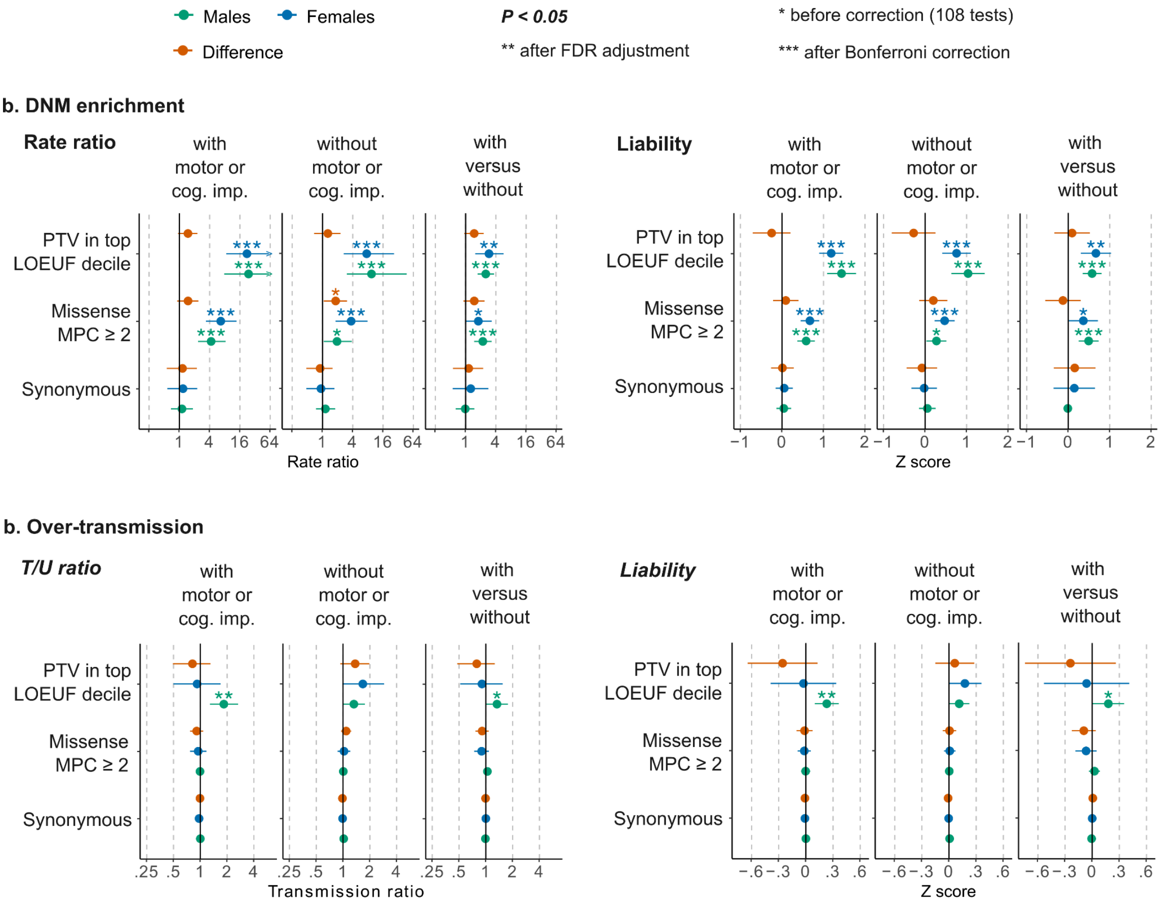


Figure S12: Enrichment of *de novo* mutations and rare inherited in SFARI genes in autistic individuals with and without motor and cognitive impairment in SPARK.

**a**, Enrichment in *de novo* mutations (DNM) in SFARI high-confidence and syndromic genes in two SPARK sub-cohorts of autistic individuals ascertained to have autism with or without co-occurring developmental delay or cognitive impairment (*versus* siblings), and directly between these two groups. The liability attributed to *de novo* mutations in these cohorts and phenotypic groups is shown on the right-hand side. **b,** Over-transmission analysis and liability attributed to inherited variants in the same cohorts. Similar tests for the complete

ASC and SPARK cohorts were presented in Figure S7a (DNMs) and Figure S9a (inherited alleles). Multiple testing correction was done jointly for these tests and those presented in Figure S16 (180 tests).

##### 4.1.2. Damaging missense DNMs:

Damaging *de novo* missense mutations increased the liability both in males (Z = 0.17; 95% CI = 0.077 - 0.25; p = 0.00024 ; Bonferroni-corrected p = 0.0016) and in females with motor or cognitive impairment (Z = 0.27; 95% CI = 0.16 - 0.34; p = $2.2x10^{-6}$; FDR-adjusted p =0.00012) - without a significant difference (p = 0.16). In those without motor or cognitive impairment, these DNMs significantly increased autism liability in females (Z = 0.016; 95% CI: 0.05 - 0.27 ; p =0.0043; FDR-adjusted p = 0.018) but not in males (Z = 0.029; 95% CI: -0.07 - 0.13 ; p =0.57), though the difference was not significant (p = 0.081). In direct comparisons between those with and without these difficulties, damaging missense DNMs had an increased liability to co-occurring difficulties in males (Z = 0.23; 95% CI = 0.10 - 0.35; p = 0.00032; Bonferroni-corrected p = 0.017) but the liability was not significantly increased in females (Z = 0.21; 95% CI = -0.012 - 0.43; p = 0.071). The difference was not statistically significant ($Z_{Difference}$ = -0.023; 95% CI = -0.28 - 0.23; p = 0.86). There was no significant sex difference in DNM enrichment in SFARI genes after accounting for multiple testing (Figure S12); these DNMs were more enriched in those with motor or cognitive difficulties versus those without these co-occurring difficulties.

##### 4.1.3 Imbalance of synonymous DNMs

As presented in the main text (Figure 2a), synonymous DNMs were more prevalent in autistic females without motor or cognitive impairment compared to those with these co-occurring difficulties as well as sex-matched siblings (Rate ratio = 1.23; 95% CI = 1.082 - 1.38; p = 0.00099). We examined the burden and liability in ultra-rare DNMs in this sub-cohort (Figure S13) and found better-balanced DNM counts between the probands and siblings, yet not completely controlling the spurious association (Rate ratio = 1.22; 95% CI = 1.038 - 1.43; p = 0.015), which is likely to be an artifact of the diverse ancestries included in the sample.

We then examined whether limiting the analysis to a stringently ancestry-matched set of 868 autistic females and 2,235 siblings could resolve this issue (see section 2.4 of the extended methods). This ancestry matching alone resulted in better-balanced DNM counts between the probands and siblings with some residual imbalance (Rate ratio = 1.17; 95% CI = 1 - 1.37; p = 0.047). Next, we combined the two filtering strategies, i.e. by evaluating ultra-rare DNMs in samples well-matched on genetic ancestry. Here, synonymous DNMs were not associated with autism (Rate Ratio = 1.09; 95% CI = 0.88 - 1.35; p = 0.42).

Lastly, we turned to protein-truncating DNMs to see whether the estimated enrichment in protein-truncating DNMs differed with more stringent frequency and ancestry matching. The rare DNM rate ratio was 2.65 for rare DNMs in the complete cohort (95% CI = 1.61 - 4.37; p = $6.2 \times 10^{-5}$) *versus* 2.68 in ancestry-matched samples (95% CI = 1.47 - 4.9; p = 0.00067). The ultra-rare DNM rate ratio was 2.66 in the complete cohort (95% CI = 1.6 - 4.42; p = $8.4 \times 10^{-5}$) *versus* 2.57 in ancestry-matched samples (95% CI = 1.4 - 4.7; p = 0.0018). Thus, the estimates of protein-truncating DNM enrichment obtained when examining the full cohort were robust to these differences in genetic ancestry and were slightly more precise.

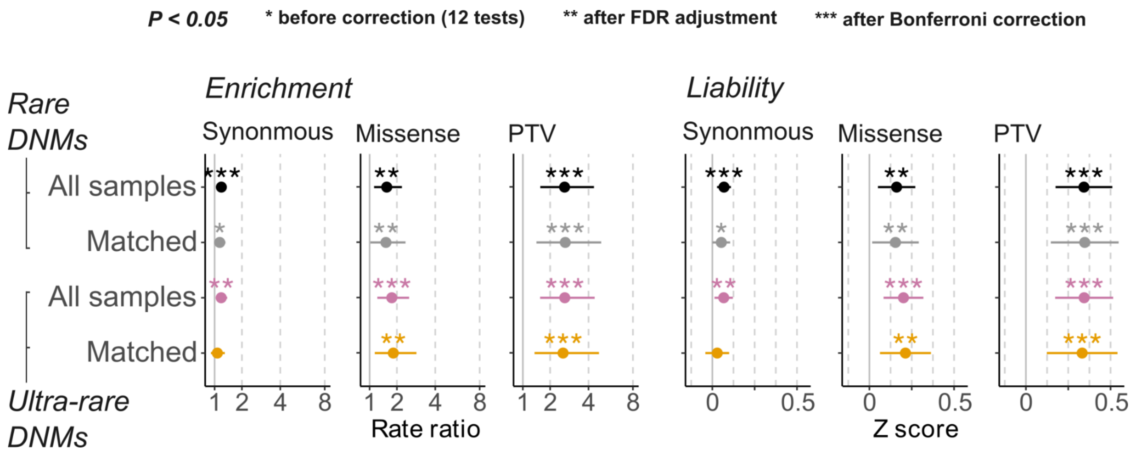


Figure S13: Enrichment of *de novo* mutations in ancestry-matched subset of autistic females without cognitive or motor impairment and siblings.

Enrichment and liability in 'All samples' was tested in 1,464 autistic females without motor or cognitive impairment and sex-matched siblings from different genetic ancestry groups (see Table S4). 'Matched' indicates comparisons between 868 female probands of European ancestry and 2,235 ancestry-matched siblings (see Extended Methods section 2.4).

### 4.3. Over-transmission

#### 4.3.1. Inherited protein-truncating variants:

In autistic individuals with motor or cognitive impairment (Figure 2b), inherited PTV liability was increased in males (Z = 0.12; 95% CI = 0.076 - 0.16; p = $1.1 \times 10^{-7}$; Bonferroni-corrected p = $6 \times 10^{-6}$) but not significantly so in females (Z = 0.063; 95% CI =

-0.0060 - 0.13 ; p = 0.073), yet the difference was not significant ($Z_{Difference}$ = -0.057; 95% CI = -0.14 - 0.025; p = 0.17). In the other group (without co-occurring difficulties), inherited PTVs had comparable effect sizes (p = 0.54) in males (Z = 0.06; 95% CI: 0.25 - 0.10; p = 0.0049; Bonferroni-corrected p = 0.046) and in females (Z = 0.08; 95% CI: 0.25 - 0.14; p = 0.00085; FDR-adjusted p = 0.019). Notably, ultra-rare inherited variants ascertained in a separate cohort of autistic individuals with sequence data from one parent showed similar patterns in those with motor or cognitive difficulties but not in those without co-occurring difficulties (Figure S14). Specifically, inherited PTVs conferred significant liability in males with motor or cognitive co-occurring (Z = 0.13; 95% CI = 0.057 - 0.20; p = 0.00052; Bonferroni-corrected p = 0.14) but not females (Z = 0.083; 95% CI = -0.048 - 0.21; p = 0.21), yet without a significant sex difference (p = 0.54), whereas the liability in the other group was not significantly increased from the population mean in both sexes (p > 0.11). The liability attributed to inherited PTVs did not differ significantly between those with and without co-occurring difficulties (p > 0.31).

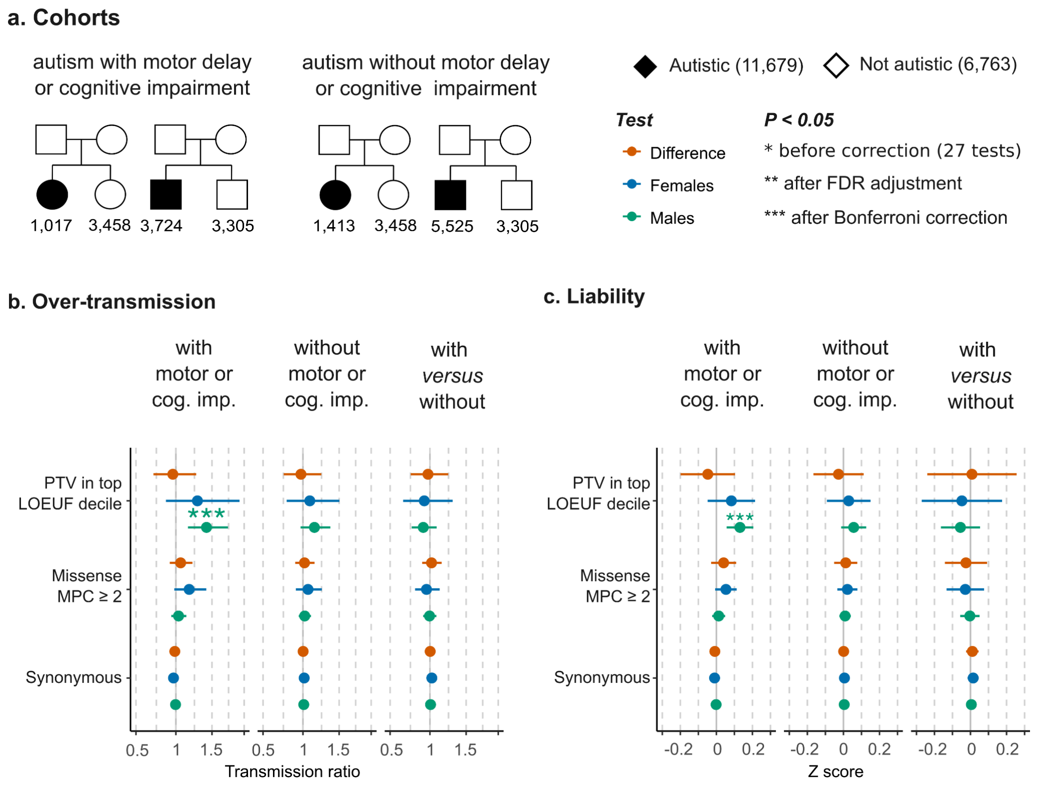

Figure S14. Over-transmission of ultra-rare inherited variants in SPARK individuals with one sequenced parent, stratified by motor and cognitive impairment.

**a**, The sample size of the different cohorts included in this analysis (note that the transmission analysis is done in probands only). **b**, Over-transmission analysis showing the ratio between rare parental alleles transmitted to autistic individuals with or without motor or cognitive impairment and untransmitted alleles. Transmission ratios were also compared between these two groups (see the Methods). **c**, The liability attributable to inherited rare variants.

##### 4.3.2. Inherited missense variants:

Among those with motor or cognitive impairment (Figure 2c), inherited damaging missense variants conveyed significant liability in males (Z = 0.015; 95% CI = 0.0030 - 0.027; p = 0.015; FDR-adjusted p = 0.047) but not in females (Z = -0.011; 95% CI = -0.30 - 0.0083; p = 0.27). The difference was significant only before correction for multiple testing (Z = -0.026; 95% CI = -0.048 - 0.0032; p = 0.025). The liability attributed to inherited damaging missense variants in females without cognitive impairment was not significantly increased (Z = 0.00035; 95% CI = -0.13 - 0.02). These variants increased the liability in males (Z = 0.01; 95% CI = 0.0006 - 0.02; p = 0.037) but this did not pass FDR-adjustment (p = 0.11). There was no significant difference in the average liability of inherited missense variants between those with and without co-occurring difficulties (p > 0.43). Ultra-rare damaging missense variants in SPARK individuals with sequence data from one parent did not convey significant liability (Figure S14).

#### 4.4. Ultra-rare variants in the remaining SPARK individuals without parental sequence data

There were no significant sex differences in the enrichment and liability damaging of protein-truncating and missense ultra-rare variants among the remaining probands and siblings from SPARK (Figure S15). The effect size of ultra-rare protein-truncating variants (adjusted for synonymous variants) was similar in males (Z = 0.27; 95% CI = 0.19 - 0.34; p = $1.4 \times 10^{-11;}$ Bonferroni-corrected p = $6.4 \times 10^{-10}$) and females with motor and cognitive difficulties (Z = 0.22; 95% CI = 0.15 - 0.30; p = $2.2 \times 10^{-8}$; Bonferroni-corrected p = $1.0 \times 10^{-7}$). Damaging missense variants had an effect sizes of 0.039 in males (95% CI = 0.0096 - 0.068; p = 0.0094; FDR-adjusted p = 0.033; Bonferroni-corrected p = 0.42) and 0.036 in females with cognitive impairment or motor delay (95% CI = 0.0020 - 0.069; p = 0.038; FDR-adjusted p = 0.095).

Among those without motor and cognitive difficulties, PTVs had an effect size of 0.16 in males (95% CI = 0.074 - 0.24; p = 0.00021; Bonferroni-corrected p = 0.0096) compared to 0.040 in females (95% CI = -0.056 - 0.42; p = 0.42) although the difference was not statistically significant (p = 0.068). There was no significant enrichment in damaging missense variants in autistic individuals without these co-occurring conditions. When the two groups were compared (with *versus* without motor or cognitive impairment), the effect size of PTVs was 0.19 in males (95% CI = 0.089 - 0.29; p = 0.00023; Bonferroni-corrected p = 0.010) and 0.35 in females (95% CI = 0.17 - 0.53; p = 0.00017; Bonferroni-corrected p = 0.0079), and it was not significantly different (p = 0.14). Missense variants had a slightly higher liability to autism with (vs. without) motor or cognitive impairment in males (Z = 0.048; 95% CI = 0.0046 - 0.092; p = 0.03) but the liability did not differ between the two groups after correction for multiple testing (FDR-adjusted p = 0.08).

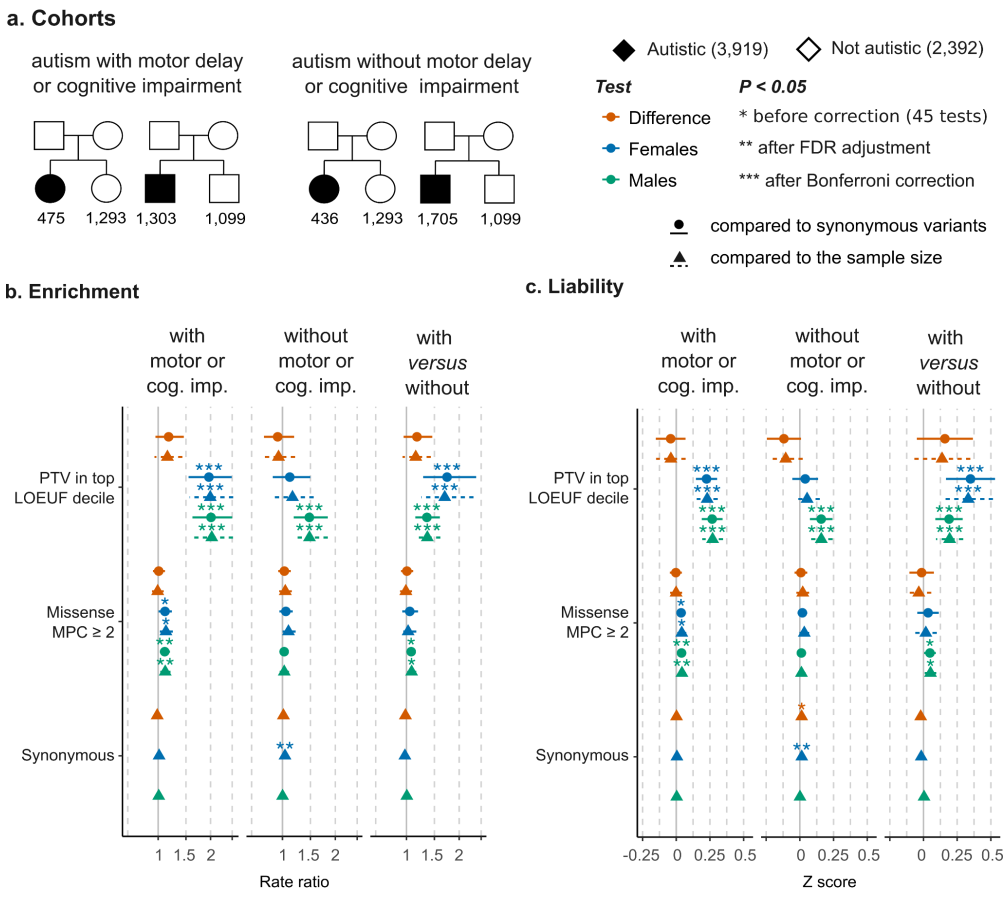


Figure S15. Enrichment of ultra-rare inherited variants in SPARK individuals without sequenced parents, stratified by motor and cognitive impairment.

The enrichment of ultra-rare variants in a cohort of autistic probands without parental sequence data in SPARK (**a**) was examined on the observed scale (**b**) and liability scale (**c**).

Variant rates were compared between autistic stratified by co-occurring cognitive or motor impairment and siblings not diagnosed with autism as well as between the two autism sub-cohorts (‘with *versus* without’) . The error bars show the 95% confidence intervals of the effect sizes. The enrichment and liability were assessed relative to the sample size of the case/control cohorts (i.e. assuming that the expected rare variant burden per sample is similar across cohorts). To account for the differences in ultra-rare variant counts arising from the differences in ancestry, damaging missense and protein truncating variants were also compared to the expected rate ratio from synonymous variants (i.e. normalizing the average variant rates using synonymous variant counts). This analysis is related to the analysis presented in Figure S11.

### 6. Gene set enrichment relative to matched genes

#### 6.1. High-confidence and syndromic autism predisposition genes

In the full ASC and SPARK cohorts, the enrichment of damaging protein-truncating DNMs (rate ratio in autistic females and males *versus* sex-matched siblings), and the sex-difference in DNM enrichment (rate ratio in autistic females *versus* autistic males), were significantly higher than what is expected from matched genes, i.e, significantly higher that the rate ratio calculated from random genes of similar LoF-constraint, sex-averaged expression and coding length profiles (Figure S16a). The sex-difference in SPARK relative to matched genes was driven by autistic probands who have motor or cognitive impairment, and was not seen in those co-occurring difficulties. We have shown that males and females in the 'motor and cognitive impairment' group do not show a significant sex difference in DNM rates *per se*, i.e. relative to what is expected from the sample size of the cohort (Figure S12). This apparent discrepancy may be due to an ascertainment bias in the discovery cohorts behind some of these genes (more females with cognitive impairment and *de novo* protein-truncating mutations). A significant sex-bias in damaging missense DNM enrichment relative to matched genes was seen in SPARK only, and was driven by DNMs seen in autistic individuals without motor or cognitive impairment.

These observed sex differences in DNM risk ratios did not translate to a significant sex-bias on the liability scale, i.e. the 'excess' autism liability conferred by these variants on top of what is expected from matched genes was comparable between males and females (Figure S16b). Protein-truncating DNMs in SFARI genes increased the liability to autism with cognitive and motor impairment by 0.99 units more than a matched gene set in males (95% CI = 0.64 - 1.34; p = $2.6\times10^{-8}$; Bonferroni-corrected p = $4.7\times10^{-6}$) and 0.86 units more that matched genes in females (95% CI = 0.59 - 1.14; p = $1.1\times10^{-9}$; Bonferroni-corrected p = $2\times10^{-7}$), without a significant sex difference (p= 0.58). In the 'autism without cognitive & motor impairment' group, the sex difference between males (0.89; 95% CI = 0.49 - 1.30; p = $1.8\times10^{-5}$; Bonferroni-corrected p = 0.0032) and females (0.52; 95% CI = 1.7 - 0.86 ; p = 0.0032; FDR-adjusted p= 0.18; Bonferroni-corrected p = 0.58) was also not significant (p = 0.17). Inherited variants did not show significant differences in over-transmission rates and liability compared to matched genes (Table S14).

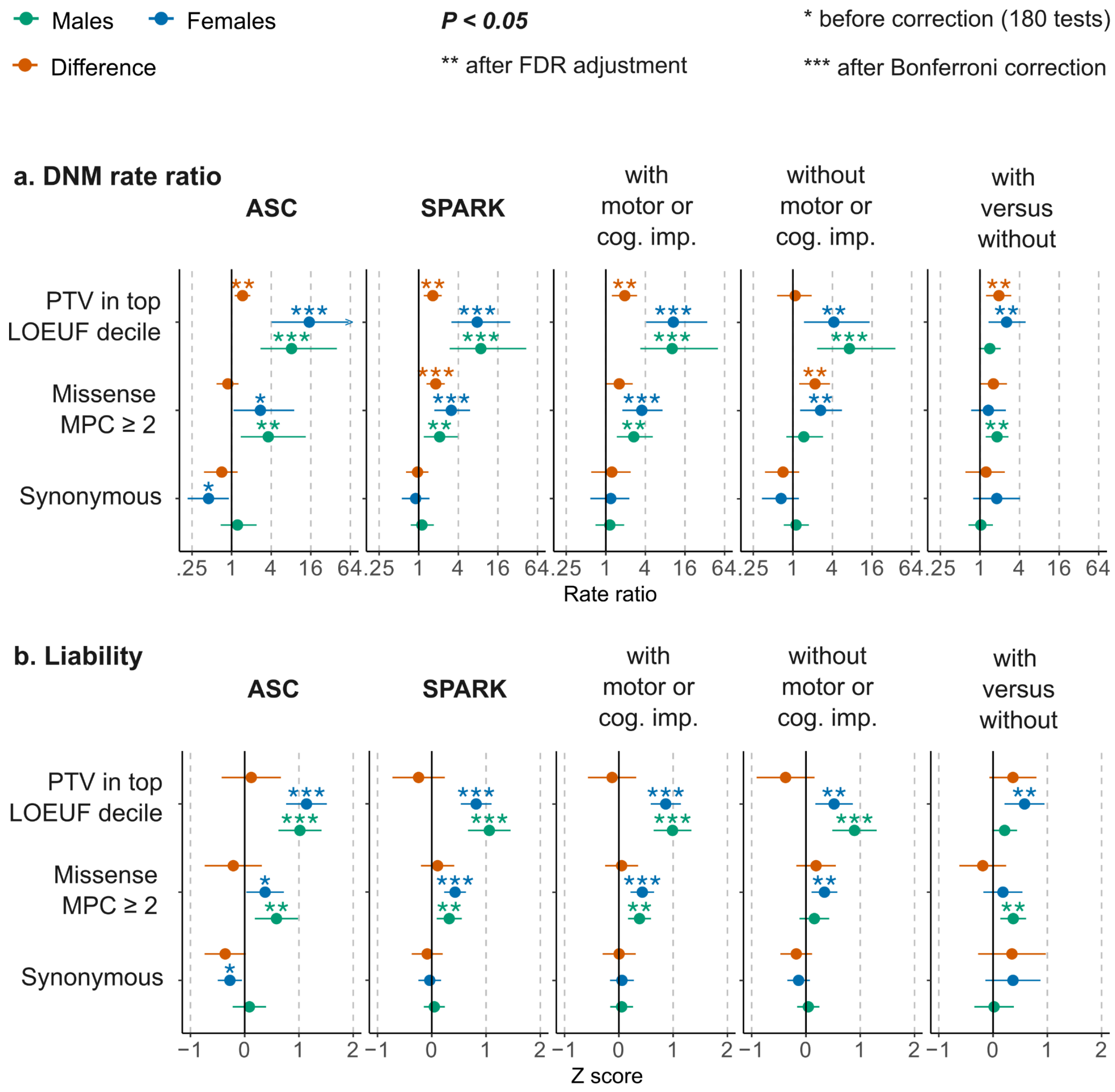


Figure S16. Enrichment and liability of *de novo* mutations in 354 SFARI genes compared to matched genes.

**a**, The enrichment (DNM rate ratio) attributed to *de novo* mutations in SFARI high-confidence and syndromic genes, compared to genes matched on LoF-constraint, sex-averaged expression and coding length. The enrichment was examined in trio-sequenced individuals from the ASC and SPARK (all individuals), in two SPARK sub-cohorts of autistic individuals ascertained to have autism with or without co-occurring developmental delay or cognitive impairment (versus siblings), and finally compared directly between the latter two groups. **b**, Liability compared to matched genes. See Table S17 for over-transmission analysis.

#### 6.2. Genes with male biased expression in the fetal cortex

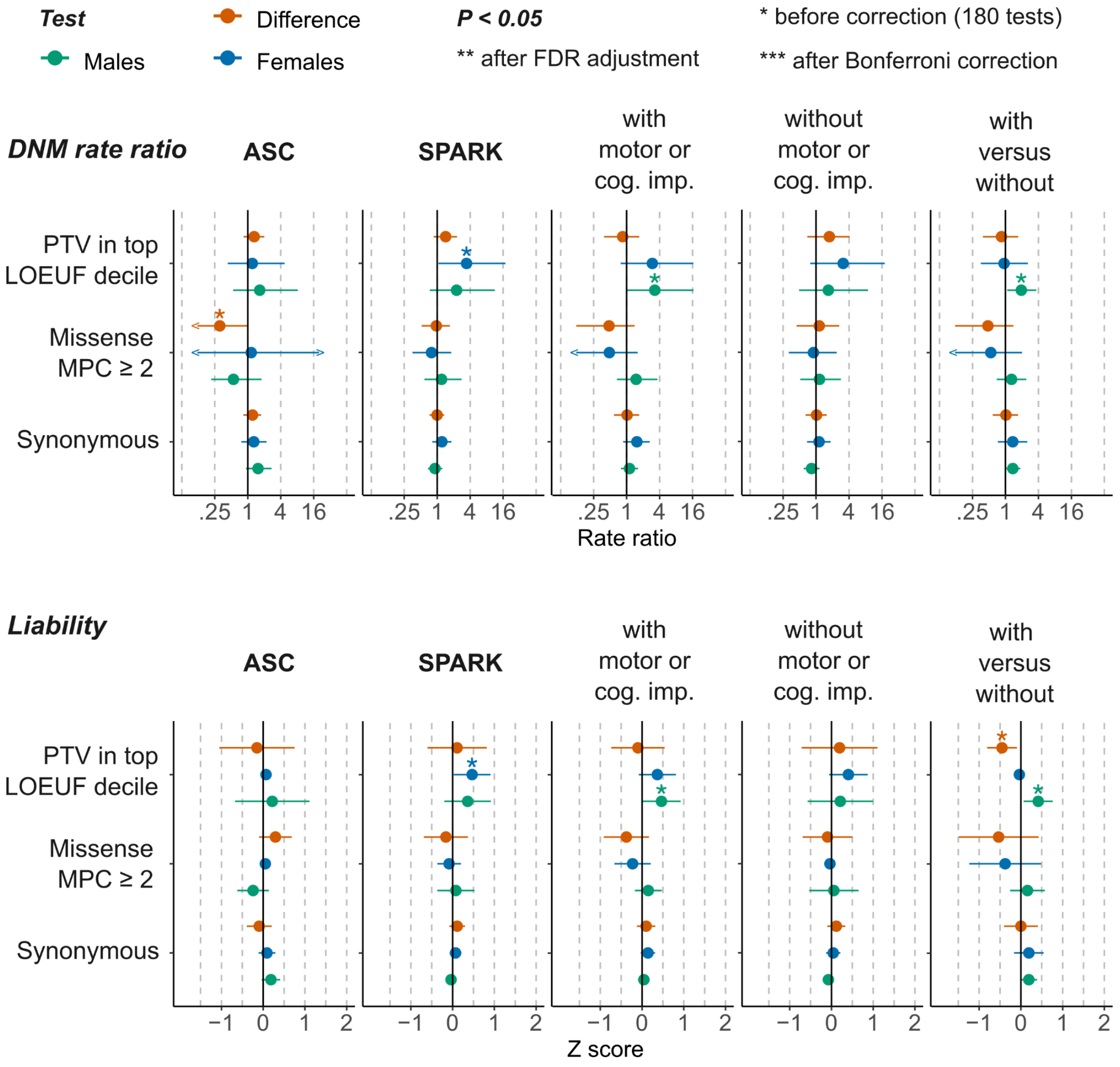


Figure S17. Burden and liability of *de novo* mutations in 856 genes with male-biased expression in the human fetal cortex relative to matched genes.

This figure presents a complementary analysis to the one shown in Figure 3. Here, de novo mutation rates are compared to those expected based on matched genes instead of based on the relative sample size of probands and sex-matched siblings. See Methods and the legend of Figure 3 for more details and Table S7 for the complete results including inherited variants.

### 7. Supplementary Tables

Tables S1-S4 present the sample size before and after QC, and are contained in the extended methods. Additional extended supplementary tables are provided in a separate ‘xlsx’ file. Tables S5-S20 present the enrichment and liability analysis outcomes presented in the main and supplementary figures.
